# Active ingredient ‘Better gut microbiome function’

**DOI:** 10.1101/2020.11.09.20228445

**Authors:** Kathrin Cohen Kadosh, Melissa Basso, Paul Knytl, Nicola Johnstone, Jennifer Y F Lau, Glenn Gibson

## Abstract

**Background:** The human gut microbiome and its effect on brain function and mental health is emerging as an area of intensive research. Both animal and human research point towards adolescence as a sensitive period when the gut-brain axis is fine-tuned, and when we can use dietary intervention to change the microbiome, with long-lasting consequences for mental health. Here we report the results of a systematic review/meta-analysis of microbiota-targeted (psychobiotics) interventions on anxiety in youth, together with a summary of consultation work of youth with lived experience.

**Methods:** Seven databases were searched (no date cut-offs), and controlled trials in clinical and healthy human samples (age range: 10-24) seeking to reduce anxiety were included. All data on between group-differences post intervention and outcomes were extracted as standard mean differences (SMDs) and pooled together based on a random-effects model.

**Findings:** 5416 studies were identified, 14 were eligible for the qualitative summary, of which 10 were included in the meta-analyses (total of 324 experimental and 293 control subjects). The heterogeneity I^2^ was12% and the pooled SMD was −0.04 (95% CI: −0.21, 0.14). One study presented with low bias risk whereas 5 with high and 4 with uncertain risk, accounting for that, sensitivities analysis revealed a SMD of −0.16 (95%CI: −0.39, 0.06).

**Interpretation:** There is currently limited evidence for use of psychobiotics to treat anxiety in youth. However, future progress will require a multidisciplinary research approach, which gives priority to specifying mechanisms in the human models, providing causal understanding and addressing the wider context.

## Introduction

This report summarises our work into the effectiveness of using dietary interventions to improve gut microbiome function in young people aged 14-24 years, with a view to reducing anxiety. It combines results of a systematic review and meta-analysis of the relevant scientific literature with a summary of consultation and focus group work of young people with lived experience of anxiety (N= 45).

How does what we eat affect how we feel? Intuitively, it seems right to expect that the one thing that we change about our bodies, on a daily basis, will also have a substantial impact on how we feel. Indeed, in their contribution to this report, just over two thirds of young people with lived experience of clinical levels of anxiety declared that they had previously attempted to influence mental health and well-being with dietary interventions, such as reducing sugar intake or increasing the amount of daily fruit and vegetables eaten, or introducing supplements, such as probiotics and prebiotics, often following the suggestions of family, friends or a GP. For example, Jane^1^ said:

> *“I have anxiety and have been taking probiotics since January for a combo of reasons (gut health, acne, mental health). 100% have noticed a difference and agree that more research in this area is needed”*.
>
> *Mehta added: “I’m really interested in how gut health affects us and I notice it’s positive effects personally too, particularly when switching between eating meat and not eating meat and consumption of sugar*.*”*

Appreciating the importance of our diet on health and well-being has a long-standing tradition. In a bold statement 2500 years ago, Hippocrates, the father of medicine, stated that *all* disease begins in the gut. A considerable body of research in both human and animal models in the last decade has now largely borne him out, with research repeatedly highlighting the important role that gut microbiota (i.e. the microorganisms that live in the digestive system) play in regulating the brain and subsequent behaviour (Sarkar, Lehto et al. 2016, Sarkar, Harty et al. 2020). This has become a particular research focus within the context of mental health problems such as anxiety or depression (Mayer 2011, Cryan and Dinan 2012, Foster and McVey Neufeld 2013). The gut and the brain are intimately connected via the gut-brain axis, which involves bidirectional communication via neural, endocrine and immune pathways, including the vagus nerve (**Figure 1**), (Grossman 1979, Grenham, Clarke et al. 2011, Mayer, Knight et al. 2014).

**Figure 1:**
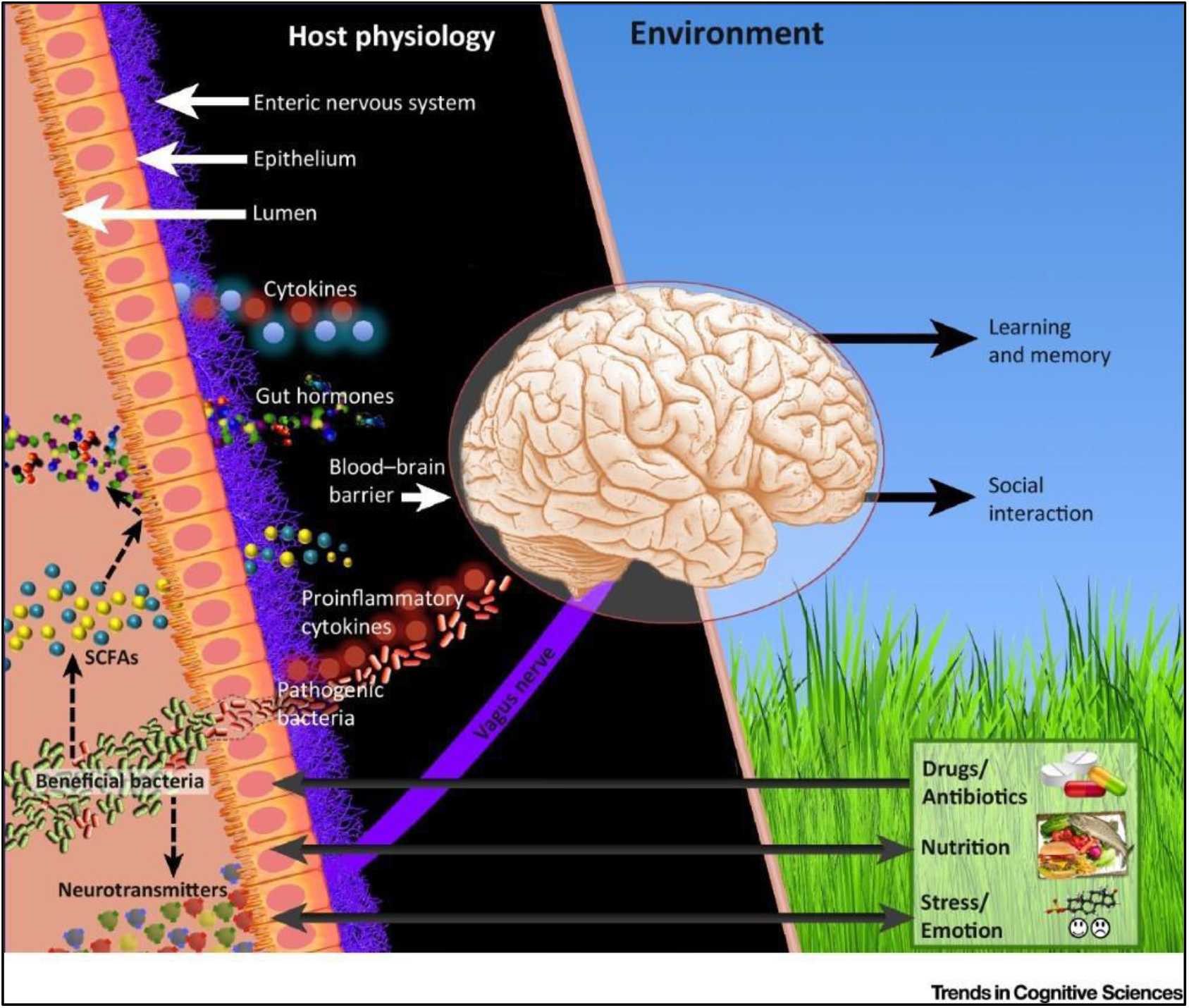
Bacteria–Brain–Behaviour Relationships. Gut bacteria communicate with the brain through various channels. The vagus nerve enables bidirectional communication between the gut and brain. Bacteria produce neurotransmitters as metabolites, and these are able to regulate activity in proximal neurons of the enteric nervous system. Bacteria also produce short-chain fatty acids (SCFAs) via the fermentation of non-digestible dietary fibres. SCFAs can modulate neural activity in the host directly, but they also stimulate the release of gut hormones such as glucagon-like peptide 1 (GLP-1) and peptide tyrosine tyrosine (PYY) which control host appetite and satiety. Beneficial bacteria can also trigger the release of anti-inflammatory cytokines which decrease local and systemic inflammation. However, pathogenic bacteria migrating out of the gut (through a weakened gut barrier) trigger proinflammatory cytokine release. Higher levels of inflammation are correlated with depression, and ‘leaky gut’ is also associated with depression. Furthermore, experiencing stress – and the attendant glucocorticoid response – can also weaken the gut barrier, potentially worsening the inflammatory pathophysiology of emotional disorders. Diet and the use of drugs and antibiotics also directly modulate composition of gut bacteria, which in turn alter the physiological signals generated. Overall, these bacterially generated signals modulate the central nervous system and affect emotion, cognition, and behaviour. **Re-printed and adapted with permission from Sarkar, 2018**.

Animal research has shown that adolescence is a critical window where microbiota help fine-tune the gut-brain axis (McVey Neufeld, Luczynski et al. 2016) and, given the link between the gut-brain axis and mental health could be one factor for a significant increase in mental health problems during this period (Paus, Keshavan et al. 2008, Keshavan, Giedd et al. 2014). This makes a healthy gut microbiome an important and possibly time-sensitive active ingredient, as interventions during this period may have long-lasting consequences both at the gut microbiome and brain level. However, evidence from human participants for the effectiveness of microbiota-targeted interventions to improve mental health is still outstanding.

One effective way of changing the gut microbiome and gut-brain axis is via nutrition, and it has been shown that drastic changes in diet can alter bacterial diversity in days (David, Maurice et al. 2014). Another method for improving the gut microbiome is the use of psychobiotics. Psychobiotics refer to active compounds capable of acting on the nervous system, consequentially shaping psychological processes, behaviour, and ultimately exerting health benefits in psychiatric conditions (Sarkar, Lehto et al. 2016, Sarkar, Harty et al. 2018). Research has also shown that bacterial dysbiosis (i.e. aberrant functioning of different microorganisms in the gut) may lead to the psychological abnormalities that characterise anxiety (Foster and McVey Neufeld 2013). Therefore, manipulation of the gut microbiome via psychobiotics may present a promising new avenue for treatment and prevention of anxiety in young people (Cryan and Dinan 2012, Sarkar, Lehto et al. 2016) (see **Figure 2** for a proposed pathway). It is not surprising therefore that the microbiome, and its effect on behaviour and mental health has captured the interest and imagination of scientists, clinicians, health professionals and the wider public alike. As a result, research in this relatively new area has intensified, and young people in particular seem very enthusiastic about this line of research, as they see it as an innovative, approachable and critically a non-stigmatising intervention approach. Sonia wrote in her initial assessment of the consultation:

**Figure 2:**
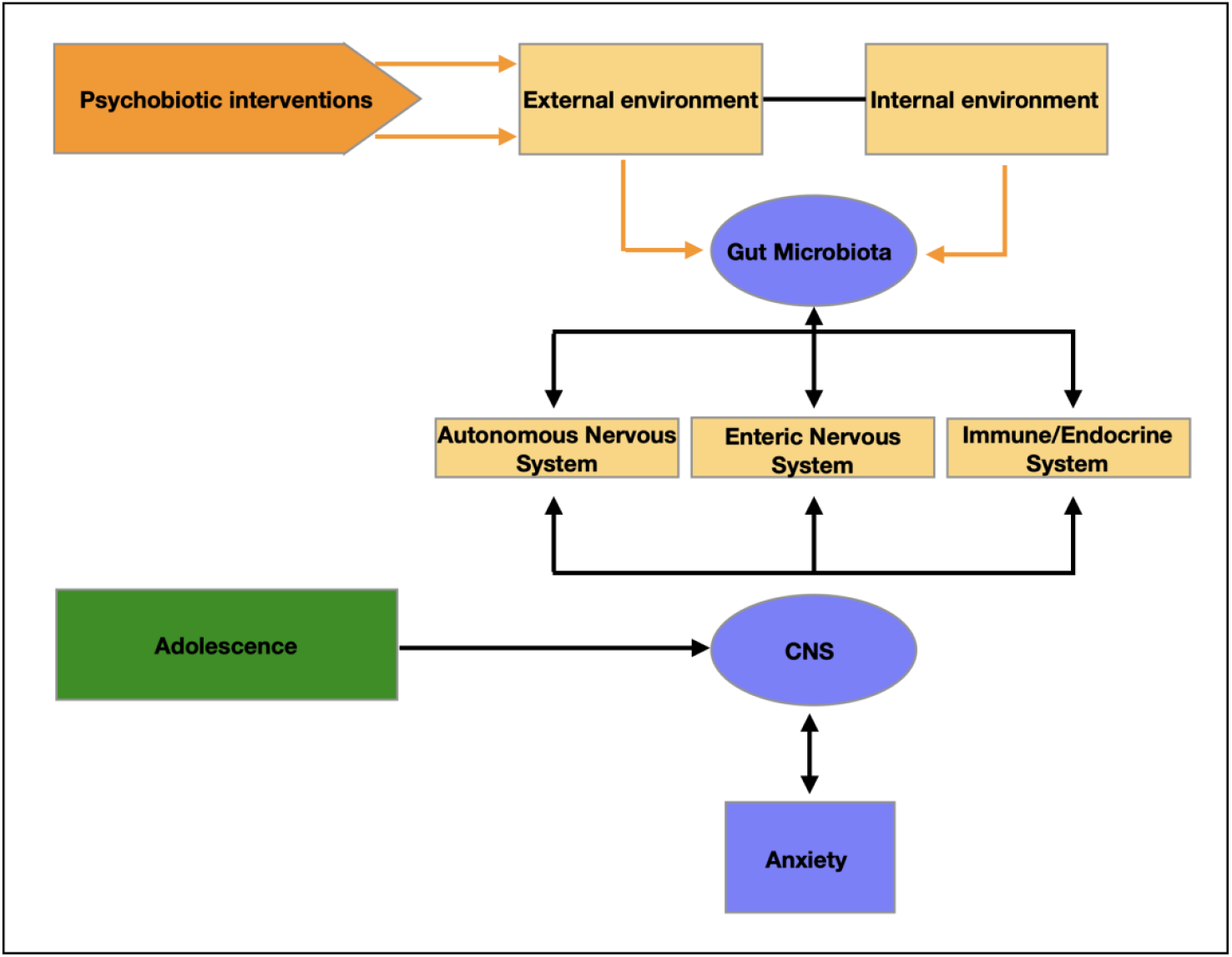
Proposed intervention pathway for the active ingredient. Adolescence is a time period of ongoing neuro-cognitive development, allowing brain structures and circuities to flexibly adapt-or maladapt to the environment. In this context, gut microbiota might play be a causal role as a mediator between the environment and the CNS via multiple pathways. As easily manipulated throughout diet, it could be a promising and cheap therapy target in the redirection of neurodevelopmental trajectories and improving the mental health outcome for the individual.

> *“This is so interesting. We do not receive many requests on the topic of diet, which is a shame because it was one of top 25 questions in our ‘Right People, Right Questions’ (RPRQ) project”*.

Note that the RPRQ project was conducted by the McPin foundation whereby children and young people (including those with lived experience of mental health problems), families, professionals and academics identified the top priorities for research in children and young people’s mental health. The fact that diet was selected as a top priority further underlines the importance of gaining a better understanding how it can be used to improve mental health.

In the following, we provide a brief overview of the research conducted in animal models (summarised in **Table 1**) before presenting the results of our systematic review. We will then make specific suggestions for future research, which integrate the input by the young people with lived experience.

**Table 1.**
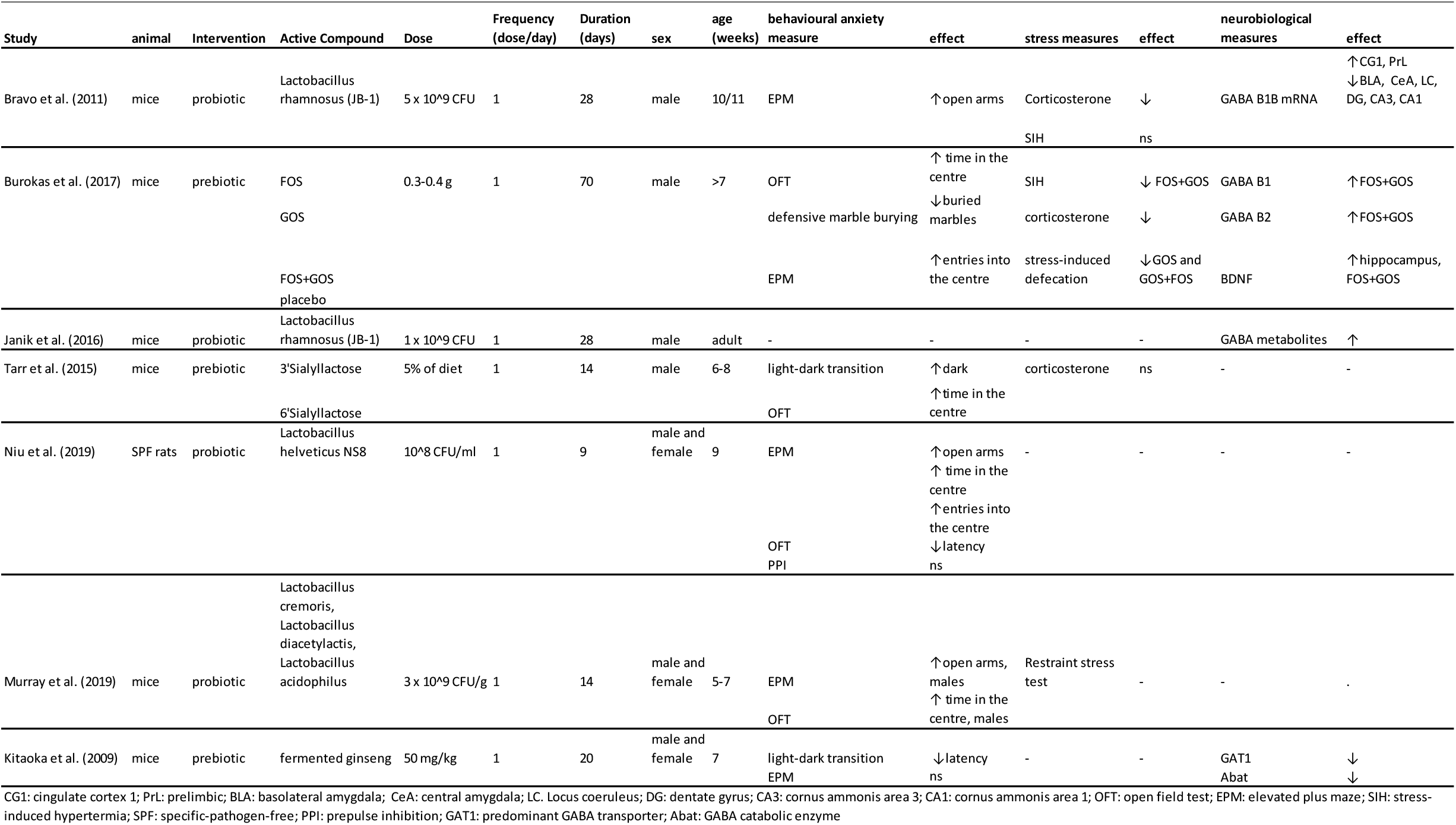
Characteristics of the pre-clinical intervention studies

One class of psychobiotics are known as probiotics – live gut bacteria capable of releasing neuroactive substances (Dinan, Stanton et al. 2013), depending on the bacterial strain. For example, the *Bifidobacterium* family is linked to GABA expression in the brain, whereas the *Enterococcus* and *Streptococcus* families are shown to produce serotonin and *Lactobacillus* are linked to GABA and acetylcholine (Sarkar et al., 2016). Research has consistently outlined the psychotropic effects of probiotics. For example, in an animal study, Barrett et al. (Barrett, Ross et al. 2012) showed that *Lactobacillus (L). brevis* and *Bifidobacterium (B*.*) dentium* increased GABA concentrations in-vitro, a finding which was replicated in an in-vivo model which showed that a *L. brevis* ingested strain regulated emotional behavioural and central GABA receptor expression in a mouse model (Bravo, Forsythe et al. 2011). In this study, mice fed *L. rhamnosus* exhibited reduced GABA mRNA expression in the amygdala, along with lower levels of stress-induced corticosterone and anxiety- and depression-related behaviour. Another study replicated this finding (Janik, Thomason et al. 2016) and showed that increases in GABA metabolites in the brain were evident after about 4 weeks, a lag comparable with other pharmaceutical interventions, such as serotonin-reuptake inhibitors (Kodish, Rockhill et al. 2011). Notably, the same study pointed out that re-colonisation of gut bacteria in adolescent germ-free (GF) mice (i.e. mice without any microorganisms living in or on them) was not sufficient to reverse anxiety-like behaviour, once again suggesting that early deficits in gut microbiota may not be reversible.

Along with probiotics, some prebiotics have lately been also classified as psychobiotics. Prebiotics are specific non-digestible food ingredients (including non-digestible oligosaccharides) which selectively feed intrinsic beneficial bacteria, consequentially stimulating their growth and activity with notable effects on brain development and function (Gibson, Scott et al. 2010, Sarkar, Lehto et al. 2016). To date, fructooligosaccharides (FOS) and galactooligosaccharides (GOS) have been the most studied ones, showing promising effects in animal and human trials (Tarr, Galley et al. 2015, Burokas, Arboleya et al. 2017). In the context of cognitive function specifically, Tarr et al. (2015) have shown that milk oligosaccharides administration can prevent stress-induced dysbiosis and anxiety-like behaviour in mice. Likewise, Burokas et al. (2017) have reported chronic combined FOS and GOS supplementation in mice to have anxiolytic and antidepressant effects, as well as to reduce corticosterone stress response. In addition, prebiotics have been shown to modulate hippocampal and hypothalamus gene expression, and lead to SCFAs concentration changes which positively correlate with the behavioural effects.

A further example of the developmental impact of gut microbiome composition on anxiety has recently been demonstrated in a series of animal models. One study reported that rats’ ingestion of pre-gestational *Lactobacillus helveticus* resulted in offspring which then displayed lower rates of anxiety-like behaviour in adolescence (Niu, Liang et al. 2020). In young rats who were experimentally stressed, those who received a placebo presented with abnormal pubertal timing, whilst those fed *Lactobacillus rhamnosus* and *Lactobacillus helveticus* presented normal timing (Cowan and Richardson 2019). In another study, adolescent mice were injected with the toxin lipopolysaccharide which caused immediate and temporary depression- and anxiety-like behaviour, but also increased stress-sensitivity in adulthood. Compared to placebo, mice who received a mixture of *Lactococcus lactis, L. cremoris, L. diacetylactis, L. acidophilus* around the time of toxin administration displayed shorter duration of immediate negative effects and also did not display as severe stress-responses in adulthood (Murray et al., 2019). These findings support an earlier landmark study (Sudo et al., 2004) which found that adult GF mice had enhanced stress responses which could be reversed by gut colonisation with *Bifidobacterium infantis*. Crucially, the earlier in the lifespan this intervention took place, the more fully normal stress response was restored. Thus, if these promising effects translate to humans, psychobiotics present candidate ingredients which could provide a measure of protection against stress-induced anxiety in adolescents which may carry over into adulthood.

### The effects of probiotic intake on anxiety and stress in adolescence

Here, we report results of a systematic review and meta-analysis which aimed to determine whether psychobiotics conferred either a protective or restorative effect on anxiety and stress in humans aged 14-24. Seven electronic data bases were searched (Cochrane Library, Embase, Medline, PubMed, Psychinfo, Scopus, and Web of Science) using search terms such as minors, adolescent, child, development, paediatric, puberty, young adult, teen, young men, young women, or undergraduate student (please see **Appendix 1** for a complete description of methods and **Appendix 2** for the full search list). The output of this search was 5416 studies, while subsequent screening provided 14 studies, which measured anxiety or stress either directly or as secondary outcomes (**Figure 3**). The results are summarised in **Table 2** and reviewed below.

**Table 2.**
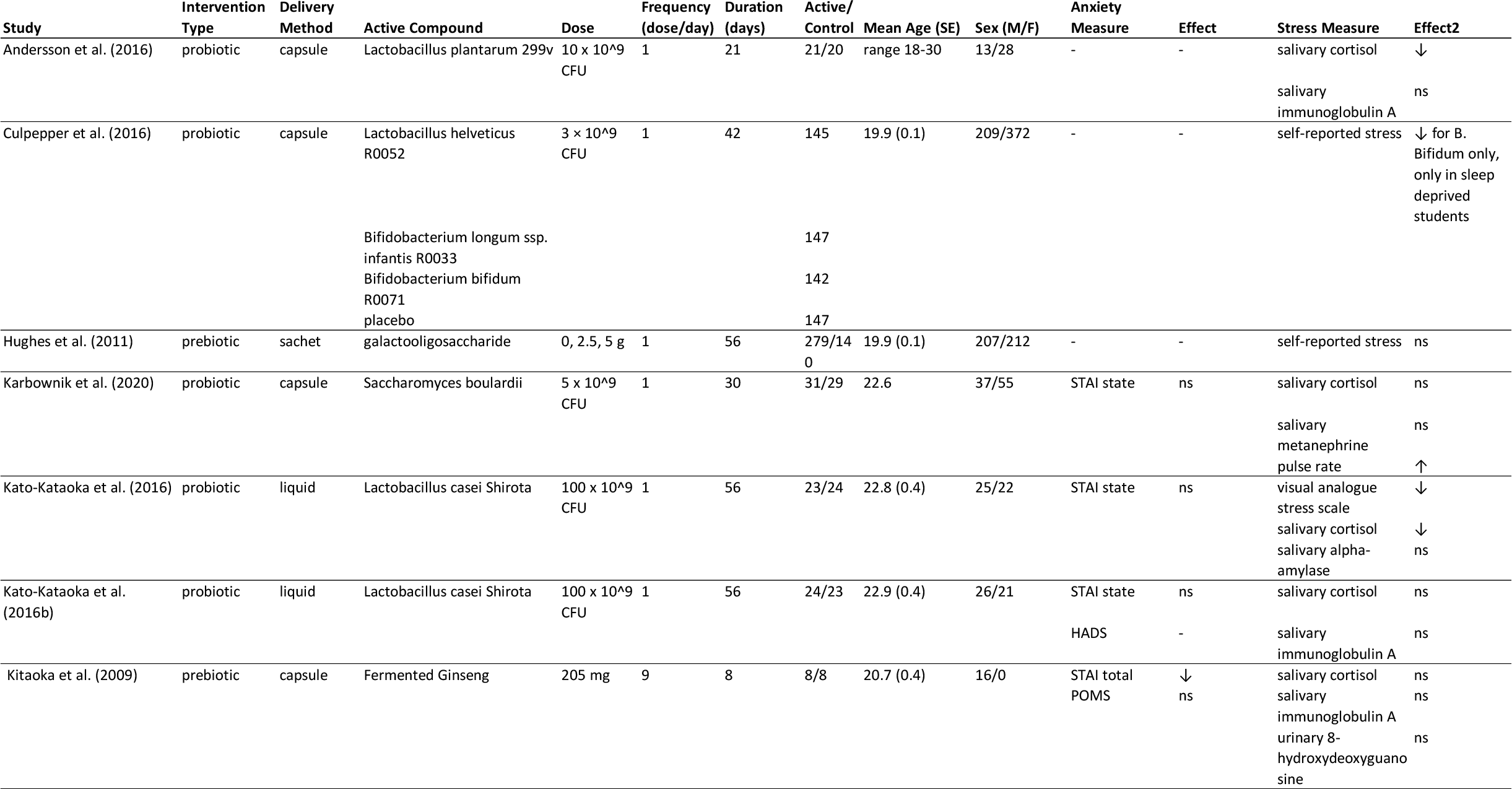
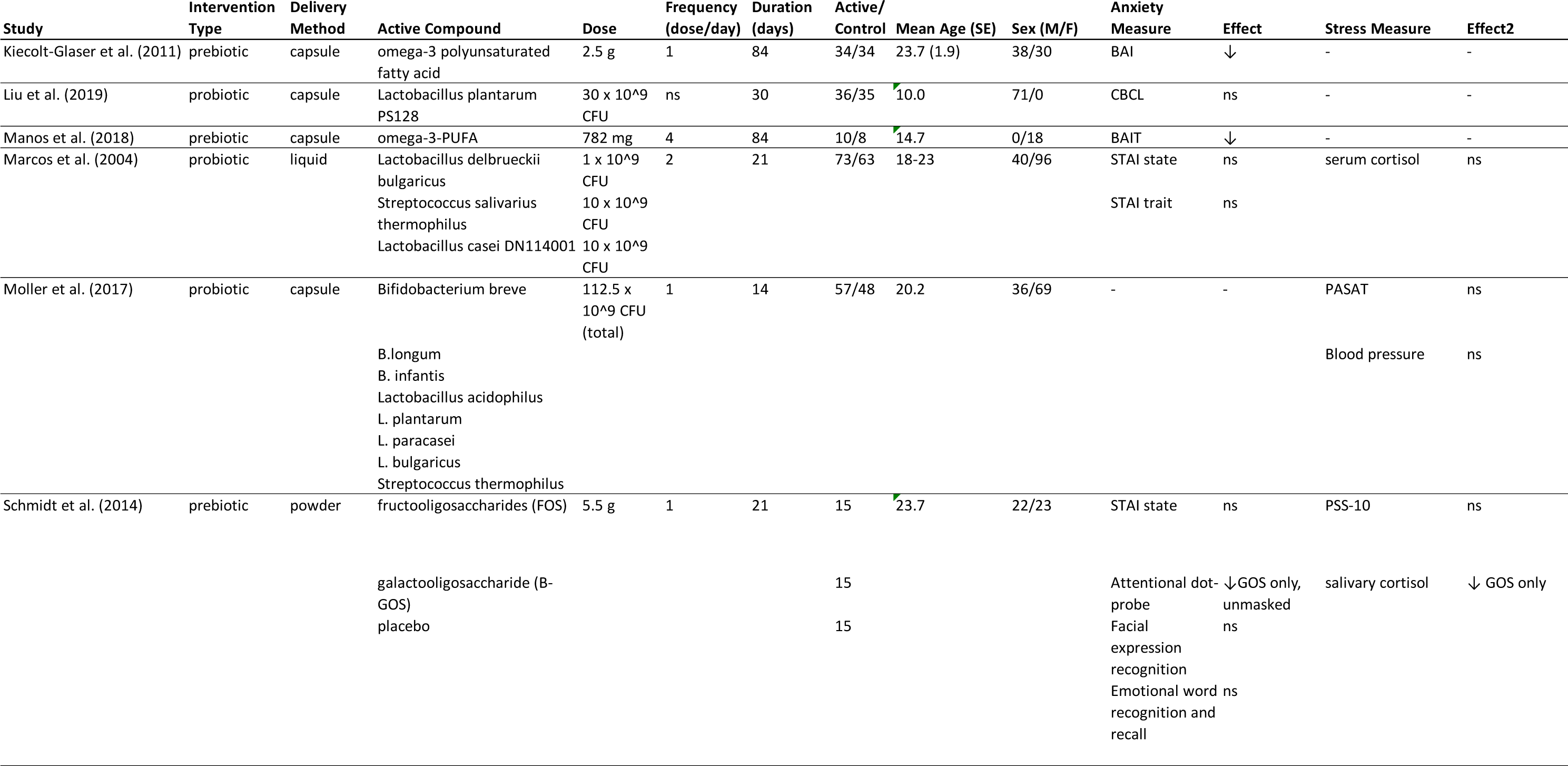
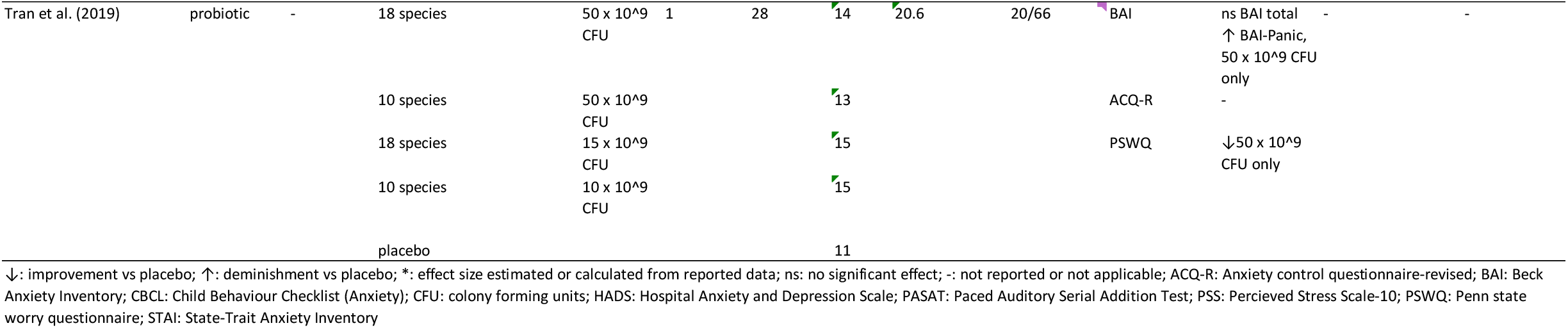
Characteristics of the studies included in the systematic review

**Figure 3:**
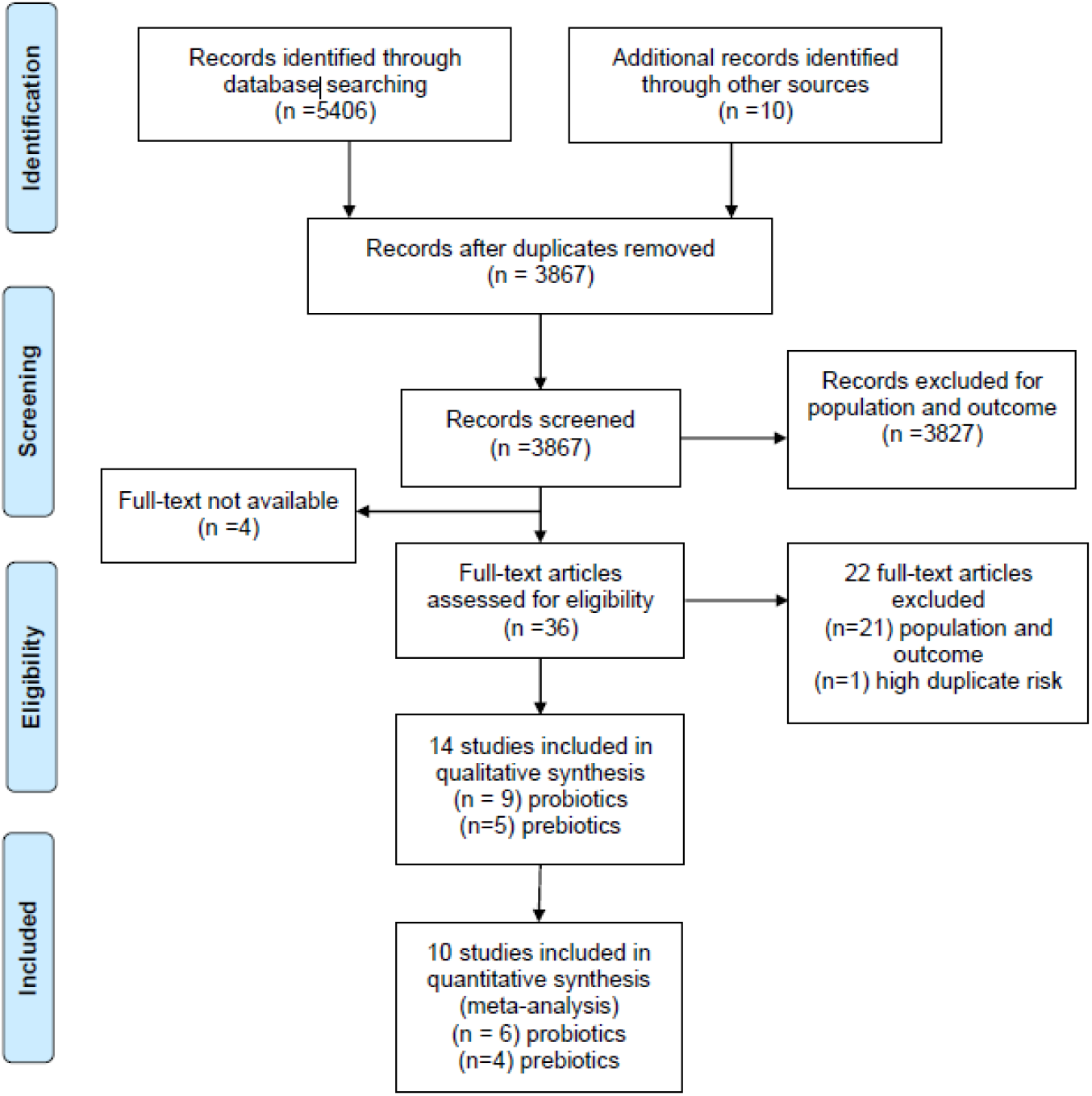
PRISMA flowchart of search results at each step of the systematic review.

The studies we reviewed were quite heterogeneous in the design of the psychobiotic interventions. In the case of probiotics, many different species were used – sometimes alone and sometimes in combination. Of the 9 probiotic studies, 4 used multiple species of probiotics in the same treatment group and one used up to 18 different species in a group (Tran 2019, Tran, Zhebrak et al. 2019). Across the review, species used included *L. plantarum, helveticus, rhamnosus, casei, casei* Shirota, *paracasei, plantarum, bulgaricus, delbrueckii bulgaricus, acidophilus; B. longum, bifidum, breve, infantis; Saccharomyces boulardii; Streptococcus salivarius thermophilus;* and *Streptococcus thermophilus*. Dose size and frequency and duration of intervention also varied widely. While often on the order of tens of billions of colony-forming units (CFU), doses ranged from 10^9 CFU to 10^11 CFU, once to twice a day from 14 to 56 days. Similarly, a number of different prebiotic compounds were employed: galactooligosaccharide, fructooligosaccharide, omega-3 polyunsaturated fatty acid, and fermented ginseng. Doses of prebiotics were more comparable, ranging from 1.8 g/day to 5.5 g/day for from 8 to 56 days, however study duration was also quite variable from 8 to 56 days. Furthermore, there was a large degree of outcome heterogeneity. When anxiety was measured in the studies, it was with a number of validated instruments such as the Beck Anxiety Inventory (BAI), State-Trait Anxiety Inventory (STAI), Hospital Anxiety and Depression Scale (HADS), or Penn State Worry Questionnaire (PSWQ). Moreover, a variety of stress measures were used, including salivary cortisol, immunoglobulin A, metanephrine, alpha-amylase; self-reported stress; blood pressure; pulse rate; serum cortisol; urinary 8-hydroxydeoxyguanosine; and performance on a behavioural task. Taken together, this heterogeneity is likely due to the novelty of the field, however it makes direct comparison between studies more challenging. Future studies should focus on systematic replications of the findings reported in these studies and, where applicable, confirmation of effective and tolerable doses needs to be assessed.

Perhaps in part because of heterogeneity in methods employed, the review revealed mixed results for the reduction of stress using probiotics in adolescents. Of 6 probiotic studies which measured anxiety, 5 failed to find a significant effect. The remaining study (Tran, Zhebrak et al. 2019) reported both an improvement in PSWQ and paradoxical worsening on the BAI-Panic subscale. Caution is warranted with this study however, since errors in the statistical analysis in the initial manuscript warranted a subsequent correction which significantly altered the initially promising reported outcome (Tran 2019). By far the most common study design for both pre- and probiotics involved following a cohort of selected and unselected university students before, during, and after final exams (Marcos, Warnberg et al. 2004, Andersson, Tullberg et al. 2016, Culpepper, Christman et al. 2016, Kato-Kataoka, Nishida et al. 2016, Kato-Kataoka, Nishida et al. 2016, Takada, Nishida et al. 2016, Karbownik, Kreczynska et al. 2020). These studies often only used stress or anxiety as a secondary outcome, focussing on stress-related GI problems or immune performance, mostly with salivary cortisol as the main outcome. Here, there was some evidence (this assessment is based on the statistical analysis) to support *L. casei Shirota* having protective effects on salivary cortisol and self-reported stress in students approaching final exams (Kato-Kataoka, Nishida et al. 2016, Takada, Nishida et al. 2016). A further study from another group reported reduced stress in sleep deprived students after *B. bifidum* administration during exam periods (Culpepper, Christman et al. 2016). About 50% of studies which measured stress in some way found non-significant effects of their probiotic intervention on stress. One study (Karbownik, Kreczynska et al. 2020) even reported increased pulse rate in the probiotic group (*Saccharomyces boulardii*) which could be interpreted as increased physiological stress, although there was no change in self-reported anxiety.

Taken together, the literature revealed that probiotics used had mixed results reducing stress in youth. This makes sense given the large variety of strains available and tested. Furthermore, the literature currently does not support their use in reducing anxiety, and two studies reported adverse effects, e.g. increased BAI (Tran, Zhebrak et al. 2019) and increased pulse rate (Karbownik, Kreczynska et al. 2020).

### Prebiotic interventions

Potentially more promising, but less investigated are prebiotic interventions. Of the five studies that met our inclusion criteria, one using galactooligosaccharide (GOS; (Hughes, Davoodi-Semiromi et al. 2011) found no significant effect on participant stress. Another study of girls with *anorexia nervosa* (Manos et al., 2018) reported no significant effect of omega-3 polyunsaturated fatty acid (PUFA)^2^ on anxiety as measured by the Beck Anxiety Inventory-trait (BAIT) - however this may have been due to lack of statistical power (n = 18).A larger study also used PUFA as the active ingredient (n = 68; (Kiecolt-Glaser, Belury et al. 2011) but reported reduced BAIT in otherwise healthy university students. Another small study (n = 16) investigating the effects of fermented ginseng (Kitaoka, Uchida et al. 2009) reported reduced total STAI score in healthy university students. Finally, one study using GOS (Schmidt, Cowen et al. 2015) reported both reduced attention to negatively valenced stimuli (hypervigilance to negative stimuli is associated with anxiety and depression) and decreased salivary cortisol (although STAI was unaffected). Furthermore, unpublished work by our lab replicates this vigilance reducing effect of GOS in adolescent females (Johnstone, Milesi et al. under review), along with a reduction in self-reported trait anxiety levels. Interestingly, in this study, a reduction in self-reported anxiety levels (STAIT) was only found in participants with high trait anxiety, which suggests that prebiotic interventions may be most effective in cases where there is already some evidence of microbiome dysbiosis.

### Meta-analysis of anxiety outcomes

Of the 14 studies used for the systematic review, 10 were included in the statistical summary (the remaining studies did not assess anxiety but stress only) which was performed with Review Manager 5 (Collaboration 2014). For each study, the standard mean difference (SMD) between the active and control groups was calculated for continuous anxiety outcomes and a Foster plot created (**Figure 4**). Combination of the SMDs revealed a pooled effect size of –0.04 (95% CI: −0.21, 0.14), pointing towards absence of any intervention effect. However, given that the singular SMDs differed substantially amongst each other, ranging from outcomes showing a consistent between-groups anxiety increase (Kitaoka, Uchida et al. 2009, Manos, Bravender et al. 2018), as well as a significant decrease (Kiecolt-Glaser, Belury et al. 2011), we also performed a sensitivity analysis based on study quality (please see **Appendix 3** and **Appendix 4** for the complete risk of bias assessment tables) in order to assess whether any other additional variables other than intervention variable might have biased study results. We found that when at high risk of bias studies were excluded, the effect size increased, reaching a value of –0.16 (95% CI: −0.39, 0.06) (**Figure 5**). It is worth noting that the *only* study at low risk of bias was that reporting the highest effect size: −0.61 (95% CI: −1.09, −0.12), (Kiecolt-Glaser et al., 2011), whereas the two studies reporting an increase in anxiety (Kitaoka, Uchida et al. 2009, Manos, Bravender et al. 2018) were amongst those at highest risk.

**Figure 4:**
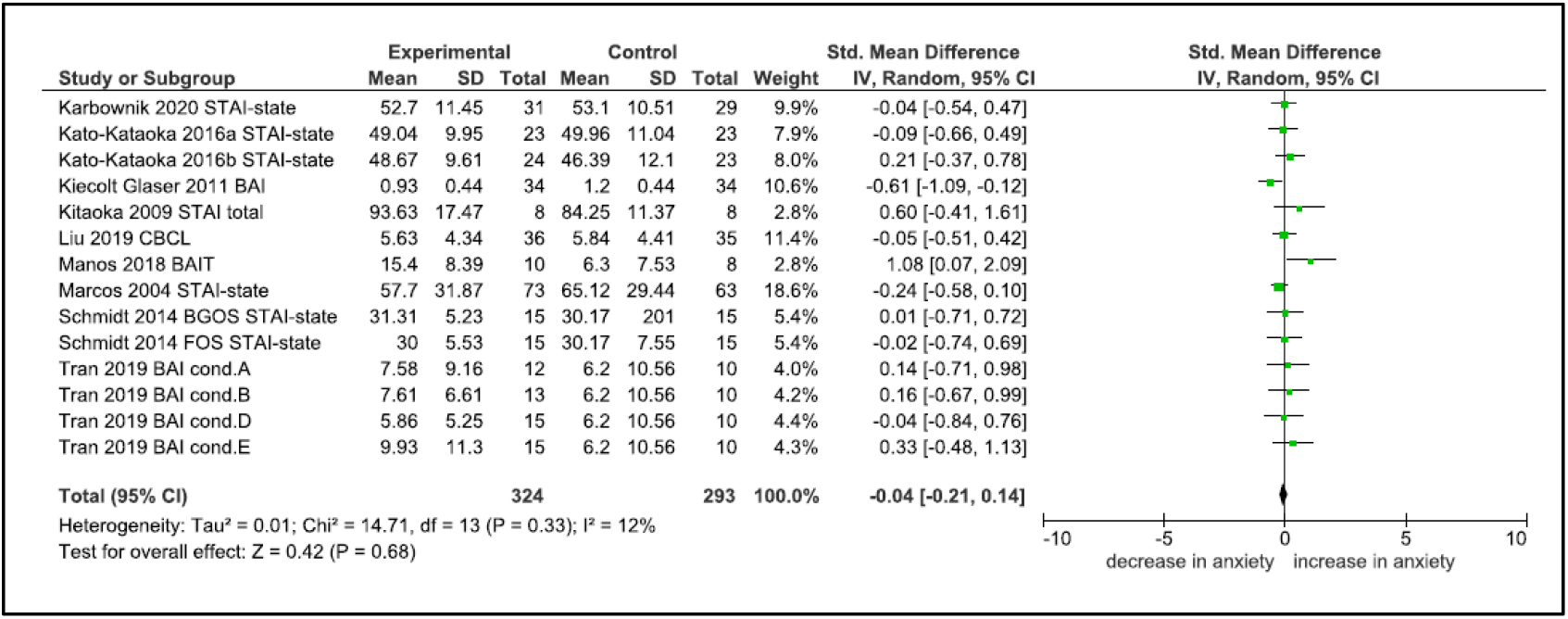
Foster plot of the studies investigating the effect of psychobiotics on anxiety measures.

**Figure 5:**
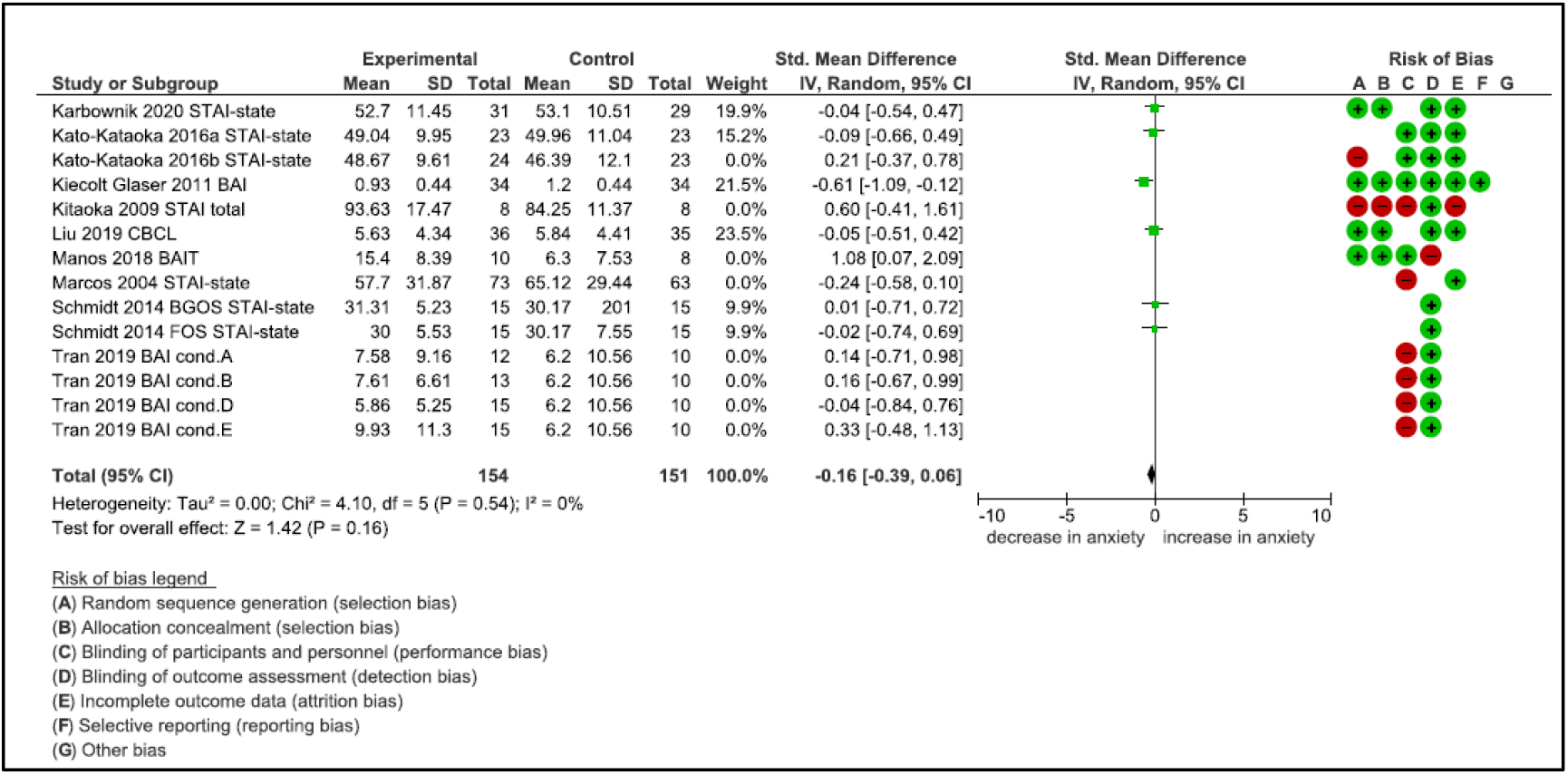
Foster plot excluding the studies at high risk of bias. Reasons for high risk are as follows: **A**. Marcos et al. (2004): concerns in regards to the randomization and allocation sequence and non-blinded design; **B**. Kitaoka et al. (2009): unclear anxiety score differences at baseline between the active and control group, absence of a participants flow diagram and of any relevant information about intervention adherence and missing data; **C**. Manos et al. (2018): unclear anxiety score differences at baseline between active (severe anxiety levels) and control (moderate anxiety levels) groups, not specified reasons for no intervention adherence and missing data, no measurement of state anxiety; **D**. Kato-Kataoka et al. (2016b): no randomized allocation, significantly different anxiety scores at baseline (p<0.05) between the active and control group; **E**. Tran et al. (2019): concerns in regard with randomization and allocation process, not enough information about adherence to the intervention and missing data, concerns about the performed statistical analyses.

### Going forward: recommendations for future research

Based on our systematic review, we conclude that there is currently *limited evidence for use of psychobiotics to treat stress and anxiety in youth*. As mentioned above, strongest effects may be found in persons with high anxious traits (Johnstone, Milesi et al. under review) and therefore, whereas the vast majority of studies in this review used unselected samples, future studies should involve clinical and borderline clinically anxious populations. There is also a need for more high-quality studies which use validated anxiety instruments and behavioural tasks. In particular, it would be important to differentiate whether the intervention aims to improve state and/or trait anxiety, and whether different intervention schedules are required for changing state anxiety as opposed to trait anxiety, for example. Critically, all future studies should include stool sample collections for microbial sequencing to assess direct impacts of intervention on the gut microbiome.

The use of psychobiotics to treat anxiety is a research field still very much in its infancy and given the animal literature and some encouraging preliminary findings in humans (albeit not specifically within the age range chosen for this report), combined with the importance of vulnerability to stress in the adolescent developmental window, we conclude that further research could yield inexpensive, safe and effective means to better manage anxiety. This view is also echoed by young people with lived experience, who not only reported anecdotal evidence of successful psychobiotic interventions, but who also expressed strong interest in contributing to this research drive.

#### Young people with lived experience request clearer instructions

Based on the literature, it would appear that specifically, consumption of *L. casei Shirtoa, L. rhamnosus, L. helveticus*, and *Bifidobacterium* (e.g. *B. infantis*) may provide some protection from the anxiogenic effects of environmental stress, and further long-term studies examining whether the development of anxious traits could be partially ameliorated by these probiotics. The young people noted that such information and guidance on specific bacterial strains would be welcome as the vast number of commercially available probiotic combinations are very confusing. As Becky put it:

> *“When I go into Boots to buy probiotics, there are millions of products available and the prices vary hugely, so I simply don’t know what to choose and I end up getting nothing. It would really help to receive some clarifications on what actually works”*.

As evident from the review above and the meta-analysis, prebiotics hold particular promise as they are non-digestible (fibre) and thus, unlike some probiotics, reliably arrive fully in the gut. Best bets for further study are GOS and fructooligosaccharides which stimulate growth of beneficial *Lactobacillus* and *Bifidobacterium* species. These compounds are also more stable and not subject to the same degradation in potency over time that can be problematic in probiotics. Prebiotics such as FOS make up part of the dietary fibre in naturally occurring foods such as dairy, or are now widely available in concentrated form as food additives or supplements (Lamsal, 2012), which start from around £10 for a month supply.

#### Psychobiotic interventions could benefit from existing cultural practises

One advantage of psychobiotics in the treatment and prevention of youth anxiety that may have been as yet overlooked is the existing cross-cultural prevalence of fermented food sources containing psychobiotics. For instance, anxiolytic strains of *lactobacilli* have been found in traditional fermented doughs from the Congo and Burkina Faso (Abriouel, Ben Omar et al. 2006), Japanese fermented fish (Komatsuzaki, Shima et al. 2005), dairy and pickles from China and Mongolia (Bao, Song et al. 2016), and eastern European *kefir* fermented milk (Simova, Beshkova et al. 2002). This means that anxiolytic psychobiotics consumption could be ripe for a behavioural ‘nudge’, given that these products exist widely in many cultures and therefore potentially present fewer cultural barriers to use.

#### Young people would like to understand ‘the bigger picture’

Young people with lived experience showed much enthusiasm and support for more research in the area of dietary interventions, especially in light of the current gap for the age range of 14-24 years. However, they also asked for research approaches that would look at the bigger picture, which could include information on an individual’s personal circumstances and living situation. What was specifically discouraged was yet another prescriptive approach that would require a change in diet, irrespective of other factors such as educational pressure, work environment aspects such as shift work, sleeping patterns, exercise or a number of other, common comorbidities such as depression. Based on our systematic review, we very much agree with the young people and would like to suggest that one possible explanation for the current success of psychobiotic interventions in animal research is that these studies have maximal control over all these different factors through the use of large, longitudinal cohorts and optimal control of environmental factors, whereas human studies either have chosen not to address these potentially confounding variables or been unable to do so.

#### Sample-specific considerations for future research

The young people provided a number of important suggestions with regards to new research going forward. For example, when discussing different confounding factors, they stressed repeatedly that any data collection on nutritional intake would need to proceed with caution. Specifically, it was warned that providing participants with numerical feedback, such as total calories consumed per day, would easily trigger attempts to control these numbers, as any potential mechanism of control would be latched upon, with potentially far reaching consequences, such as the risk of developing an eating disorder. The young people conceded that while it would be important to obtain comprehensive data on nutritional intake and dietary habits, it would be more helpful to withhold specific detailed feedback and only provide general pointers about nutritional health for example. Last, the young people stated that providing faeces for microbiome sequencing could be a real obstacle for this kind of research, but that they would consider participating if there were a number of options available for stool sampling (i.e. at home or at a testing centre), and that discretion (e.g. packaging for the stool sample that would be both leak and smell-proof, as well as neutrally designed) was paramount. These are all very valuable points as both nutritional analyses, as well as microbiome sequencing are both cornerstones of dietary intervention research into the microbiome gut-brain axis.

#### Towards a new, multidisciplinary research approach

Going forward, it is our view that in order to future-proof this new research area and to allow for sustained scientific progress and breakthrough, what is now needed are systematic, multidisciplinary approaches that consider not only the effect of the dietary intervention on composition of the microbiome, but also on how interventions interact with ongoing brain maturation and functional responsiveness. Similarly, and in line with the important points highlighted by the young people in our consultation, other factors such as hormonal changes due to the specific puberty stage or menstrual cycle, sleep hygiene and patterns, as well as life-style factors such as nutrition and exercise will need to be included to obtain a comprehensive, ‘bigger picture’. Such an approach would also increase uptake and compliance with any future interventions.

Future studies could adopt a research approach that is already practised in the field of developmental cognitive neuroscience (DCN) (Johnstone and Cohen Kadosh in press), which focuses on investigating how the complex interplay of genetic, environmental and brain maturational factors shape psychological functioning in development to improve outcomes for the individual (Johnson, Halit et al. 2002, Johnson, Grossmann et al. 2009, Cohen Kadosh 2011). Moreover, placed at the intersection of nature vs. nurture, the DCN research approach always assumes a multi-level and multi-factor approach to understanding change which, by definition is multidisciplinary. Given that the field of microbiome and gut-brain axis research is still emerging and finding its shape, we would like to stress that any real progress will depend on the adoption of a similarly comprehensive multifactorial and multidisciplinary research approach for pinpointing mechanisms and translation in both animal and human models. Therefore, priority for funding should be given to projects that bring together the expertise and the collaboration of scientists from a range of fields (education, psychology, microbiology, neuroscience, and nutrition) and, importantly, guidance and advice from young people with lived experience to ensure that all research continues to address the questions that are relevant to the lives of young people with anxiety.

To conclude, the gut microbiome, and its effect on behaviour and mental health has captured the interest and imagination of scientists and the wider public alike. However, as a still relatively unexplored area, any real progress will require a multidisciplinary research approach, which gives priority to specifying mechanisms in the human and animal models, providing causal understanding and addressing realistic outcomes. This is particularly critical in light of strong public and commercial interests that are presently outpacing research efforts. Encouragingly, this approach has also been met with much enthusiasm from young people with lived experience of mental health problems, which suggests that these research approach and interventions may be ripe for a behavioural ‘nudge’ and potentially present few cultural barriers to use.

## Data Availability

Data are available from corresponding author on reasonable request.

## Appendix 1: methods

### Eligibility criteria

#### Participants

Only studies whose participants mean age fell within the age range of 10-24 years old were included, regardless of gender, religious, ethnicity and socio-demographic status. Both healthy and clinical samples were eligible, as far as either anxiety or stress were measured.

#### Study and intervention type

Only controlled trials that had assessed anxiety and stress as primary or secondary outcome were searched. At least two measures-one pre- and one post-intervention, needed to have been collected. No selection criteria were considered for follow-up and settings. Studies based on either prebiotics or probiotics supplements intake were included, regardless of the form they were administered in-tablets, powders, capsules, fortified food, food, drinks. Studies assessing probiotics and prebiotics-based interventions in combination between with each other (i.e. synbiotics) were excluded.

#### Comparators

Both placebo and treatment as usual comparison groups were eligible.

#### Outcomes

##### Primary outcome

Overall anxiety symptoms improvement as assessed by the included studies. Validated-self reported anxiety measures and questionnaires as well as structured and semi-structured interviews were eligible.

##### Secondary outcome

Behavioural (as assessed by e.g. emotional stroop, dot-probe, speech test) as well as stress measures were eligible.

#### Report criteria

Studies fulfilling the following criteria were included: data did not necessarily have to be available, but needed to be published and peer-reviewed. No restrictions were applied for date of publication.

##### Search strategy

7 databases were searched (PubMed, Embase, Cochrane, Scopus, Ovid, Orbis, Web of Science) and the search strategies reported in the Appendix 2 were implemented. In addition, hand-searching of references cited in each publication as well as the bibliography of relevant systematic review articles was performed.

#### Study records

##### Study selection and data extraction

The search output (5416 studies) was imported into EppiReviewer4(EPPI-Centre 2020) and accurately controlled for duplicates. After 1549 duplicates had been removed, 3867 studies were double-screed (double screening performed by M. Basso and P. Knytl in consultation with K. Cohen Kadosh when disagreement occurred) for titles and abstract-among them, 3827 were excluded for population and outcome, whereas 4 due to text unavailability. 36 full-text studies were then double-assessed for eligibility-21 were excluded for population and outcome while 1 was excluded as highly likely to have assessed the same sample cohorts of other two included studies (Kato-Kataoka, Nishida, Takada, Kawai, et al., 2016; Kato-Kataoka, Nishida, Takada, Suda, et al., 2016). The final output for the systematic review was 14 studies, among which 10 were included in the meta-analyses. Excel tables were used to support the qualitative data extraction process (performed by M.Basso an P.Knytl), whereas Review Manager 5 was used to extract the quantitative data. When the research report did not provide all the data needed, authors were contacted via e-mail.

##### Risk of bias assessment

In regard with anxiety studies, the risk of bias (RoB) assessment was done at outcome level, whereas the RoB of studies measuring stress was assessed at study level. Based on the Revised Cochrane risk-of-bias tool for randomization trials (RoB-2),(Sterne, Savovic et al. 2019) the following bias domains were considered:

- Random sequence generation (selection bias)
- Allocation concealment (selection bias)
- Personnel and participant blinding (performance bias)
- Outcome assessment blinding (detection bias)
- Incomplete outcome data (attrition bias)
- Selective reporting (reporting bias)
- Other sources of bias

##### Statistical analyses

Data were extracted as standardized mean differences (SMDs) with 95% CI and I^2^ statistics was used as a between-studies heterogeneity index (Higgins, Thompson et al. 2003). The pooled SMD was calculated based on a random-effects model using Review Manager 5. Sensitivity analyses were also performed removing the studies at high risk of bias from the analyses.

## Appendix 2: search terms

**PUBMED**: 449 results

#1 humans[MeSH Terms] OR patients[MeSH Terms] OR research subjects[MeSH Terms] OR human experimentation[MeSH Terms] OR human*[title/abstract] OR client*[title/abstract] OR individual*[title/abstract] OR subject*[title/abstract] OR participant* [title/abstract

#2 minors[MeSH Terms] OR adolescent[MeSH Terms] OR child[MeSH Terms] OR adolescence[MeSH Terms] OR adolescent, development[MeSH Terms] OR child, development[MeSH Terms] OR paediatric[MeSH Terms] OR puberty[Mesh Terms] OR young adult[sMeSH Terms] OR teen*[title/abstract] OR young men[title/abstract] OR young women[title/abstract] OR adolescen*[title/abstract] OR youth [title/abstract] OR undergraduate student* [title/abstract] OR college student* [title/abstract]

#3 prebiotics[MeSH Terms] OR probiotics[MeSH Terms] OR dietary carbohydrates[Mesh Terms OR dietary fiber[MeSH Terms] OR psychobiotic*[title/abstract] OR probio*[title/abstract] OR prebio*[title/abstract] OR pro-bio*[title/abstract] OR pre-bio*[title/abstract] OR lactobacill* [title/abstract] OR bifidobacteri*[title/abstract]

#4 anxiety[MeSH Terms] OR anxiety disorders[MeSH Terms] OR test anxiety scale[MeSH Terms] OR manifest anxiety scale[MeSH Terms] OR patient health questionnaire[MeSH Terms] OR survey and questionnaire/psychology[MeSH Terms] OR psychology, child[MeSH Terms] OR psychology, adolescent[MeSH Terms] OR child behavior disorders[MeSH Terms] OR panic[MeSH Terms] OR affect[Mesh Terms] OR affective symptoms[MeSH Terms] OR performance anxiety[MeSH Terms] OR stress, psychological[MeSH Terms] OR anxi*[title/abstract] OR stress[Text Word] OR PSS[Text Word] OR SAS[Text Word] OR DASS[Text Word] OR HADS[Text Word] OR HAD-S[Text Word] OR BAI[Text Word] OR STAI[Text Word] OR PHQ[Text Word] OR Emotional decision*[text word] OR psychological distress [mesh terms] OR emotions[mesh Terms] OR emotion regulation [mesh terms] OR “emotional stroop” [text word] OR “dot probe[text word]

**COCHRANE**: 372 results

#1 [mh humans] OR [mh patients] or [mh “research subjects”] OR [mh “human experimentation”] OR participant OR client OR individual OR subject

#2 [mh minors] OR [mh adolescent] OR [mh adolescence] OR [mh child] or [mh puberty] or [mh “adolescent, development”] or [mh “child, development”] OR [mh paediatric] or “young men” or “young women” or teen* or adolescen* or “undergraduate student” or “college student”

#3 [mh probiotics] or [mh prebiotics] or psychobiotic* or probio* or prebio* or [mh “dietary carbohydrates”] or [mh “dietary fiber”] or bifidobacteri* or lactobacill*

#4 [mh anxiety] or [mh “anxiety disorders”] or [mh affect] or [mh panic] or [mh “affective symptoms”] or [mh “patient health questionnaire”] or [mh “survey and questionnaires”/PX] or [mh “psychology, child”] or [mh “psychology, adolescent”] or [mh “test anxiety scale”] or [mh “manifest anxiety scale”] or [mh “child behavior disorders”] or [mh “performance anxiety”] or [mh “stress, psychological”] or [mh “emotion regulation”] or [mh emotions] or [mh “psychological distress”] or anxi* or sas or dass or hads or had-s or stai or bai or phq or pss or “emotional stroop” or “dot-probe”

**MEDLINE:** 105 results

#1 (MM “Humans”) OR (MH “Patients+”) OR (MH “Research Subjects+”) OR (MH “Human Experimentation+”) OR “human*” OR “subject*” OR “individual* OR “client*” OR participant*

#2 (MM “Minors”) OR (MM “Adolescent”) OR (MM “Adolescent Development”) OR (MH “Child+”) OR (MH “Child Development+”) OR “adolescen*” OR (MH “Pediatrics”) OR (MM “Puberty”) OR (MM “Young Adult”) OR “teen*” OR “youth” OR “young men” OR “young women” or “undergraduate student” or “college student”

#3 (MM “Probiotics”) OR (MM “Prebiotics”) OR (MM “Dietary Carbohydrates”) OR (MM “Dietary Fiber”) OR “psychobiotic*” OR “probio* OR “prebio*” OR “pro-bio*” OR “pre-bio*” OR “lactobacill*” OR “bifidobacteri*”

#4 (MH “Anxiety+”) OR (MH “Anxiety Disorders+”) OR (MM “Test Anxiety Scale”) OR (MM “Manifest Anxiety Scale”) OR (MM “Patient Health Questionnaire”) OR (MM “Surveys and Questionnaires”/PX) OR (MM “Psychology, Child”) OR (MM “Psychology, Adolescent”) OR (MM “Child Behavior Disorders”) OR (MM “Panic”) OR (MH “Affect+”) OR (MH “Affective Symptoms+”) OR (MM “Performance Anxiety”) OR (MH “Stress, Psychological+”) OR (MH “Emotions+”) OR (MH “Psychological Distress”) OR (MM “Emotional Regulation”) OR “SAS” OR “DASS” OR “HADS” OR “HAD-S” OR “BAI” OR “STAI” OR “PHQ” OR “PSS” OR “anxi*” OR “emotional stroop” OR “Emotional decision*” OR “dot probe” OR “stress”

**SCOPUS:** 2071 results

((TITLE-ABS-KEY(human) OR TITLE-ABS-KEY(patient) OR TITLE-ABS-KEY(“research subject”) OR TITLE-ABS-KEY(“human trial”) OR TITLE-ABS-KEY(subject) OR TITLE-ABS-KEY(client) OR TITLE-ABS-KEY(individual))) and ((TITLE-ABS-KEY(minor) OR TITLE-ABS-KEY(adolescen*) OR TITLE-ABS-KEY(child) OR TITLE-ABS-KEY(“young adult”) OR TITLE-ABS-KEY(teen*) OR TITLE-ABS-KEY(“young m?n”) OR TITLE-ABS-KEY(“young wom?n”) OR TITLE-ABS-KEY(youth) OR TITLE-ABS-KEY(“undergraduate student*”) OR TITLE-ABS-KEY(“college student*”))) and ((TITLE-ABS-KEY(prebiotic) OR TITLE-ABS-KEY(probiotic) OR TITLE-ABS-KEY(psychobiotic) OR TITLE-ABS-KEY(lactobacill*) OR TITLE-ABS-KEY(bifidobacter*) OR TITLE-ABS-KEY(“dietary fiber”))) and ((TITLE-ABS-KEY(anxi*) OR TITLE-ABS-KEY(affect*) OR TITLE-ABS-KEY(“emotion regulation”) OR TITLE-ABS-KEY(“emotional disorder”) OR TITLE-ABS-KEY(“patient health questionnaire”) OR TITLE-ABS-KEY(panic) OR TITLE-ABS-KEY(psychologic*) OR TITLE-ABS-KEY(sas) OR TITLE-ABS-KEY(dass) OR TITLE-ABS-KEY(had?s) OR TITLE-ABS-KEY(bai) OR TITLE-ABS-KEY(stai) OR TITLE-ABS-KEY(pss) OR TITLE-ABS-KEY(phq) OR TITLE-ABS-KEY(“emotional stroop”) OR TITLE-ABS-KEY(“dot?probe”) OR TITLE-ABS-KEY(stress) OR TITLE-ABS-KEY(distress)))

**EMBASE:** 347 results

#1 ‘human’/exp OR ‘patient’/exp OR ‘research subject’/exp OR ‘human experiment’/exp OR human*:ti,ab OR client*:ti,ab OR individual*:ti,ab OR subject*:ti,ab OR participant*:ti,ab

#2 ‘minor’/exp OR ‘adolescent’/exp OR ‘child’/exp OR ‘adolescence’/exp OR ‘young adult’/exp OR ‘young m#n’:ti,ab OR ‘young wom#n’:ti,ab OR teen*:ti,ab OR adolescen*:ti,ab OR youth:ti,ab or ‘undergraduate student*’:ti,ab or ‘college student*’:ti,ab

#3 ‘probiotic agent’/exp OR ‘prebiotic agent’/exp OR ‘dietary fiber’/exp OR ‘psychobiotic agent’/exp OR ‘lactobacillus’/exp OR ‘bifidobacterium’/exp OR psychobiotic*:ti,ab OR pro?bio*:ti,ab OR pre?bio*:ti,ab OR lactobacill*:ti,ab OR bifidofacteri*:ti,ab

#4 ‘anxiety’/exp OR ‘anxiety disorder’/exp OR ‘anxiety psychology’/exp OR ‘anxiety assessment’/exp OR ‘child psychology’/exp OR ‘affect’/exp OR ‘emotion regulation’/exp OR ‘emotional disorder’/exp OR ‘emotional stress’/exp OR ‘patient health questionnaire’/exp OR ‘behavior/psychological aspect’ OR sas:ti,ab OR dass:ti,ab OR had?s:ti,ab OR bai:ti,ab OR stai:ti,ab OR phq:ti,ab OR pss:ti,ab OR anxi*:ti,ab OR ‘emotional stroop’:ti,ab OR ‘dot?probe’:ti,ab OR distress:ti,ab OR stress:ti,ab OR ‘emotional decision*’:ti,ab

**PSYCHINFO:** 33 results

#1 DE “Experimental Subjects” OR AB participant* OR TABsubject* OR AB individual* OR AB human*

#2 DE “Adolescent Psychopathology” OR DE “Adolescent Psychology” OR DE “Adolescent Psychiatry” OR DE “Adolescent Development” OR DE “Adolescent Behavior” OR DE “Adolescent Characteristics” OR DE “Adolescent Health” OR DE “Early Adolescence” OR AB child* OR AB adolescen* OR AB teen* OR AB youth OR AB “young adult*” OR AB “young women” OR AB “young men” or AB”undergraduate student*” or AB”college student*”

#3 DE “Dietary Supplements” OR AB probio* OR AB prebio* OR AB lactobacill* OR AB bifidobacteri* OR AB psychobiotic* OR AB “dietary fiber*”

#4 ((((((((((DE “Anxiety” OR DE “Anxiety Sensitivity” OR DE “Computer Anxiety” OR DE “Health Anxiety” OR DE “Mathematics Anxiety” OR DE “Performance Anxiety” OR DE “Social Anxiety” OR DE “Speech Anxiety” OR DE “Test Anxiety” OR DE “Anxiety Disorders” OR DE “Castration Anxiety” OR DE “Death Anxiety” OR DE “Generalized Anxiety Disorder” OR DE “Obsessive Compulsive Disorder” OR DE “Panic Attack” OR DE “Panic Disorder” OR DE “Phobias” OR DE “Separation Anxiety Disorder” OR DE “Trichotillomania” OR DE “Taylor Manifest Anxiety Scale” OR DE “State Trait Anxiety Inventory” OR DE “Childrens Manifest Anxiety Scale”) OR (DE “Child Behavior Checklist”)) OR (DE “Mental Health and Illness Assessment”)) OR (DE “Health Psychology Assessment” OR DE “Health Attitude Measures” OR DE “Health Behavior Measures”))) OR (DE “Psychodiagnostic Measures”)) OR (DE “Psychodiagnostic Interview” OR DE “Diagnostic Interview Schedule” OR DE “Structured Clinical Interview”)) OR (DE “Attentional Bias”)) OR (DE “Behavioral Assessment”)) OR (DE “Distress”) OR AB “emotion regulation” OR TX “emotional stroop” OR TX “dot probe” OR TX SAS OR TX DASS OR TX DA?S OR TX BAI OR TX STAI OR TX PHQ OR TX PSS

**WEB OF SCIENCE:** 2029 results

#1 TOPIC (human* OR patient* OR subject* OR client* OR individual*)

#2 TOPIC (adolescen* OR child* OR “young adult*”OR teen* OR “young m?n” OR “young wom?n” OR youth or “undergraduate student*” or “college student*”)

#3 TOPIC (prebiotic* OR probiotic* OR psychobiotic*OR lactobacill* OR bifidobacter*OR “dietary fiber*”)

#4 TOPIC (anxi* OR affect*OR “emotion* regulation” OR “emotional disorder* OR “patient health questionnaire” OR sas OR dass OR had?s OR stai OR bai OR phq OR pss OR panicOR “emotional stroop”OR “dot?probe” OR “psychological distress” OR stress)

## Appendix 3: Risk of Bias Assessment of anxiety studies

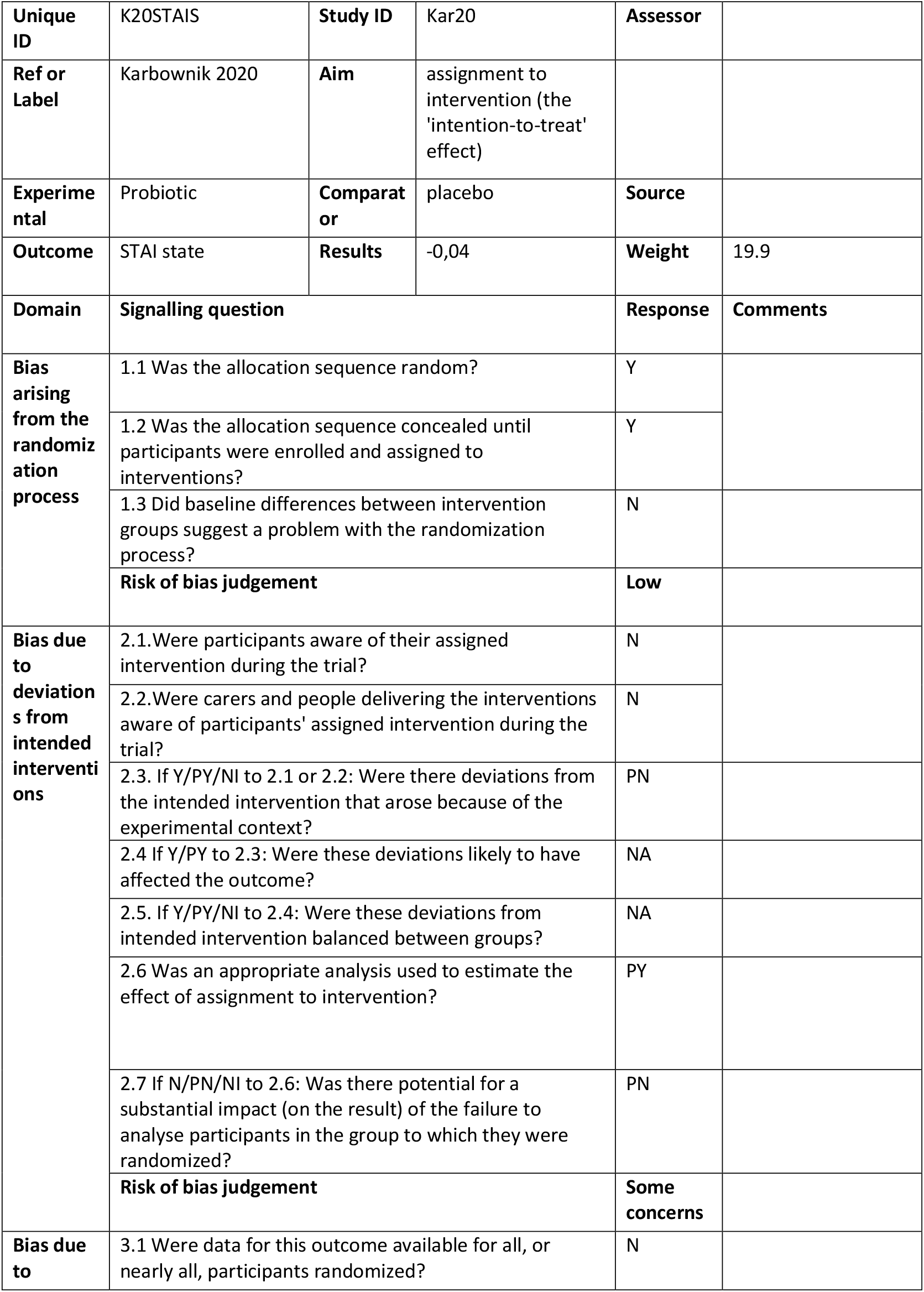
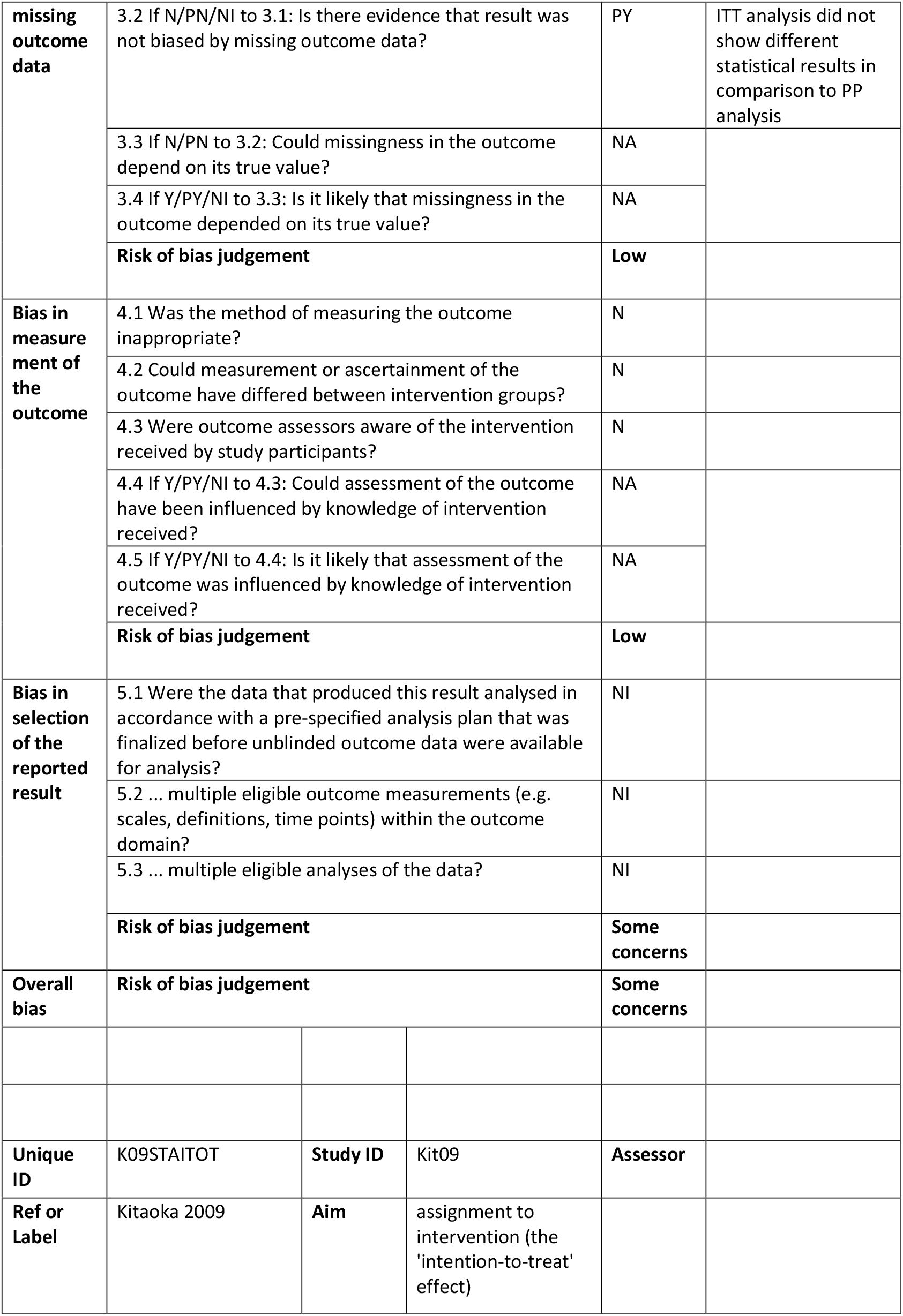
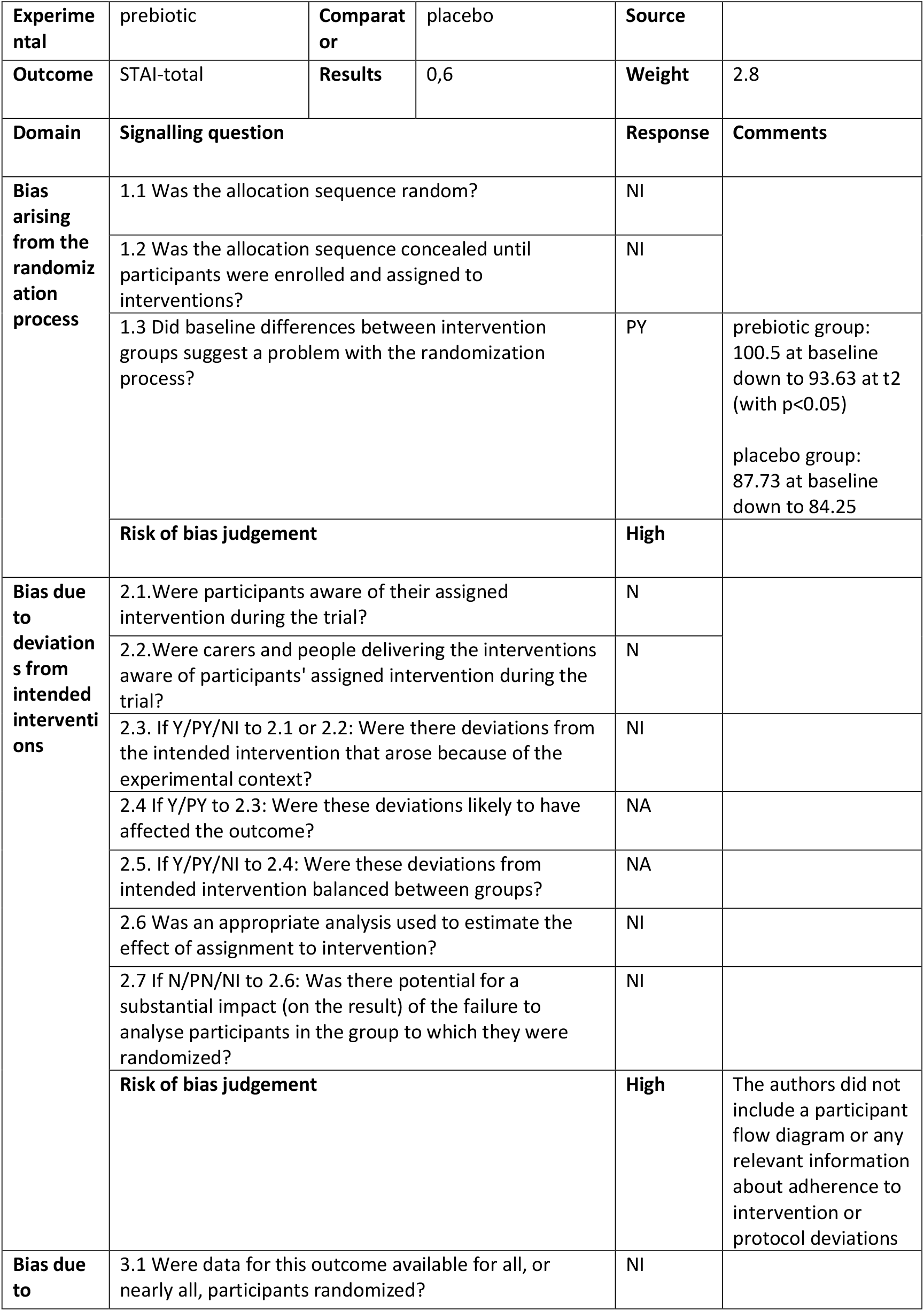
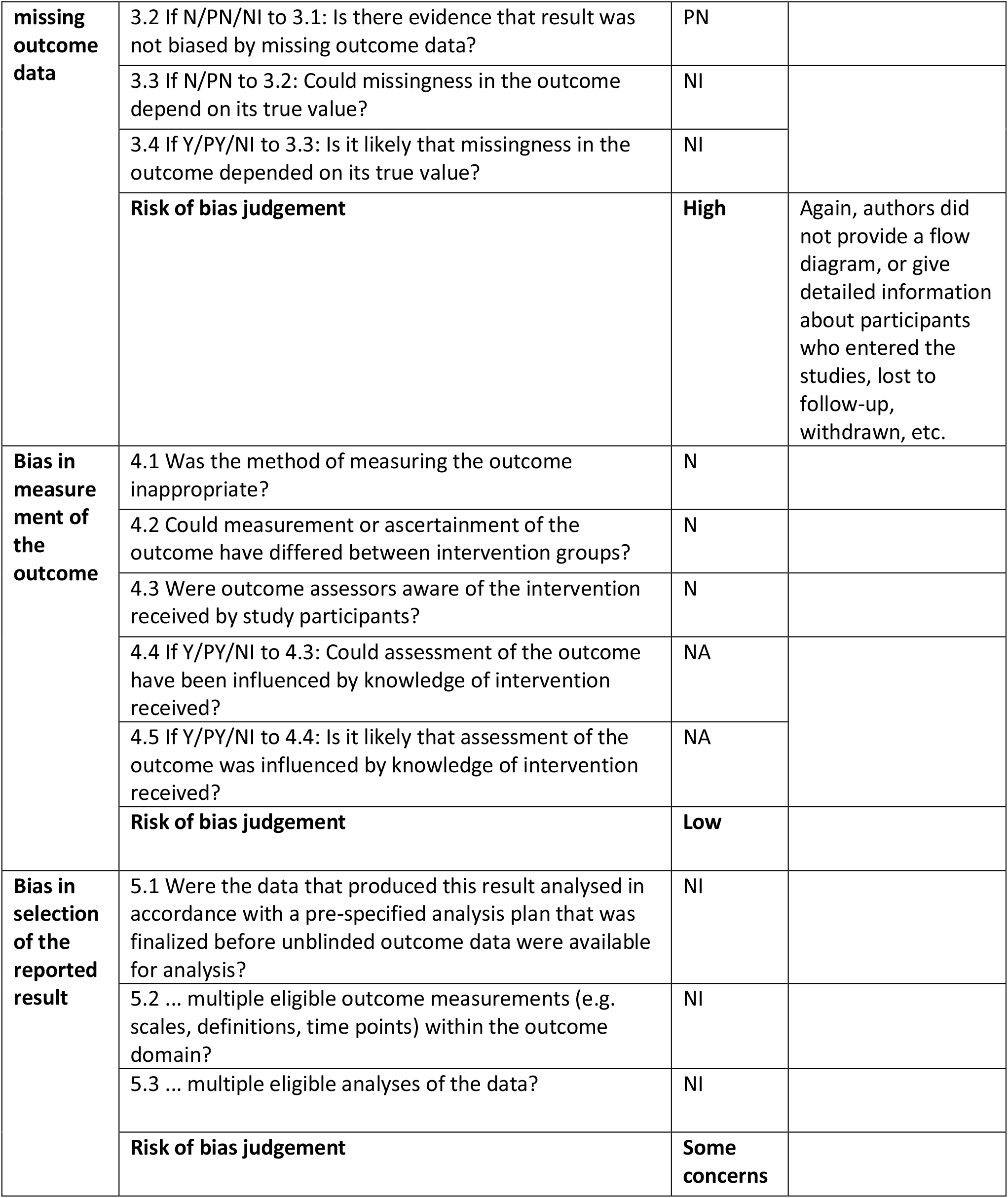
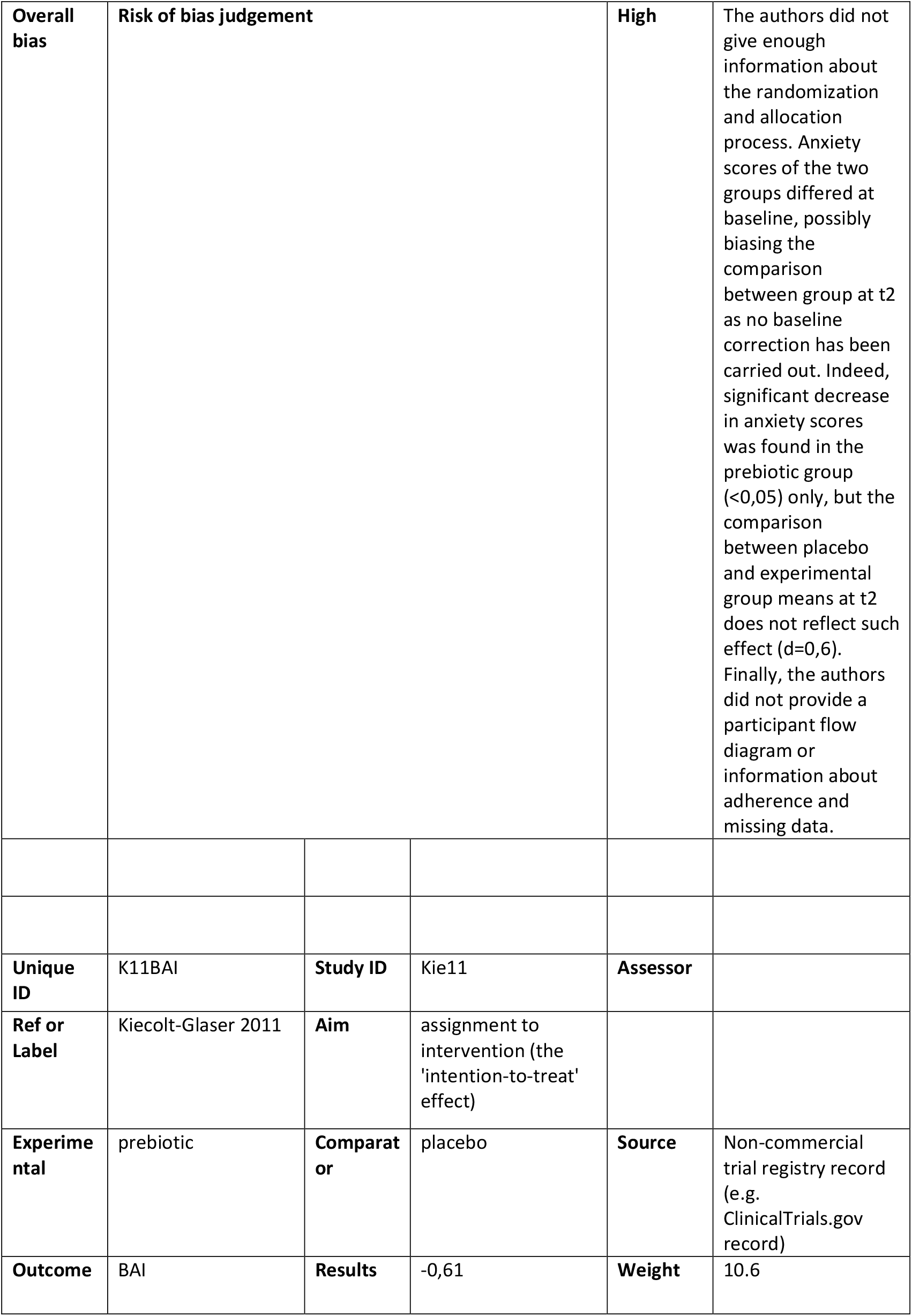
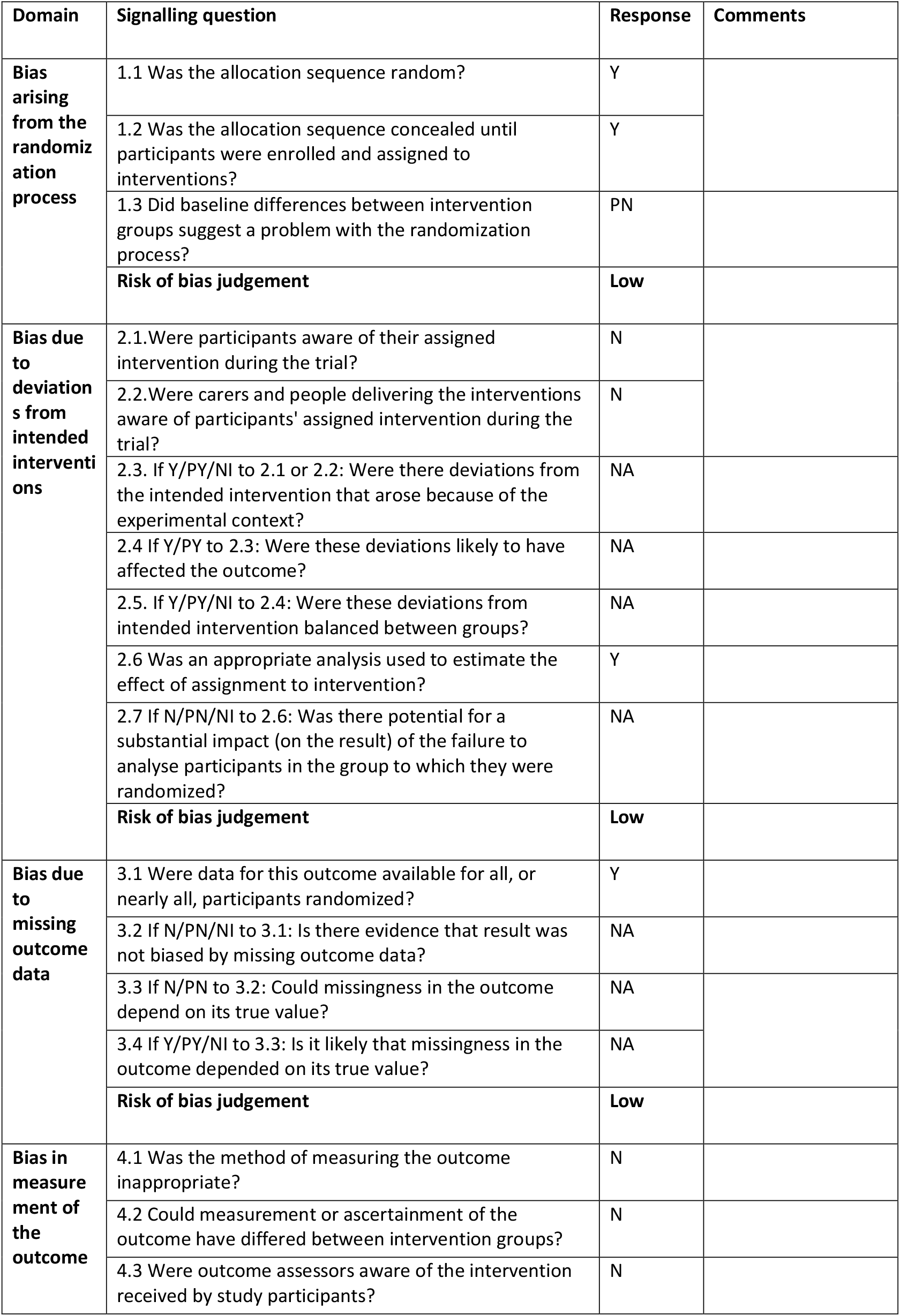
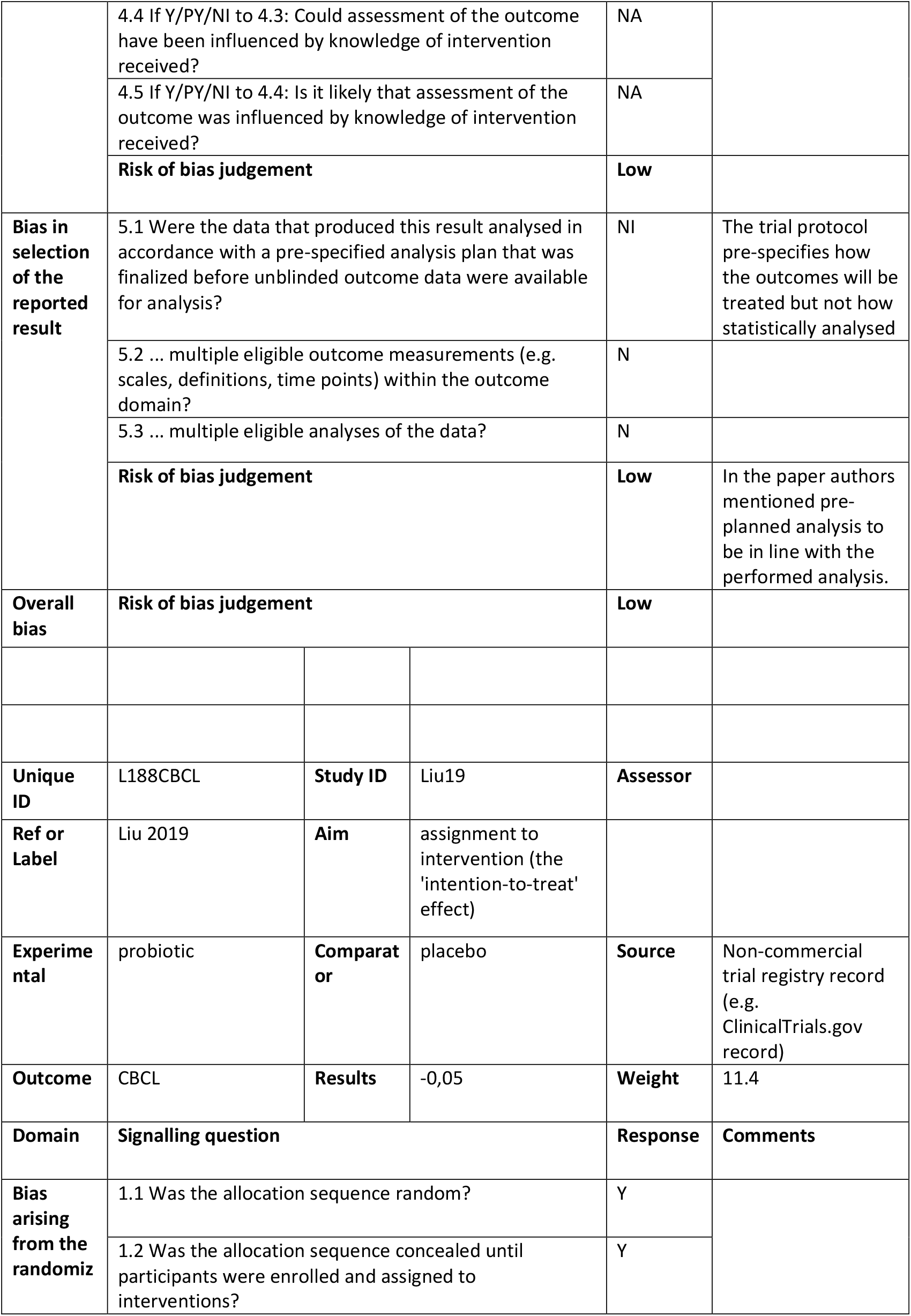
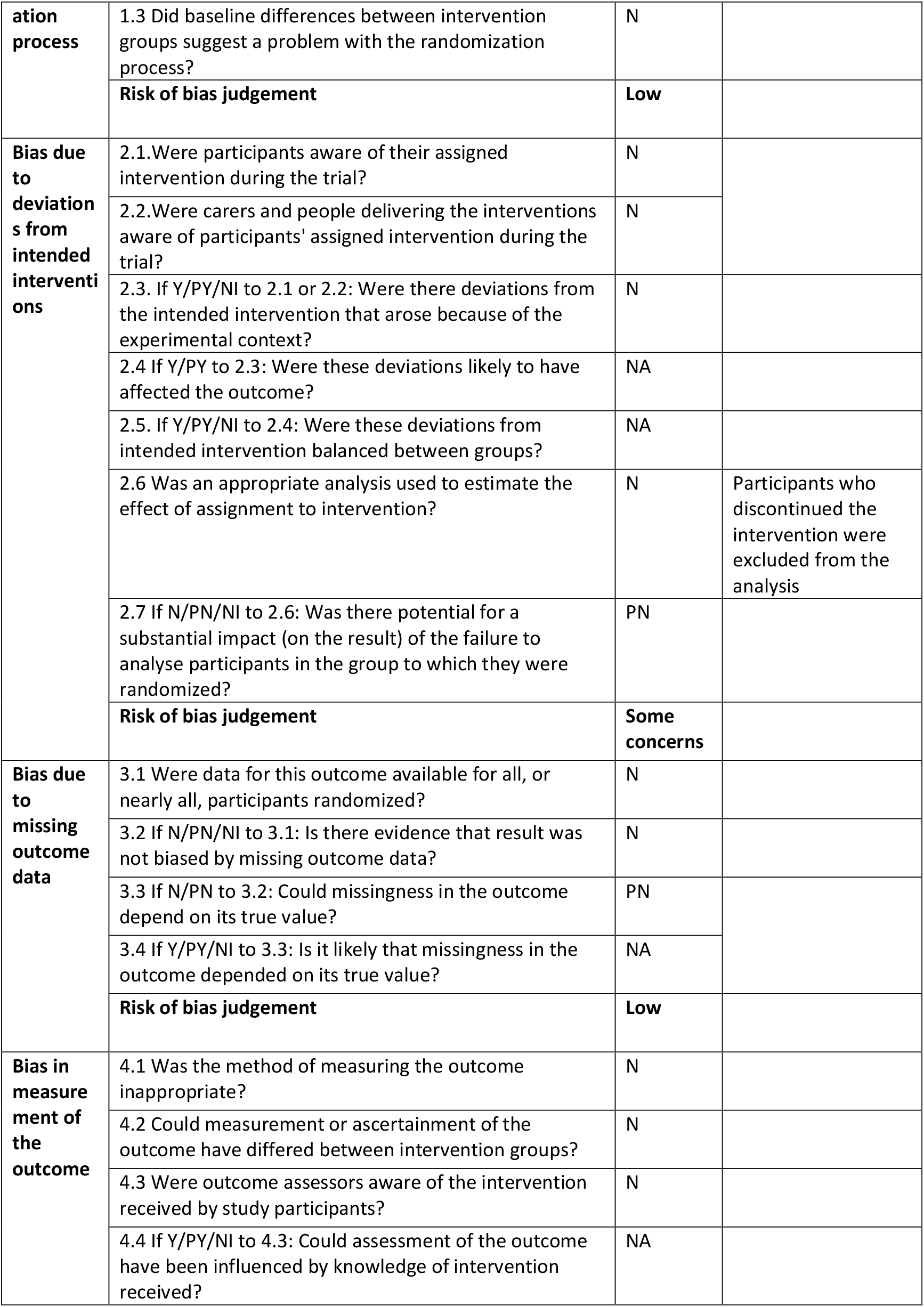
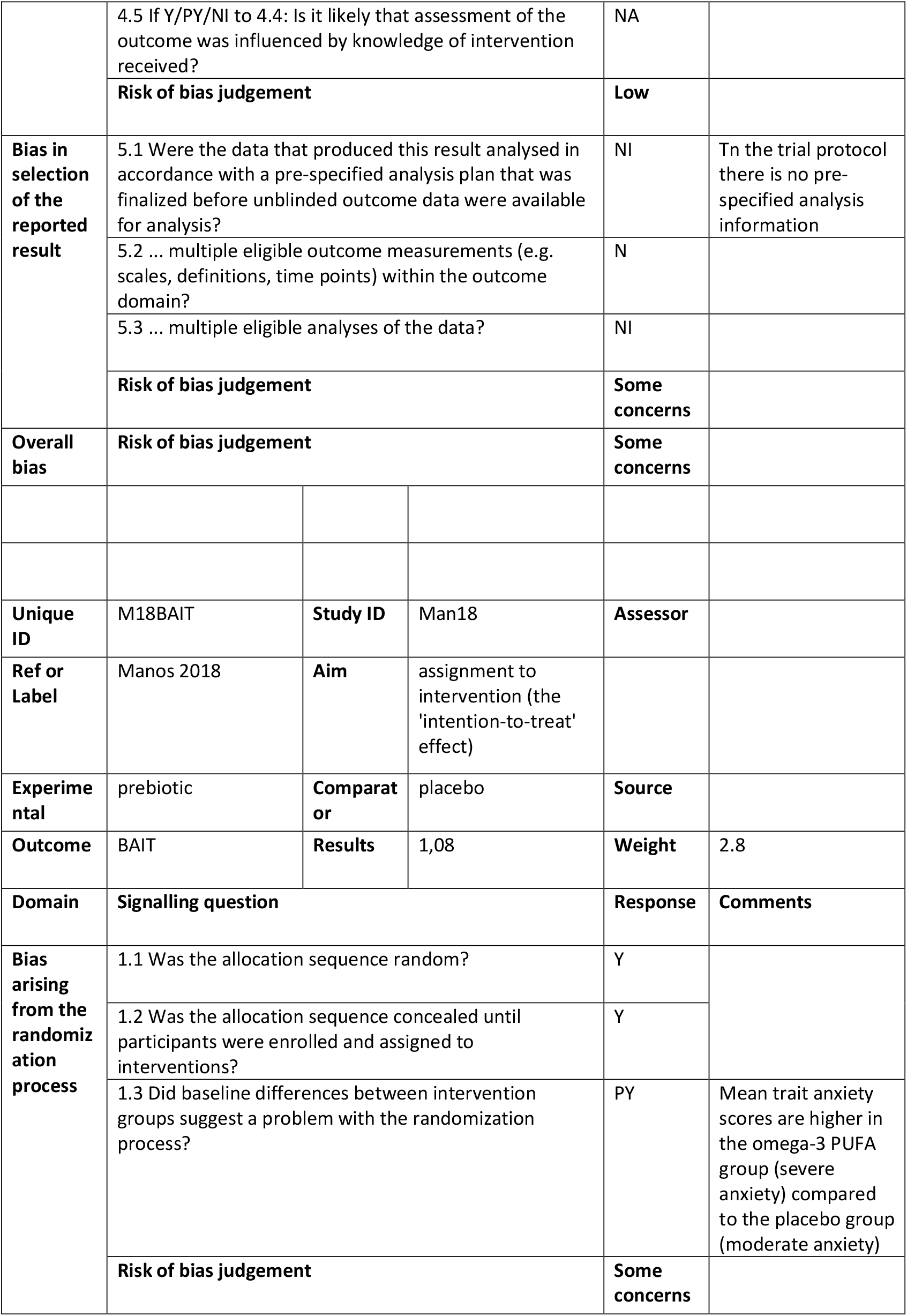
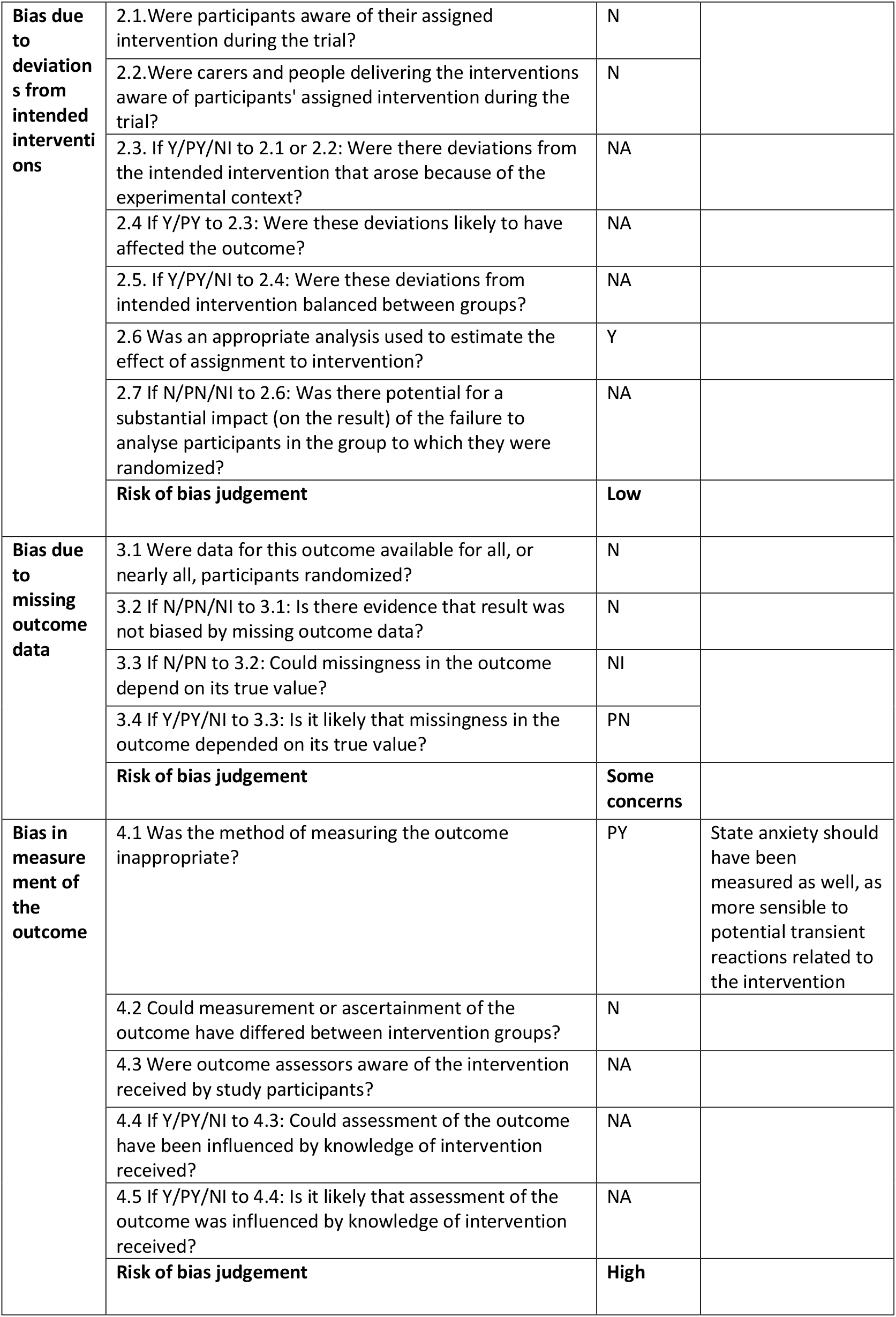
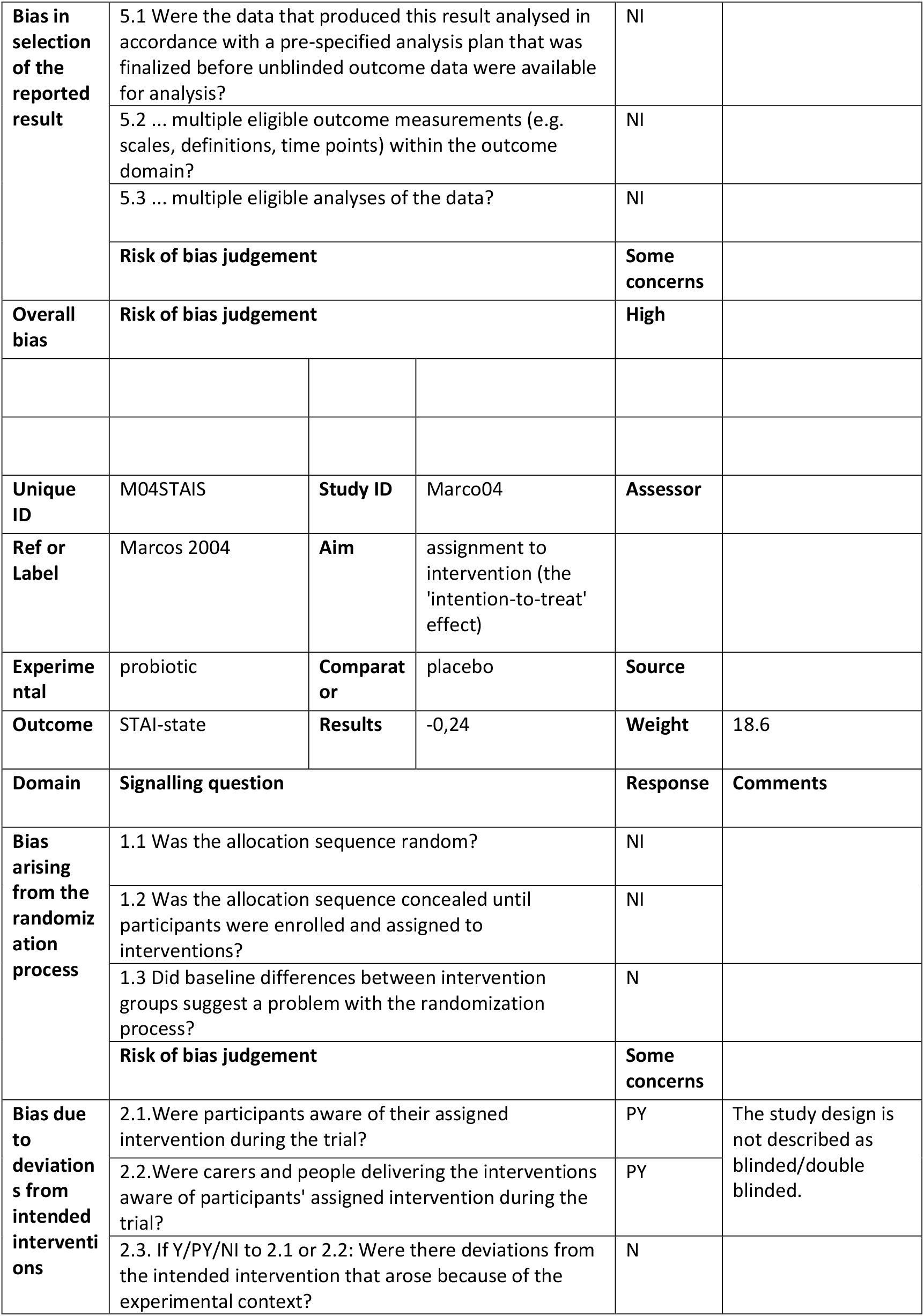
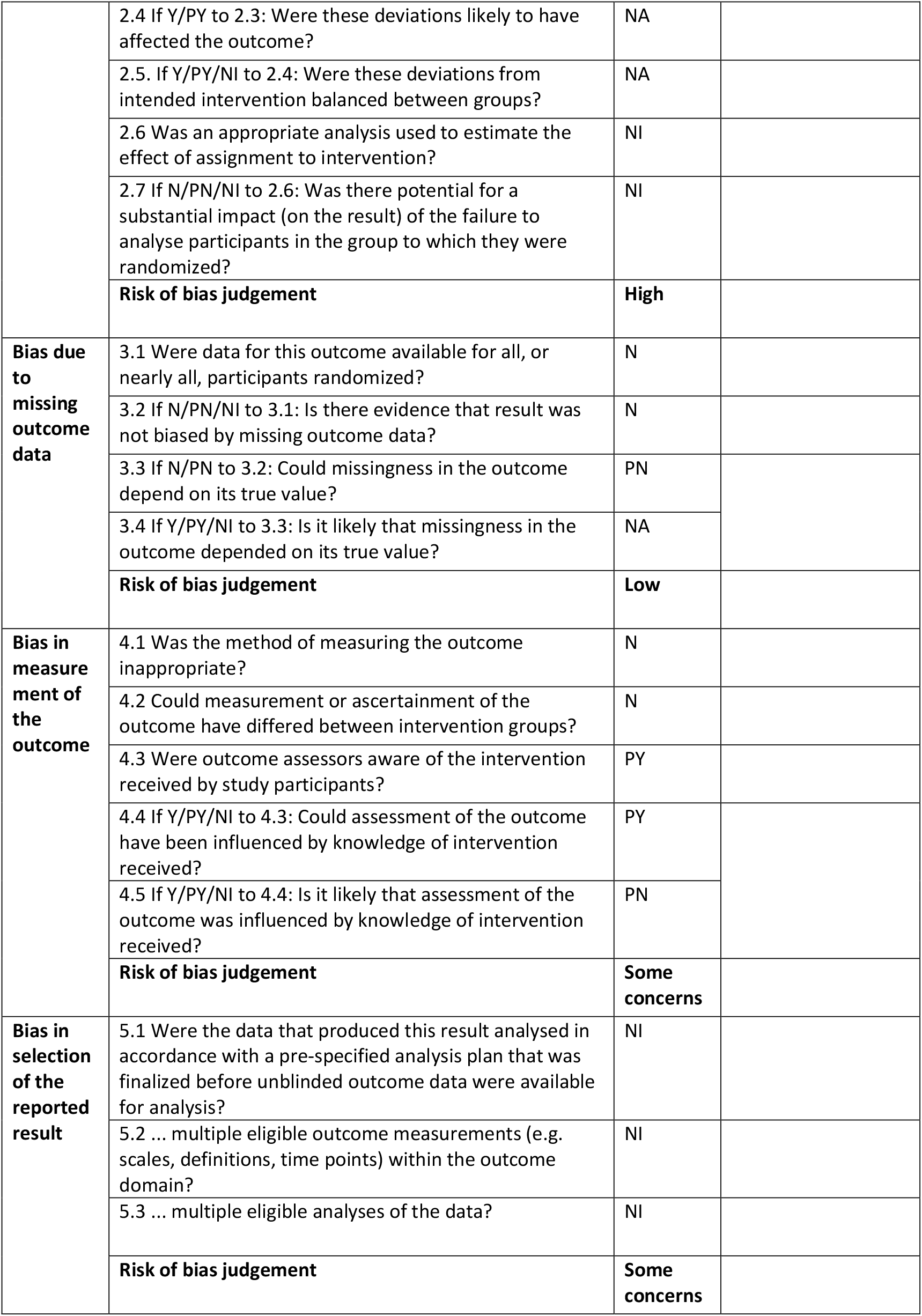
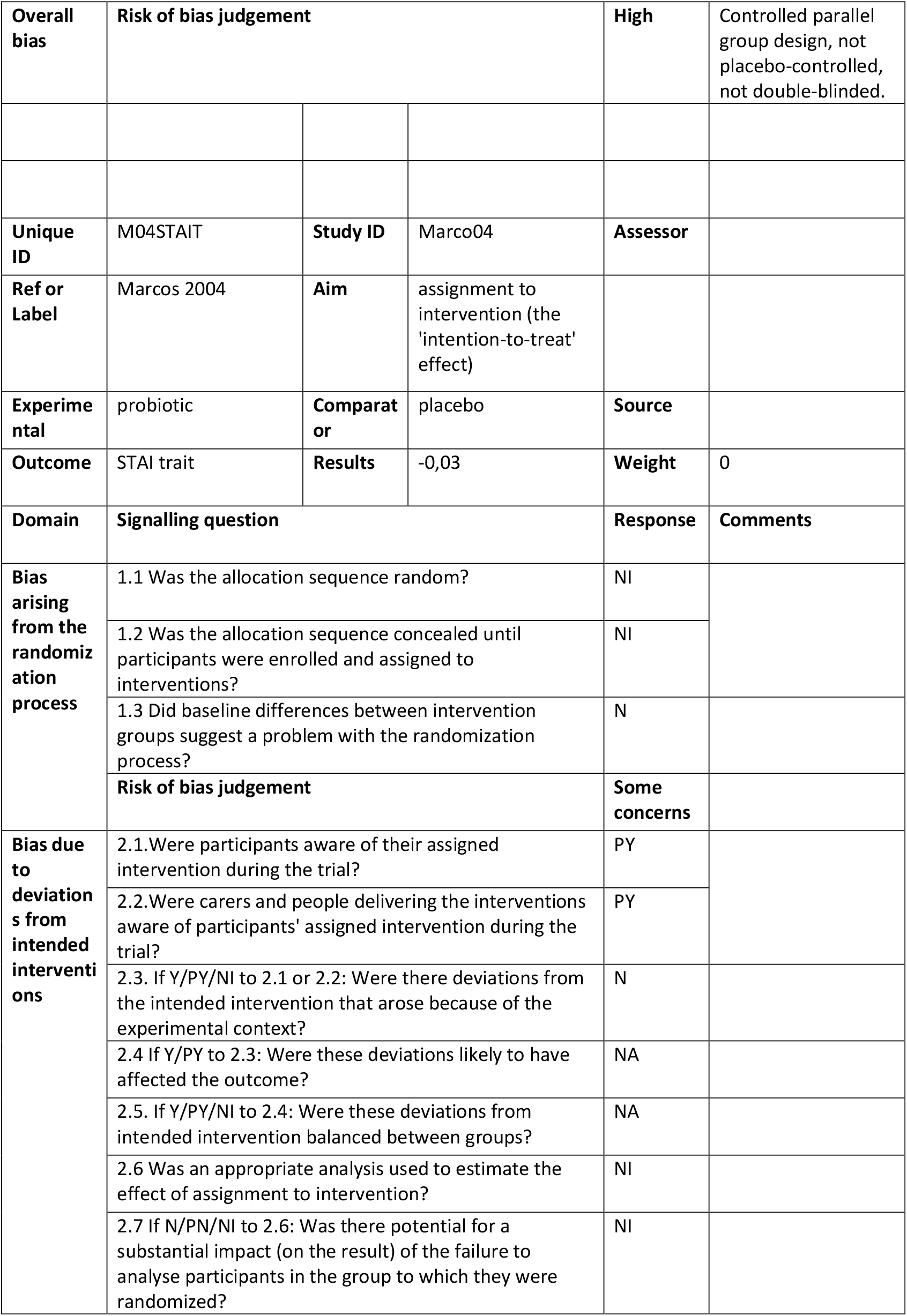
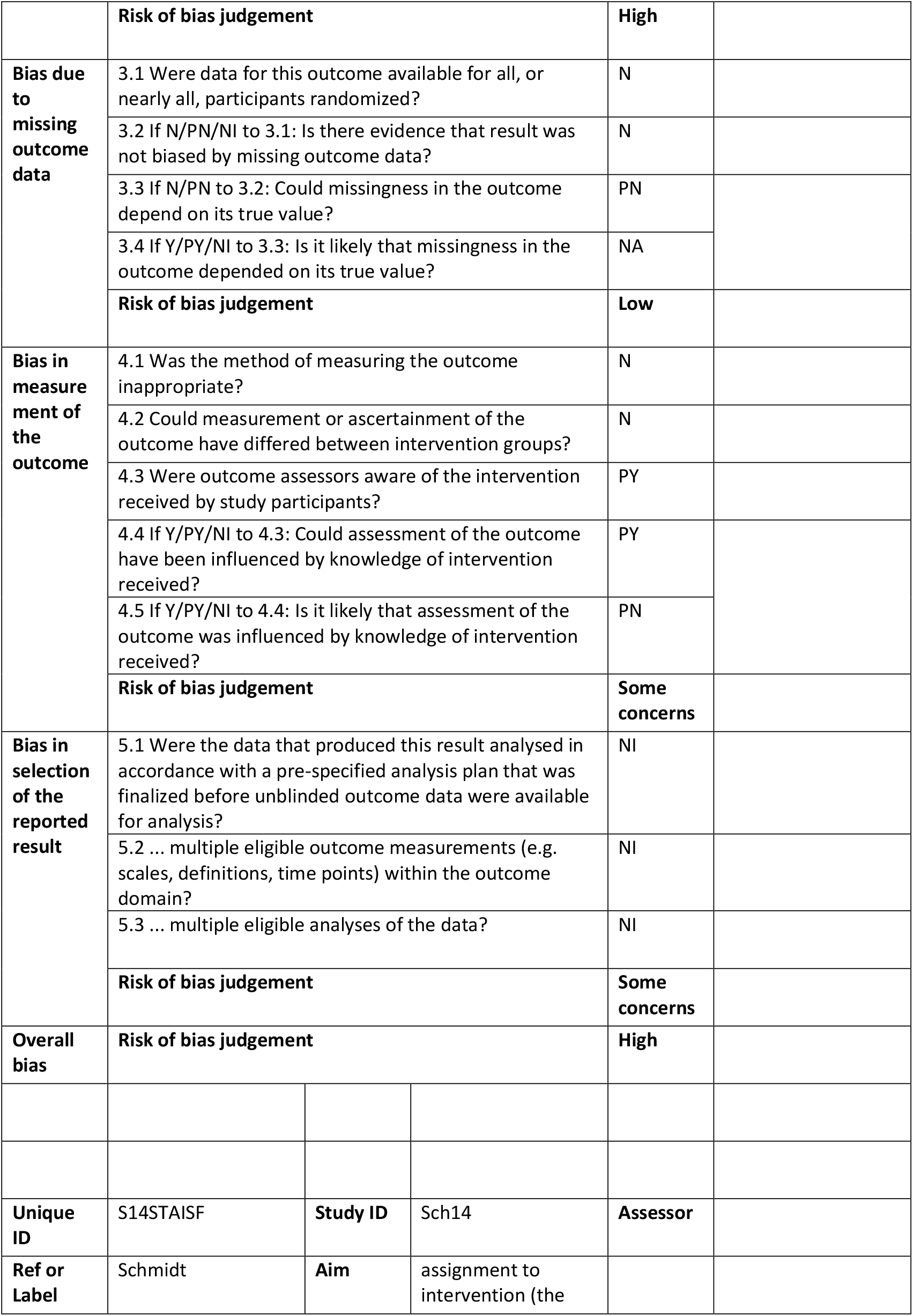
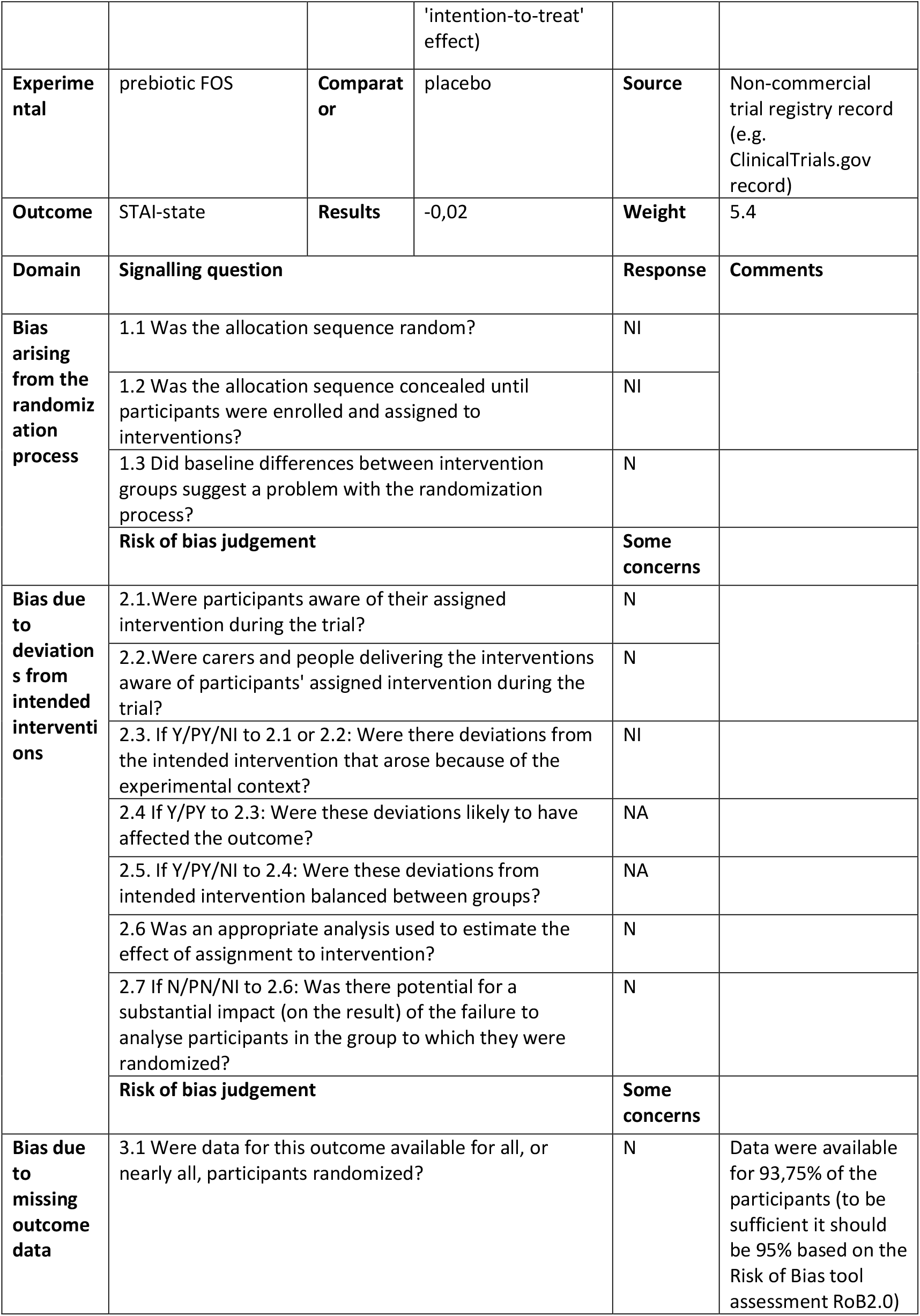
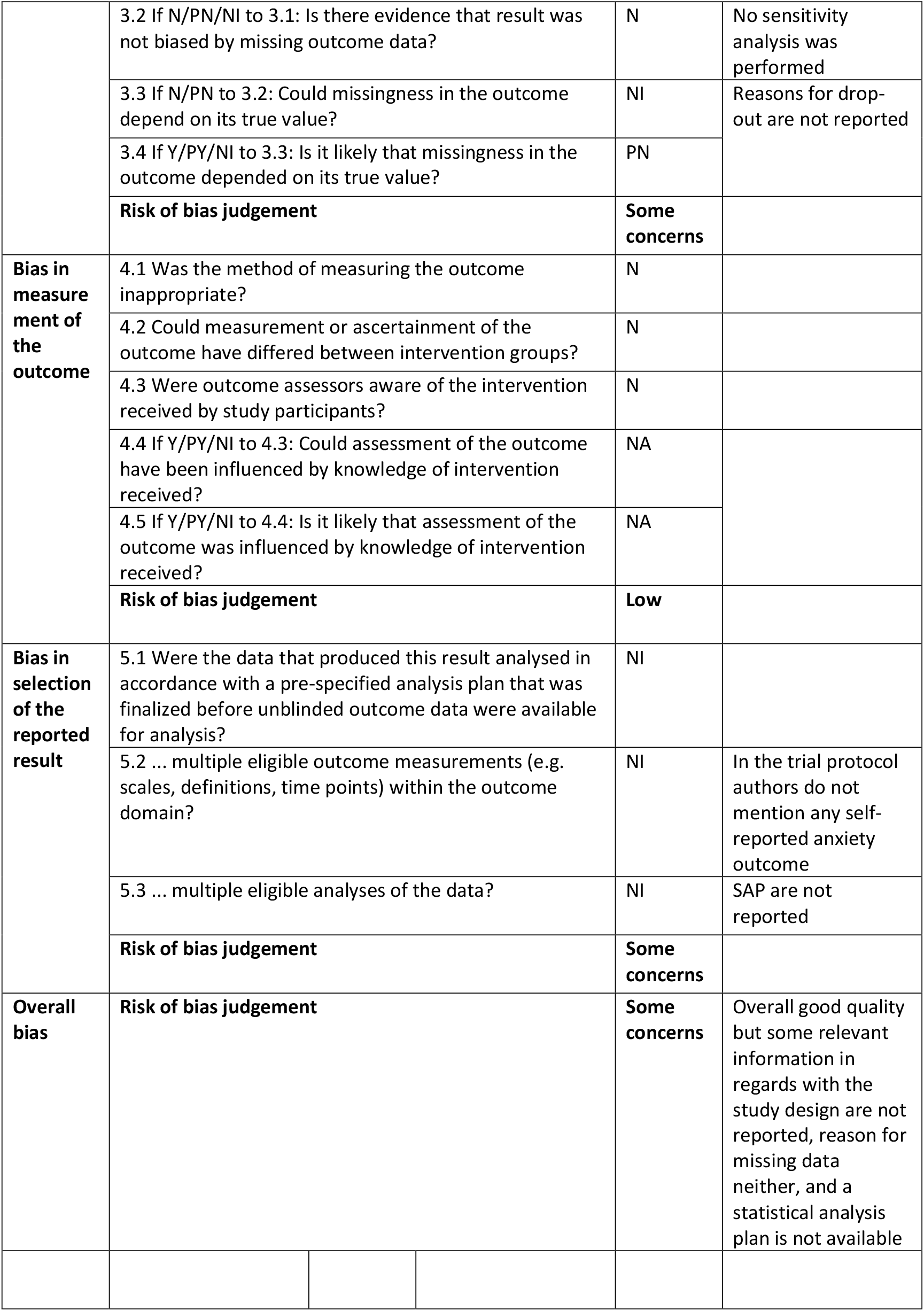
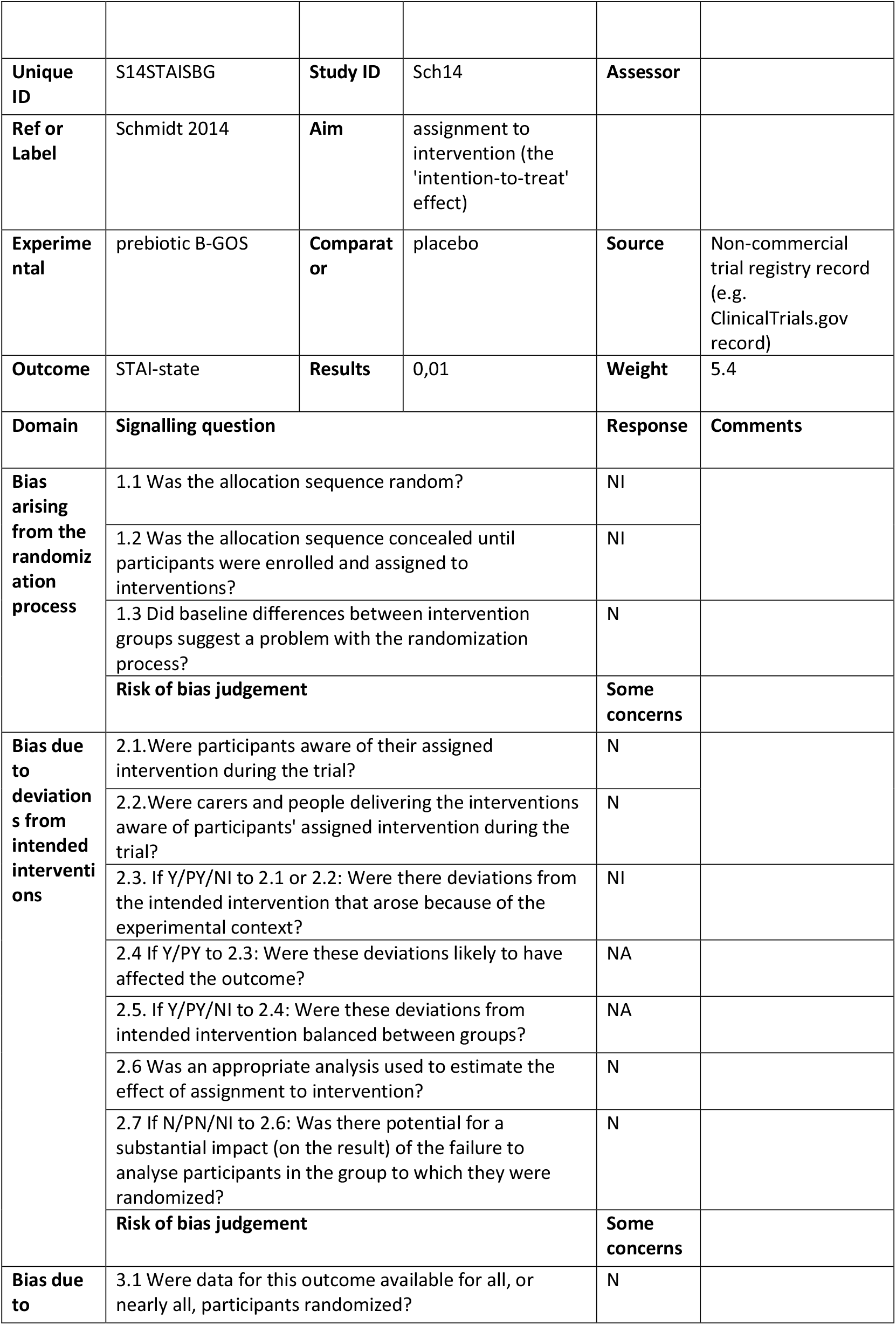
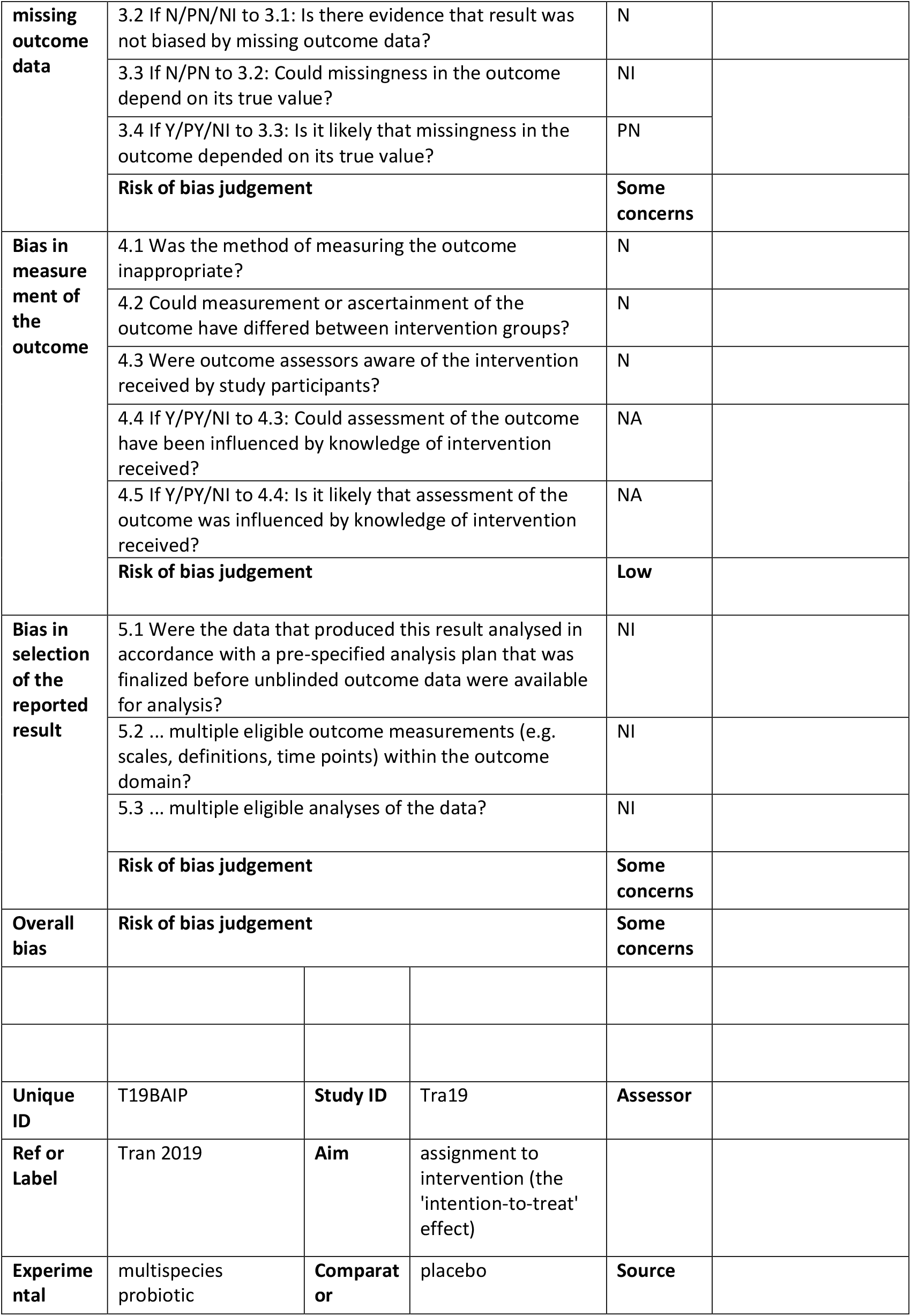
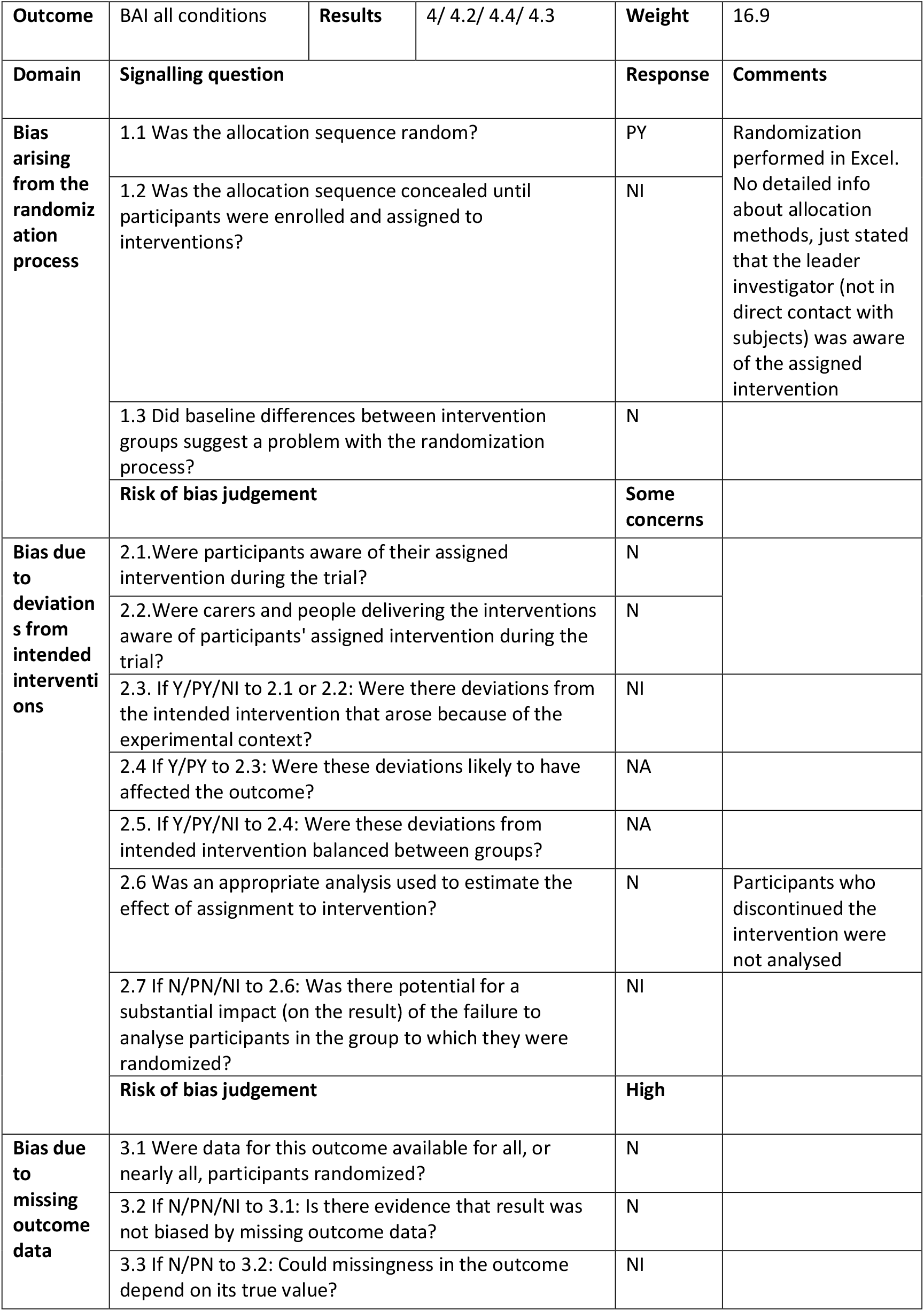
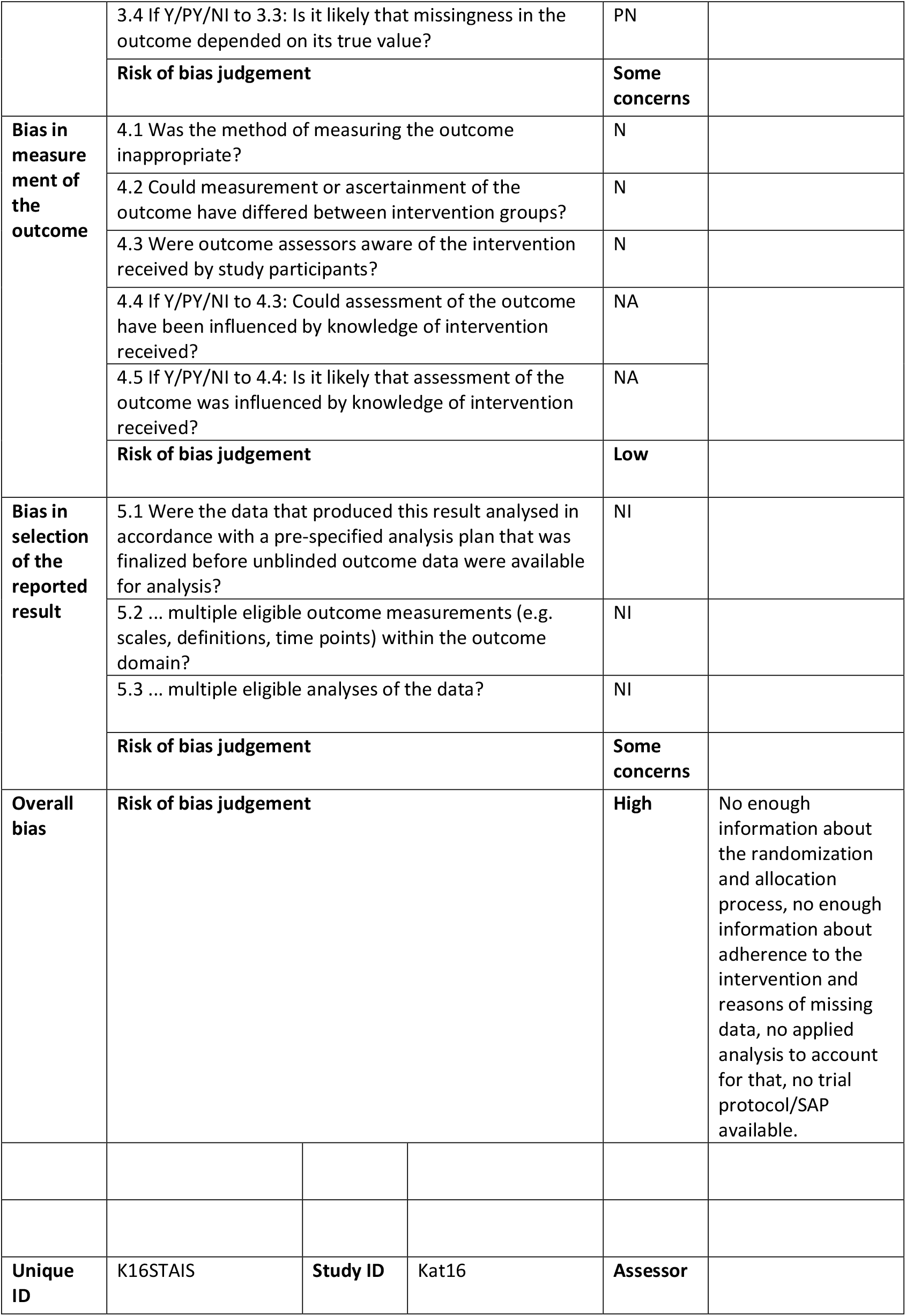
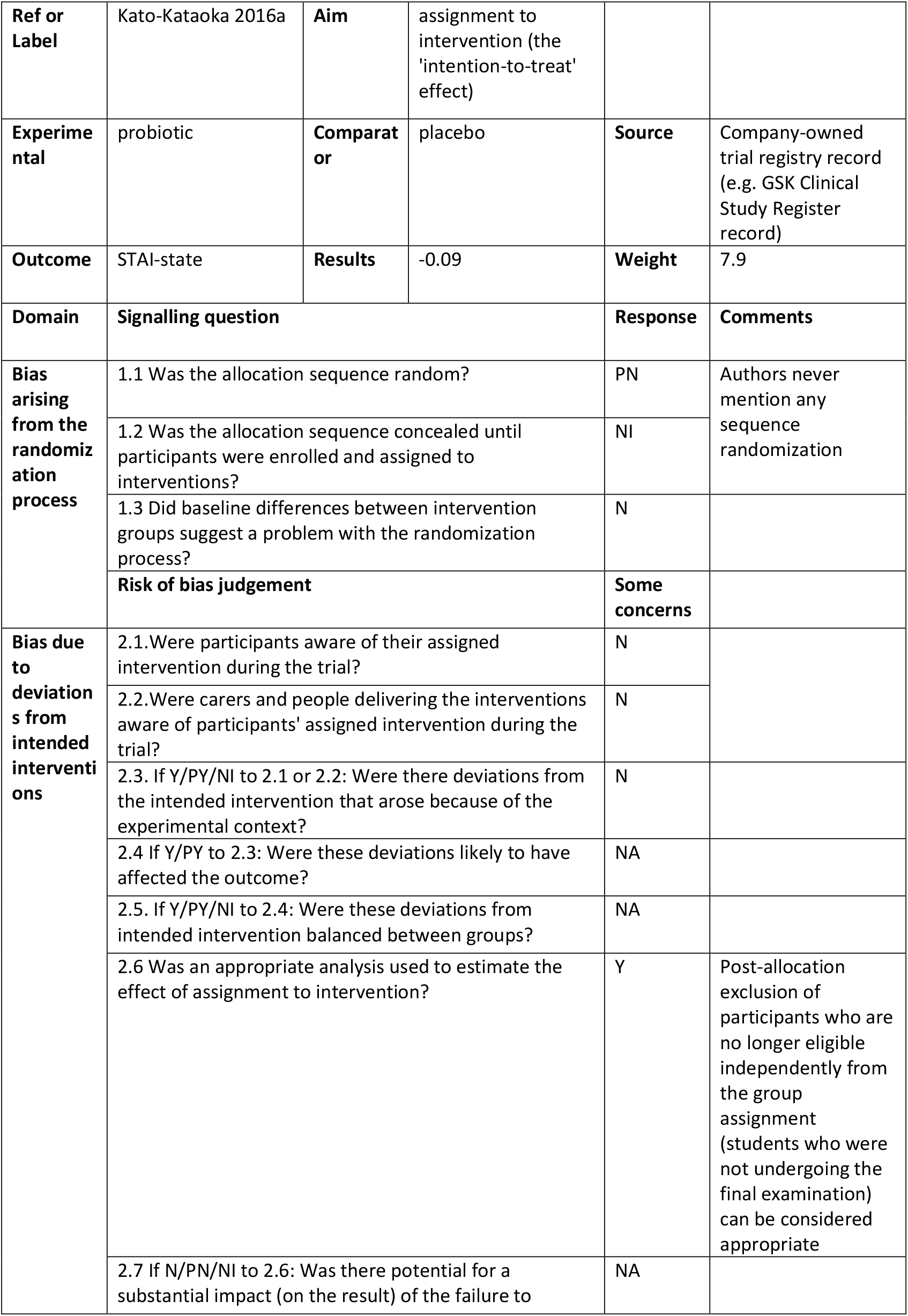
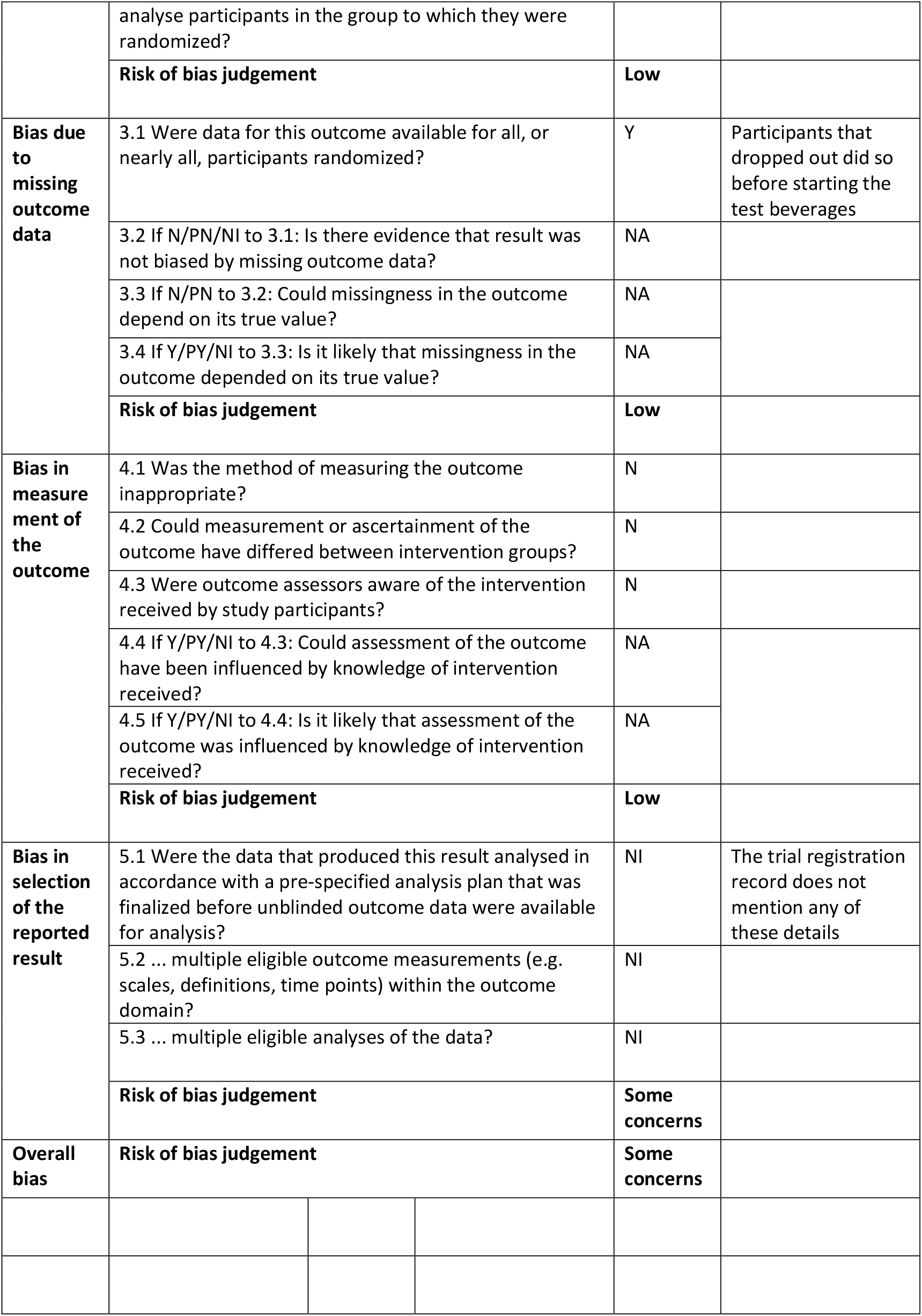
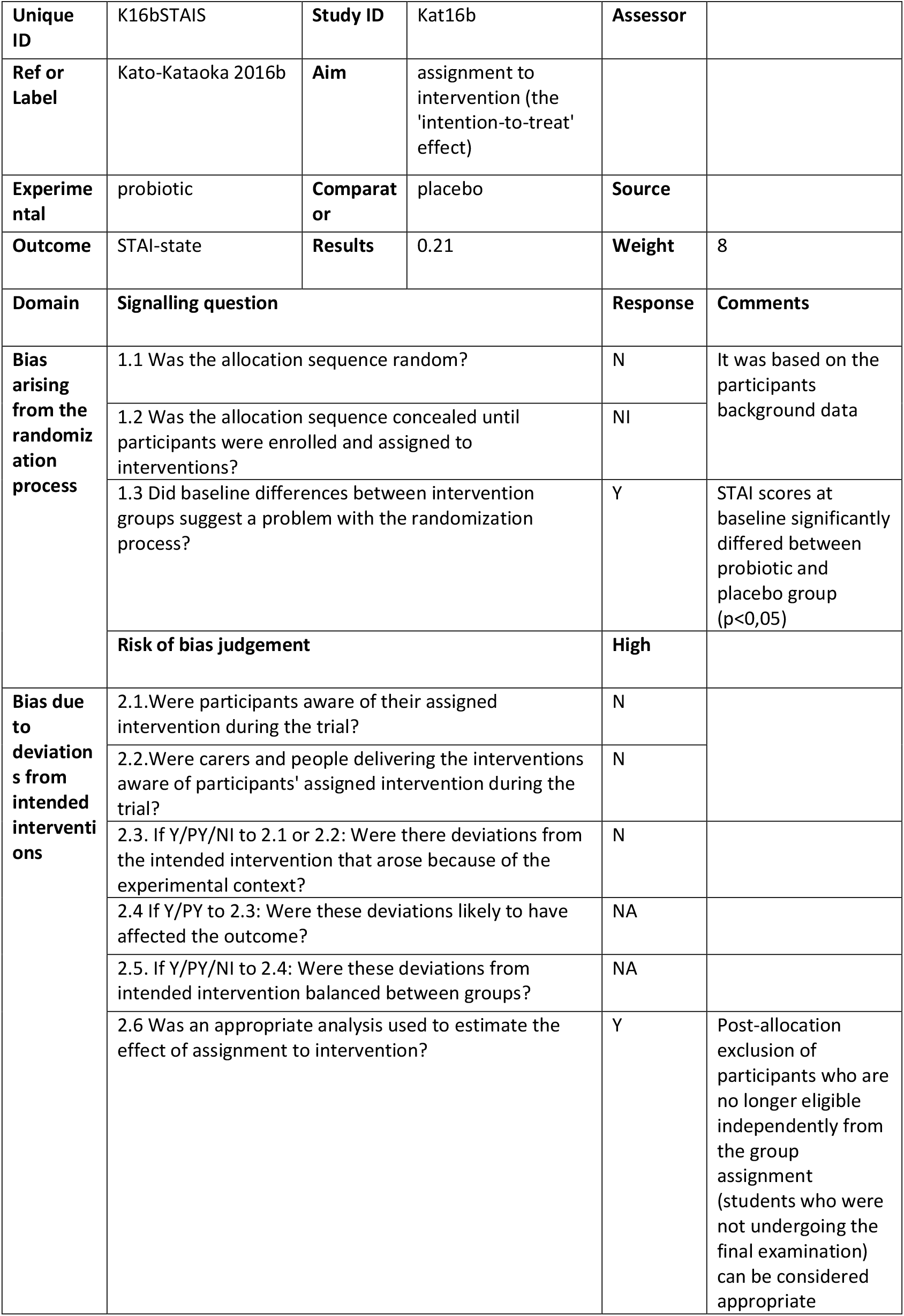
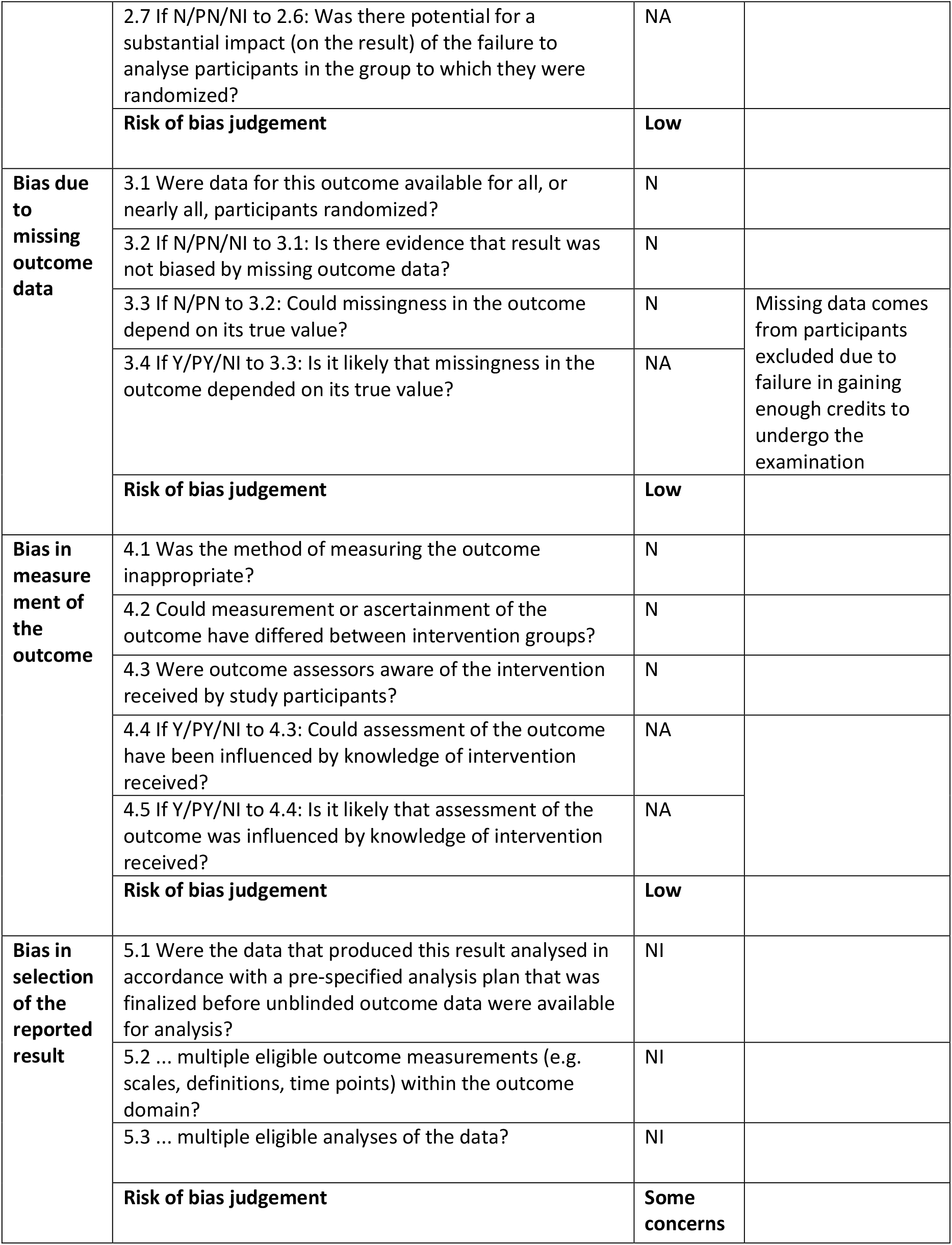
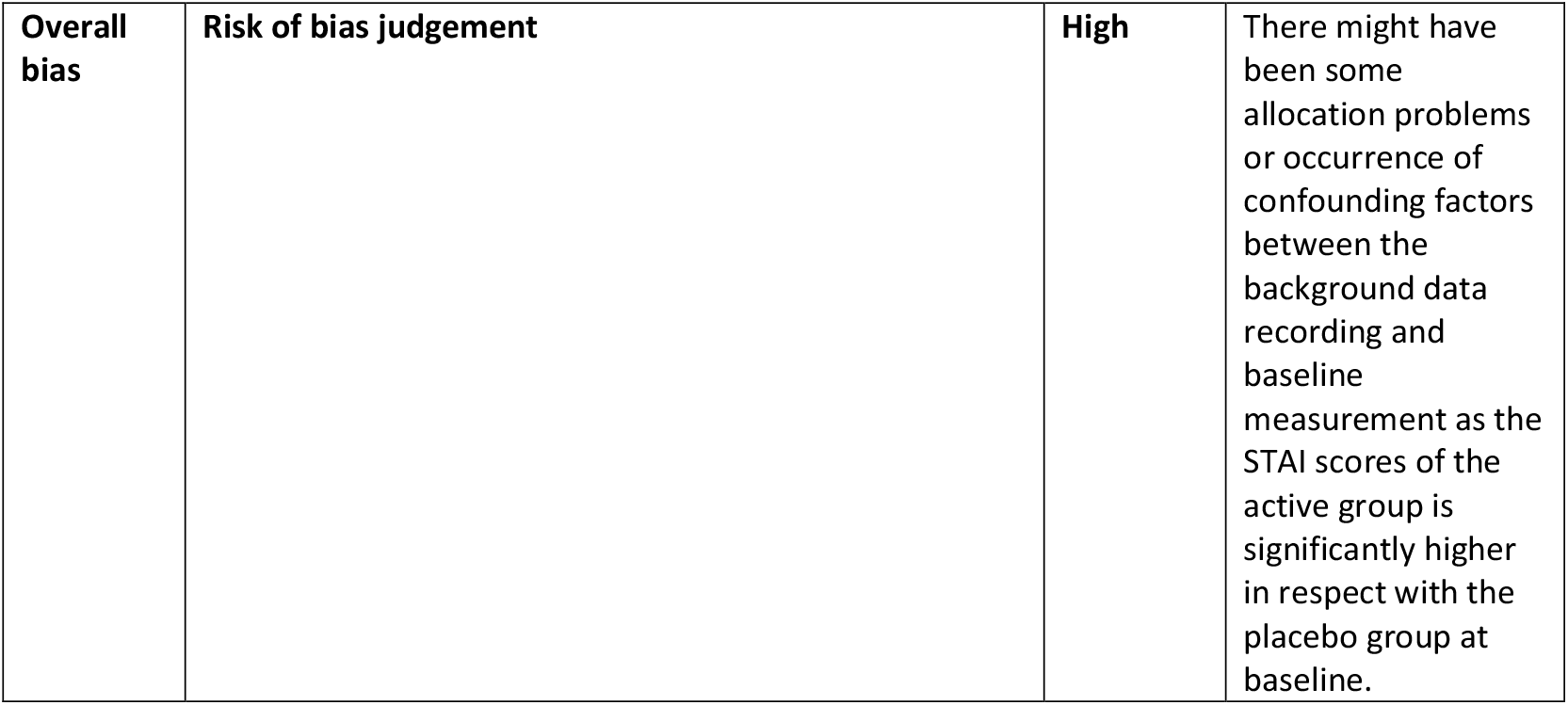

## APPENDIX 4: Risk of Bias Assessment of stress studies

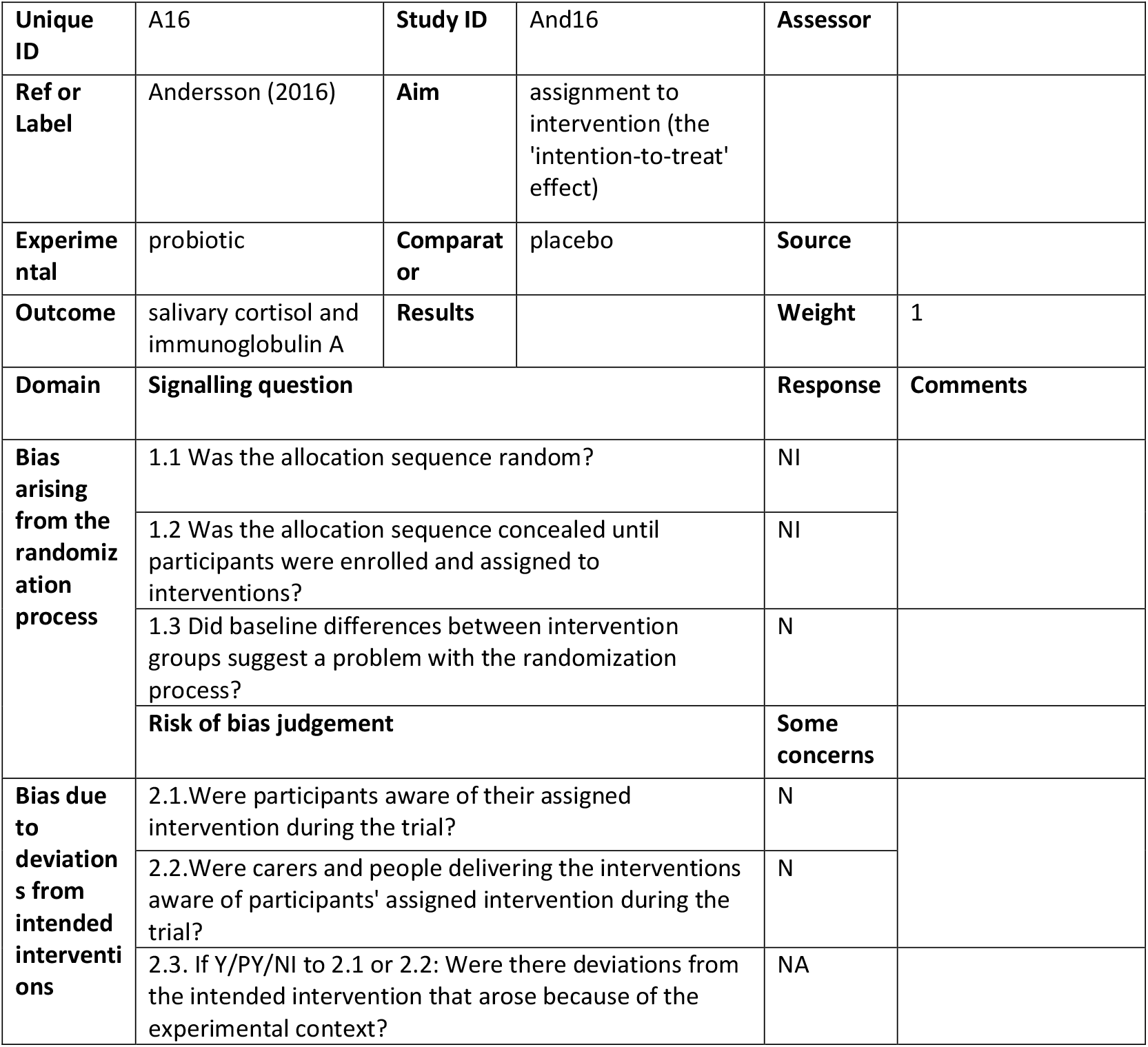
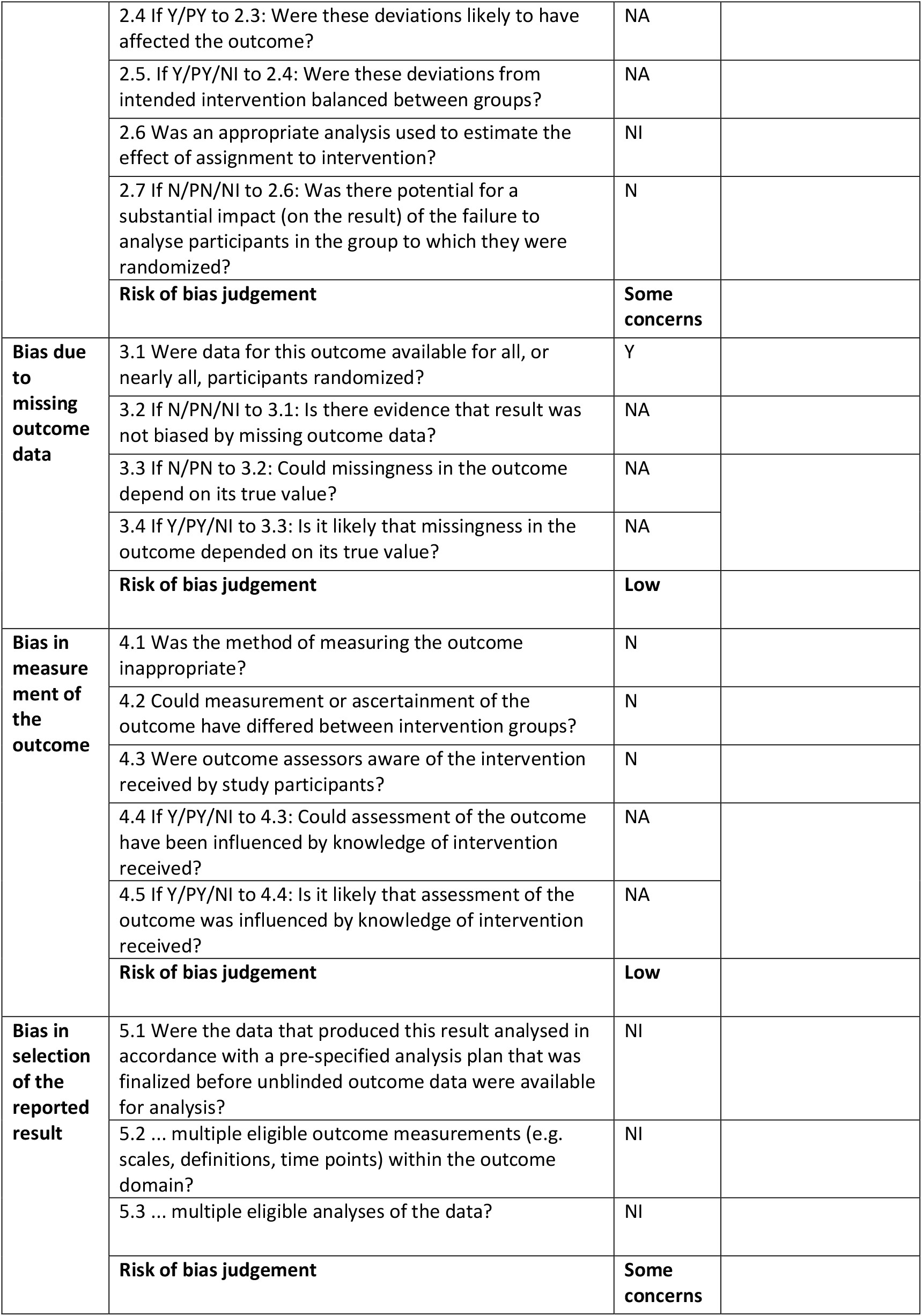
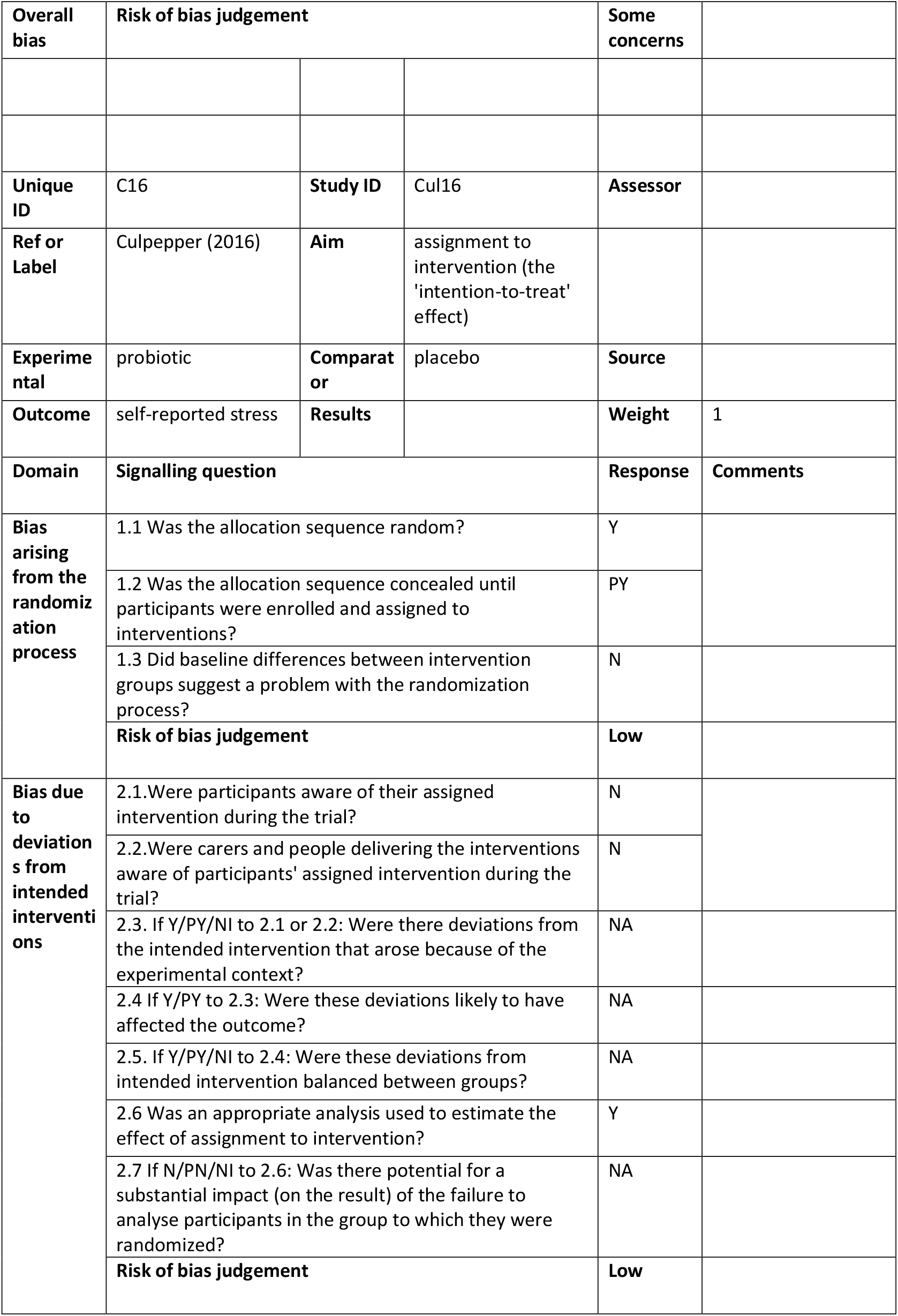
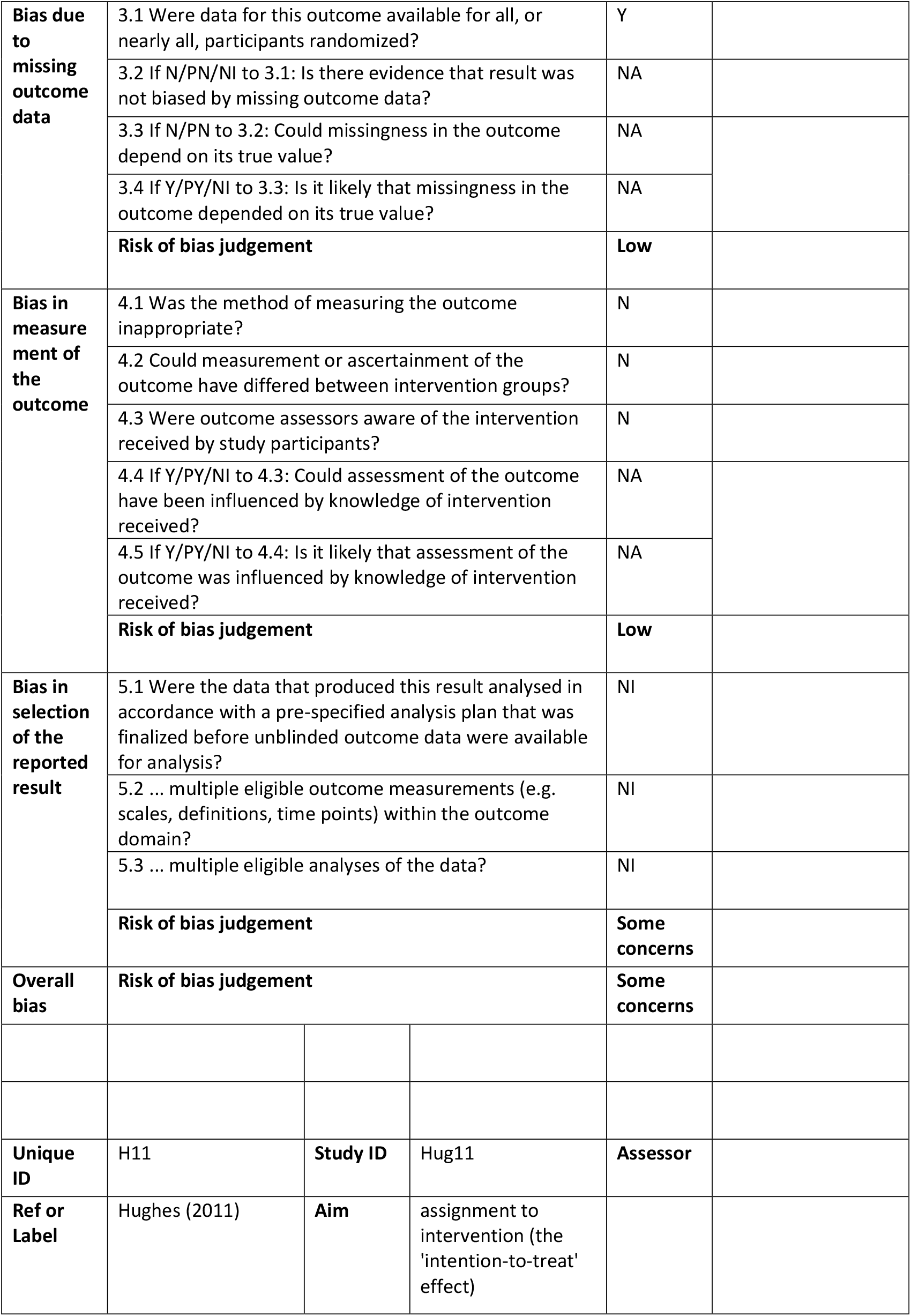
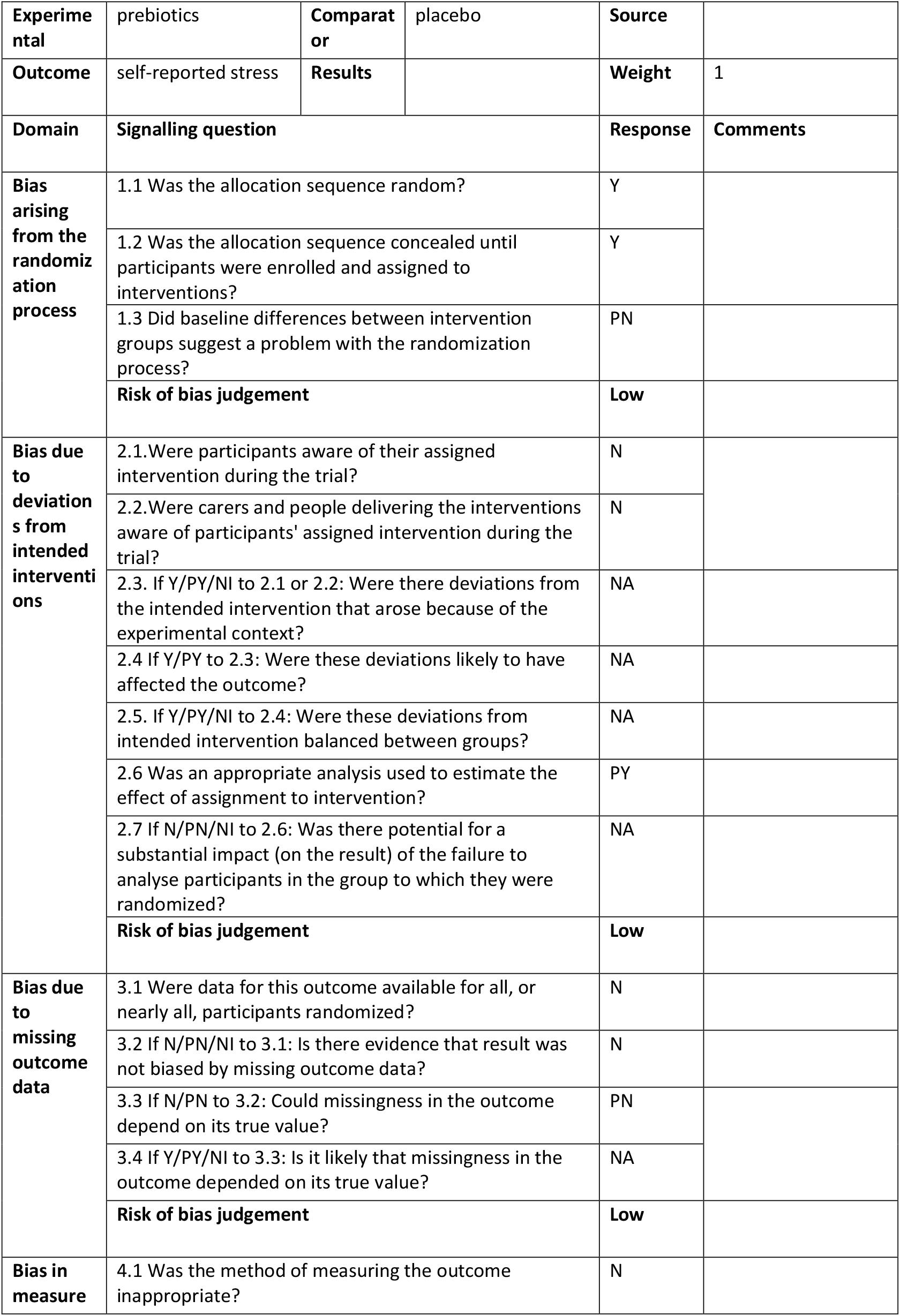
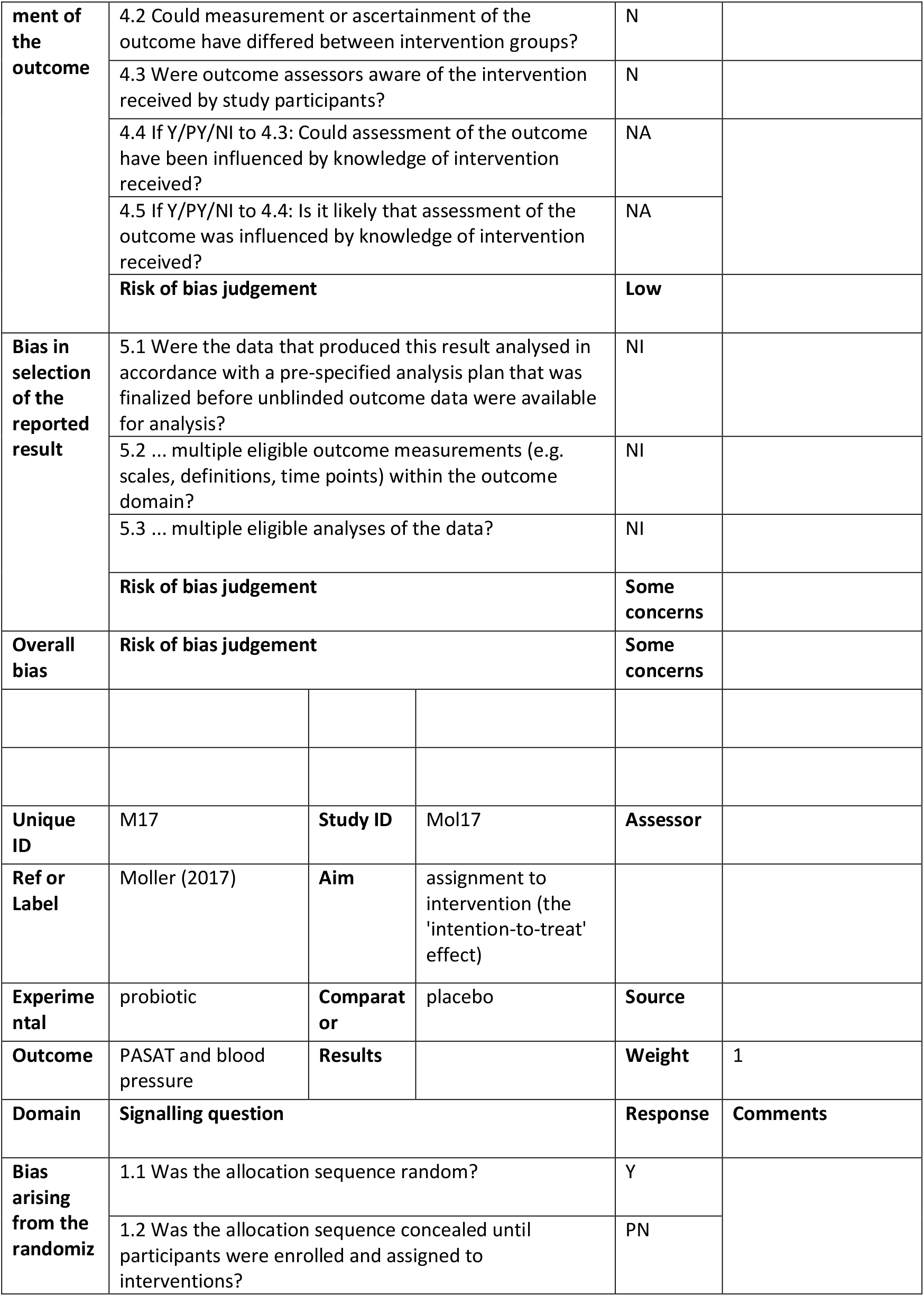
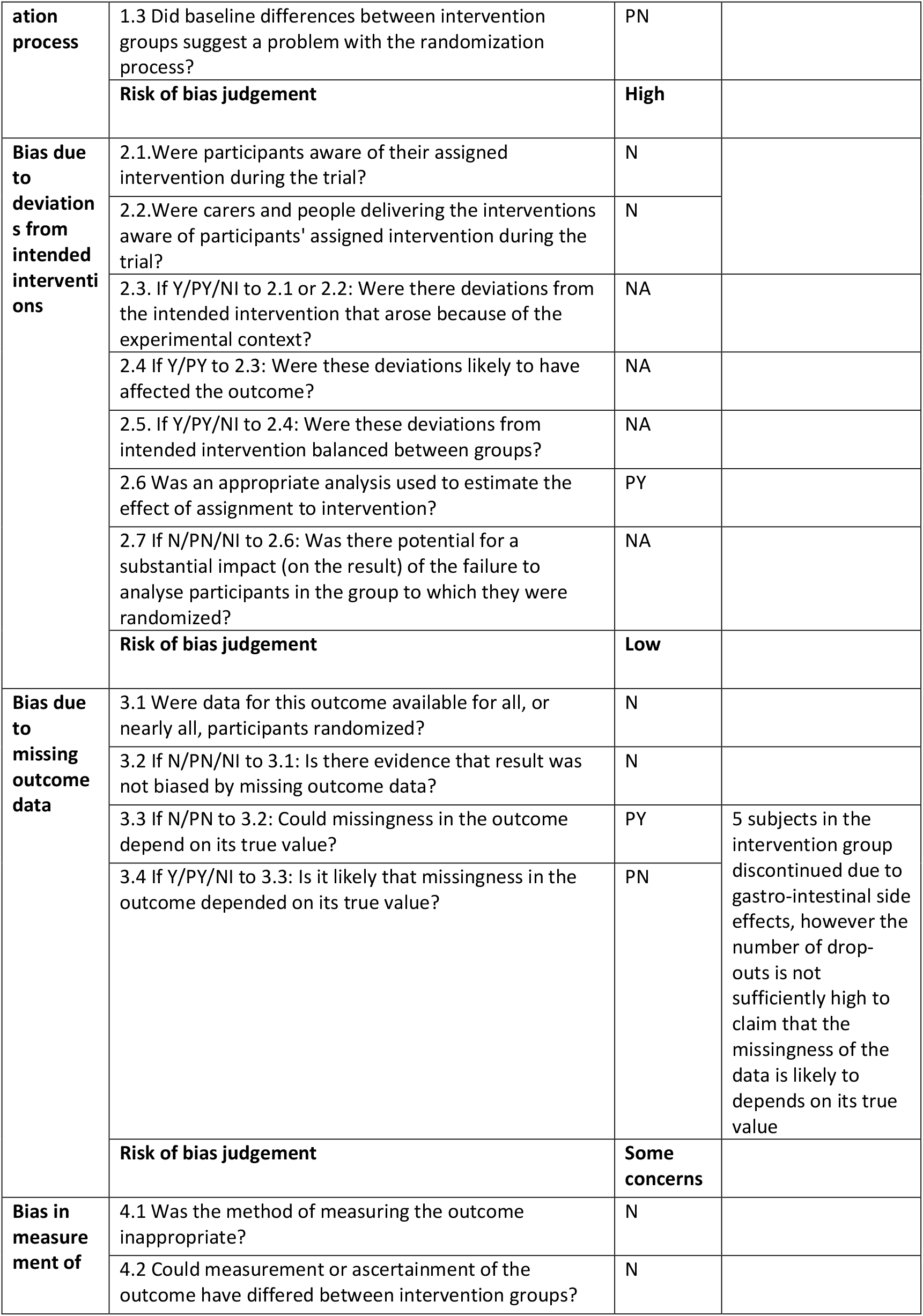
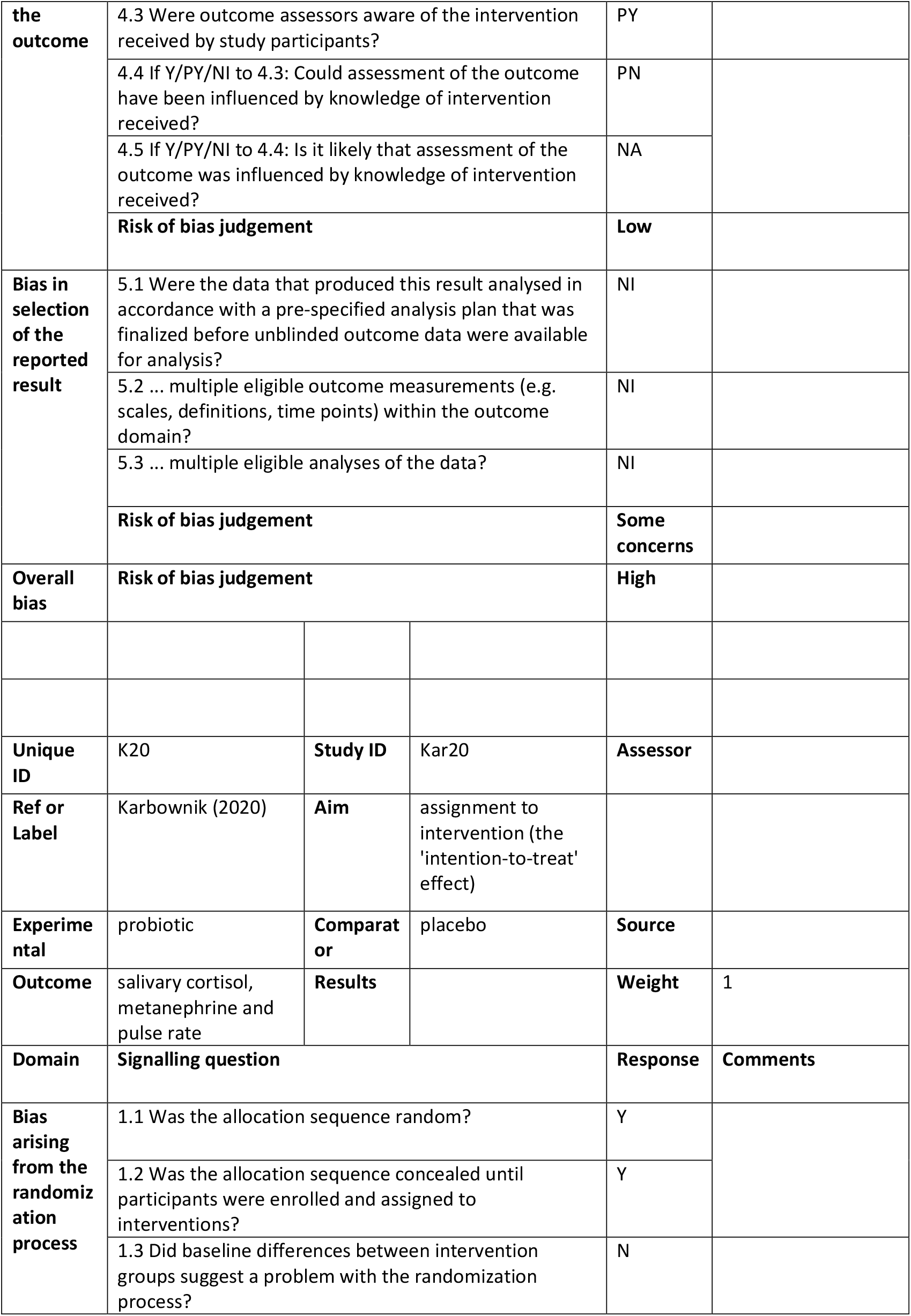
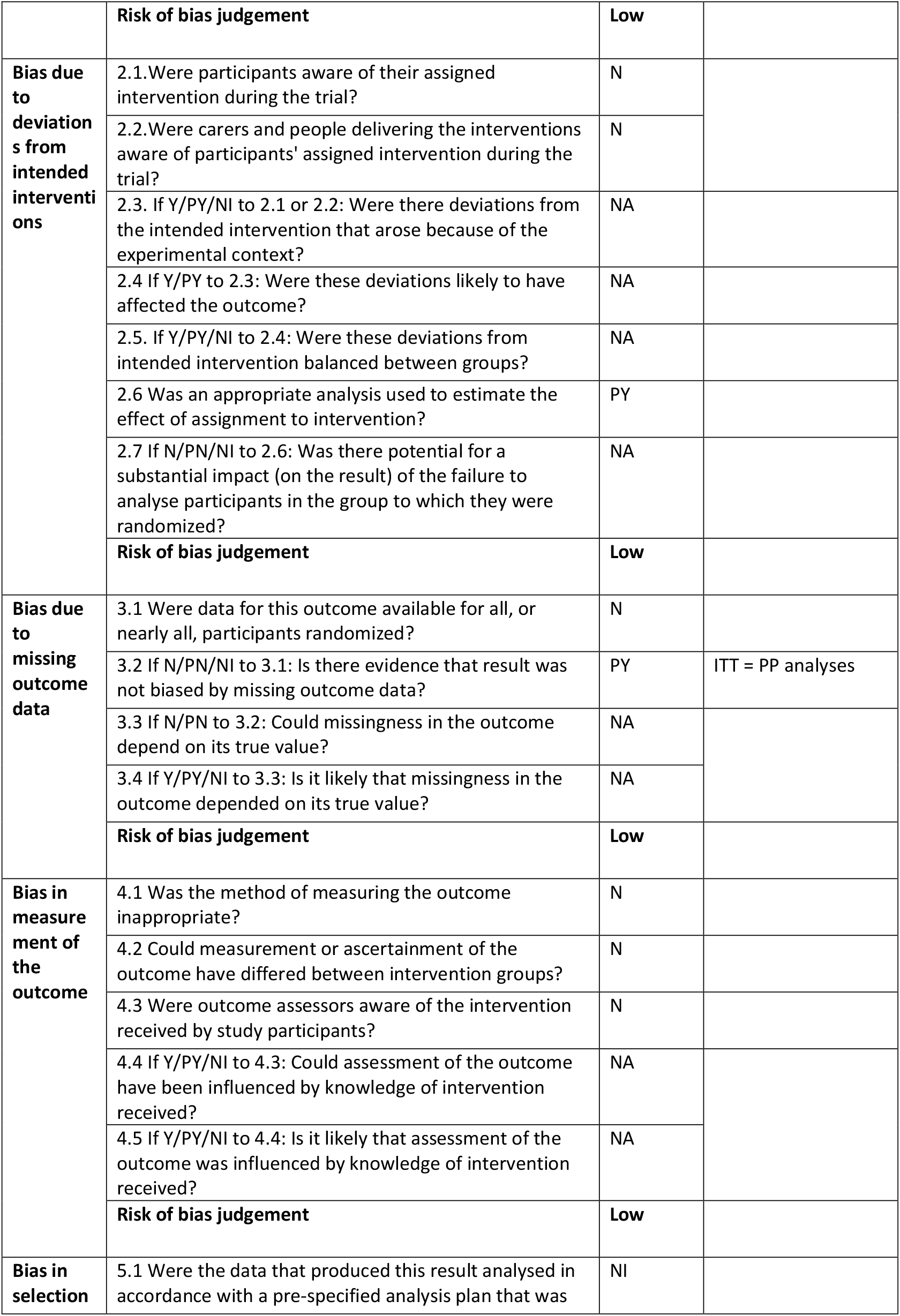
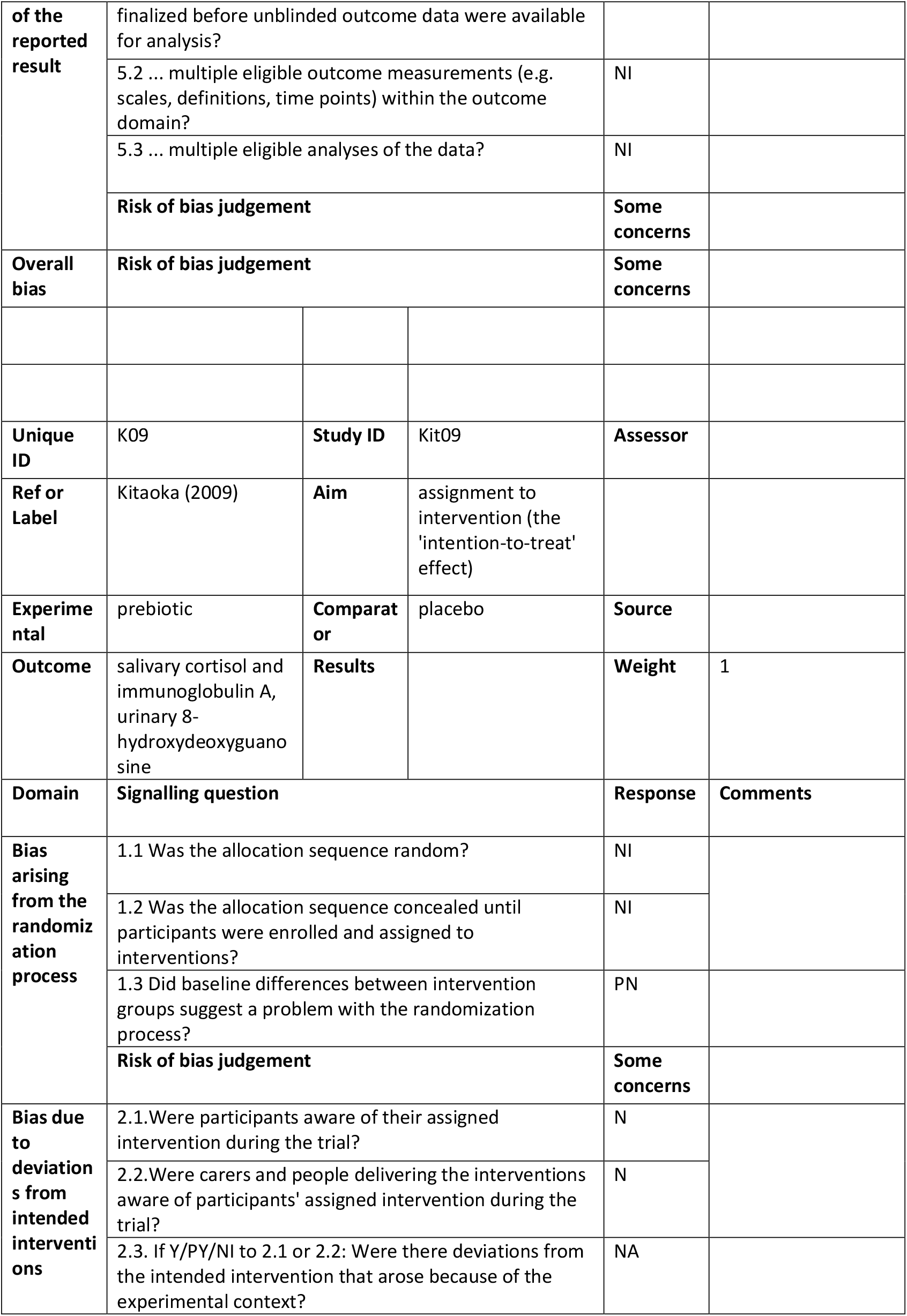
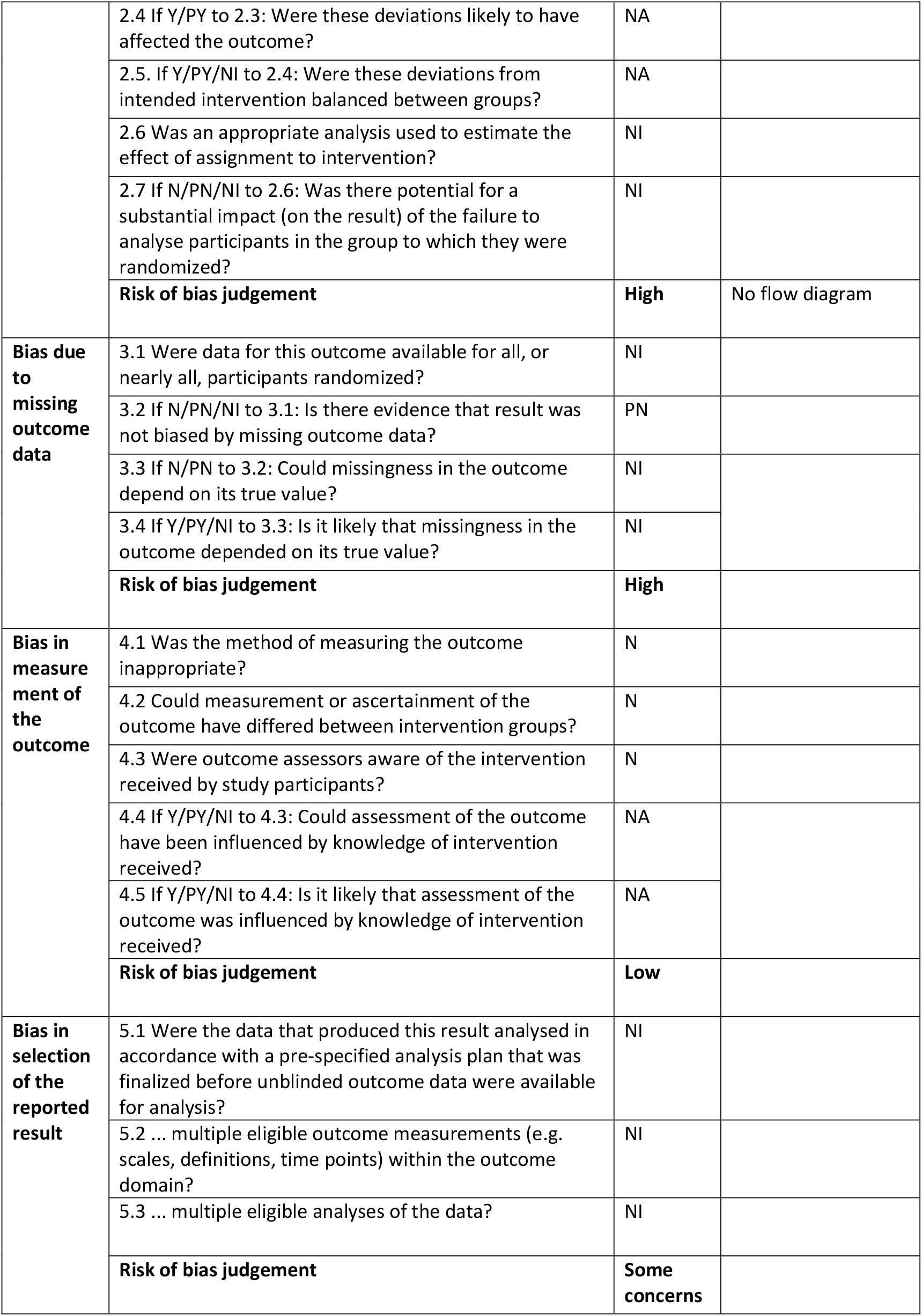
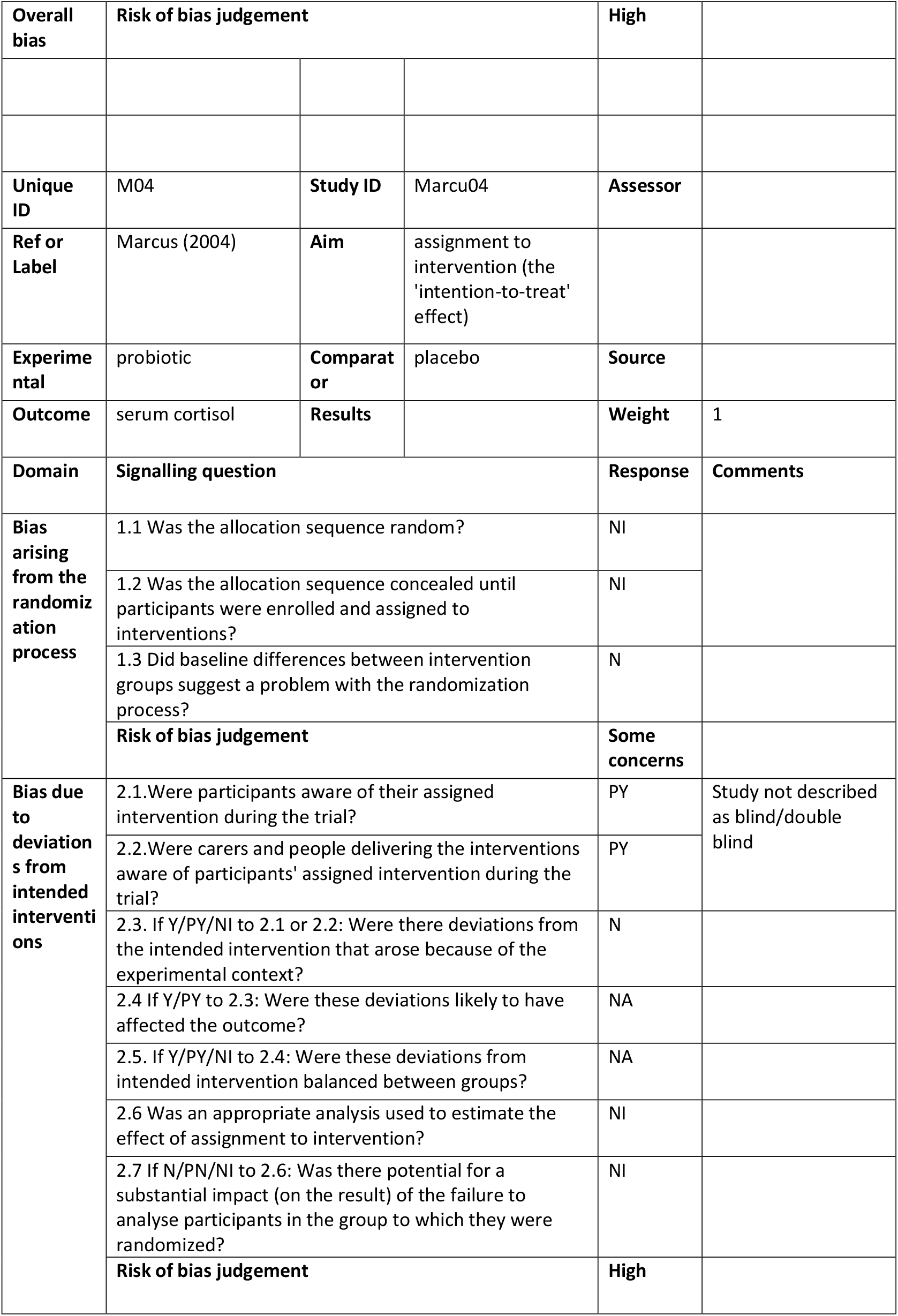
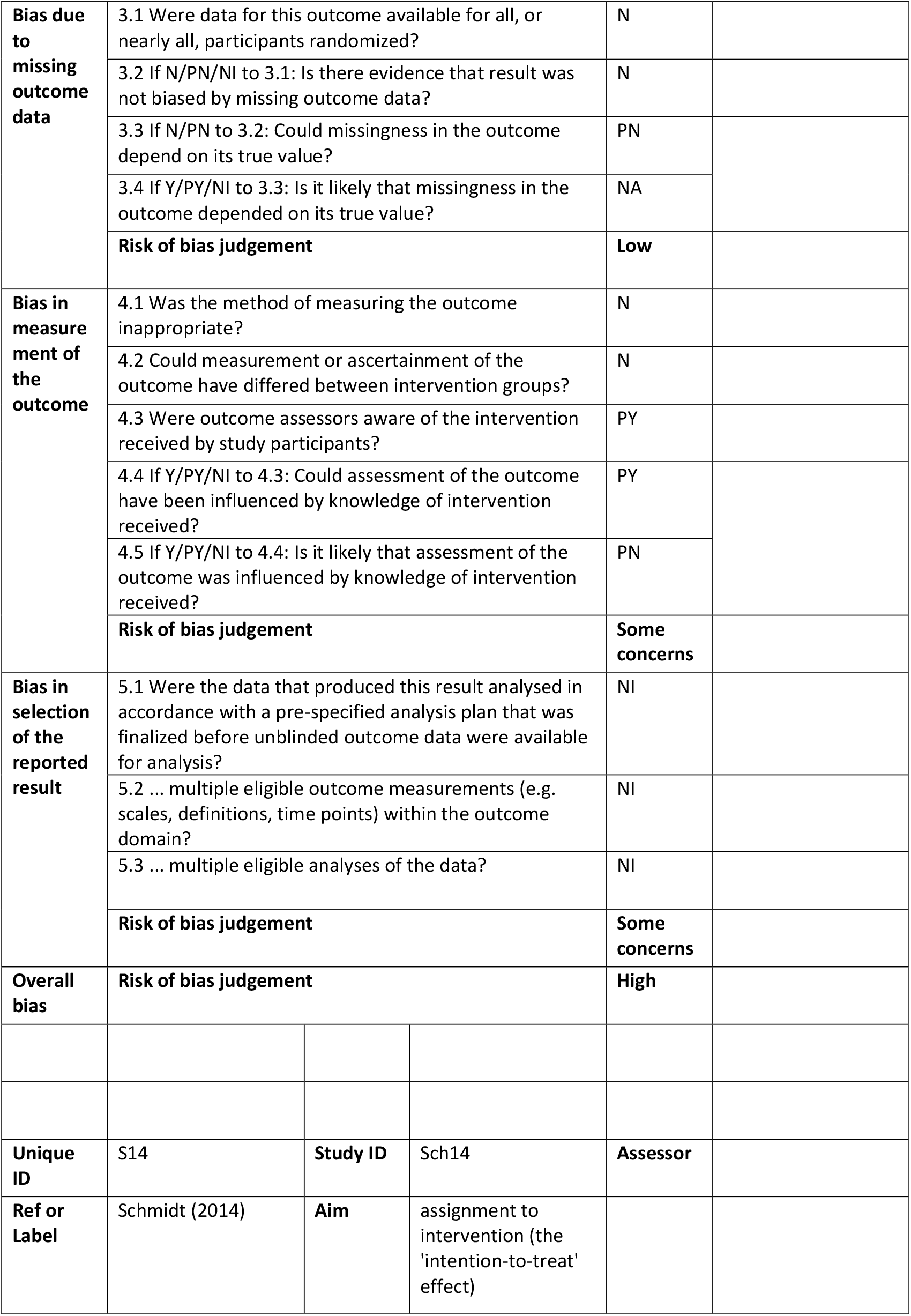
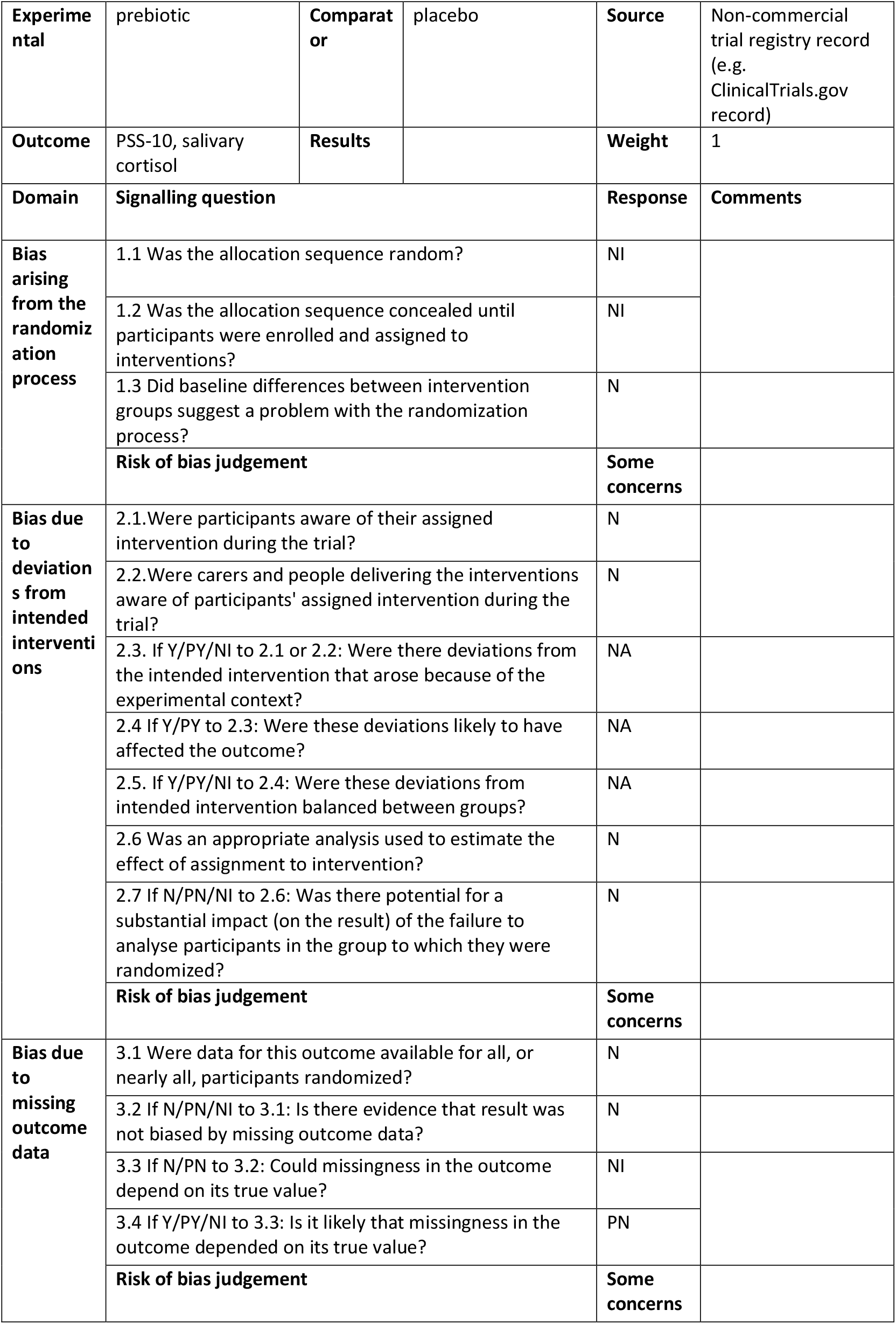
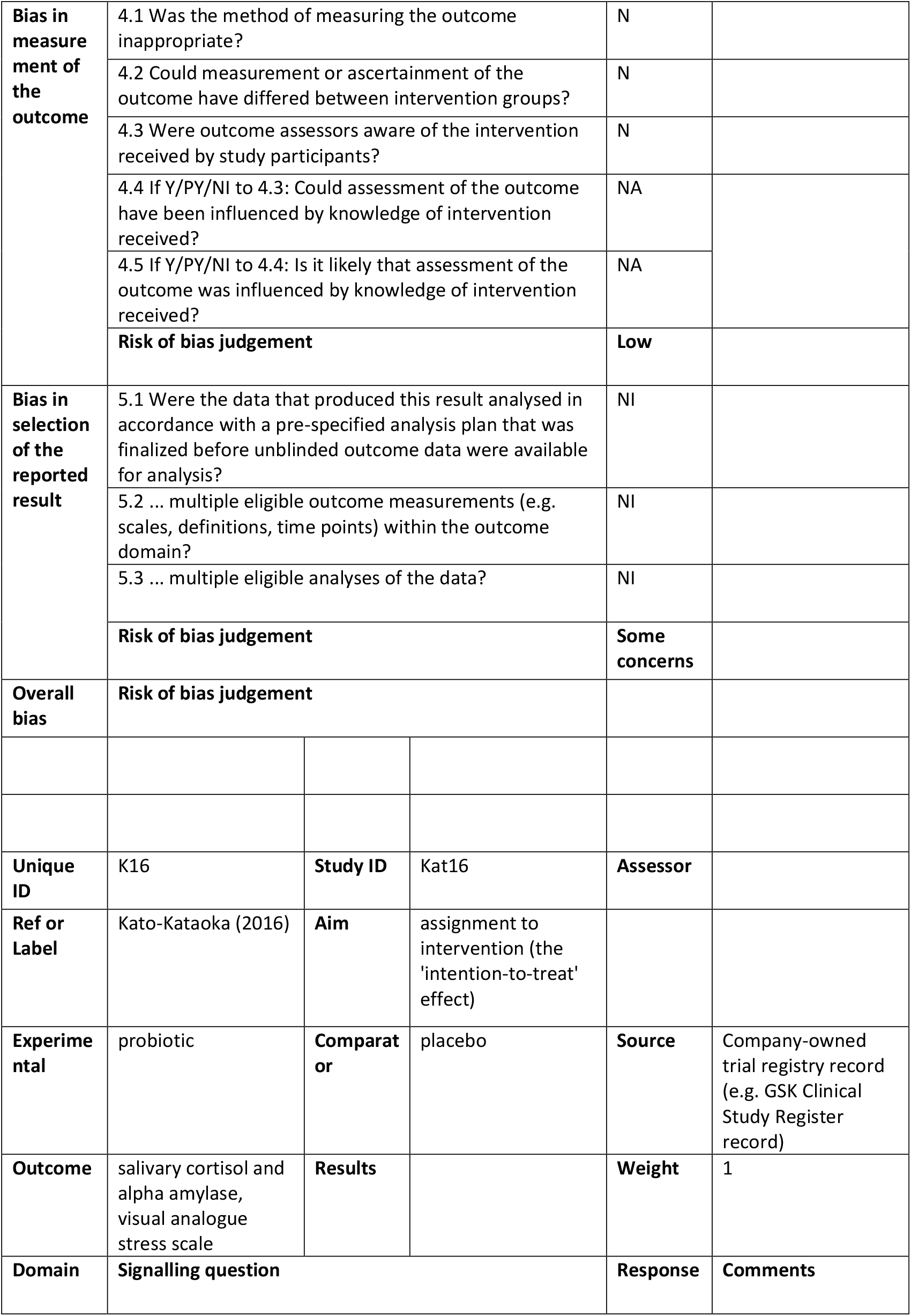
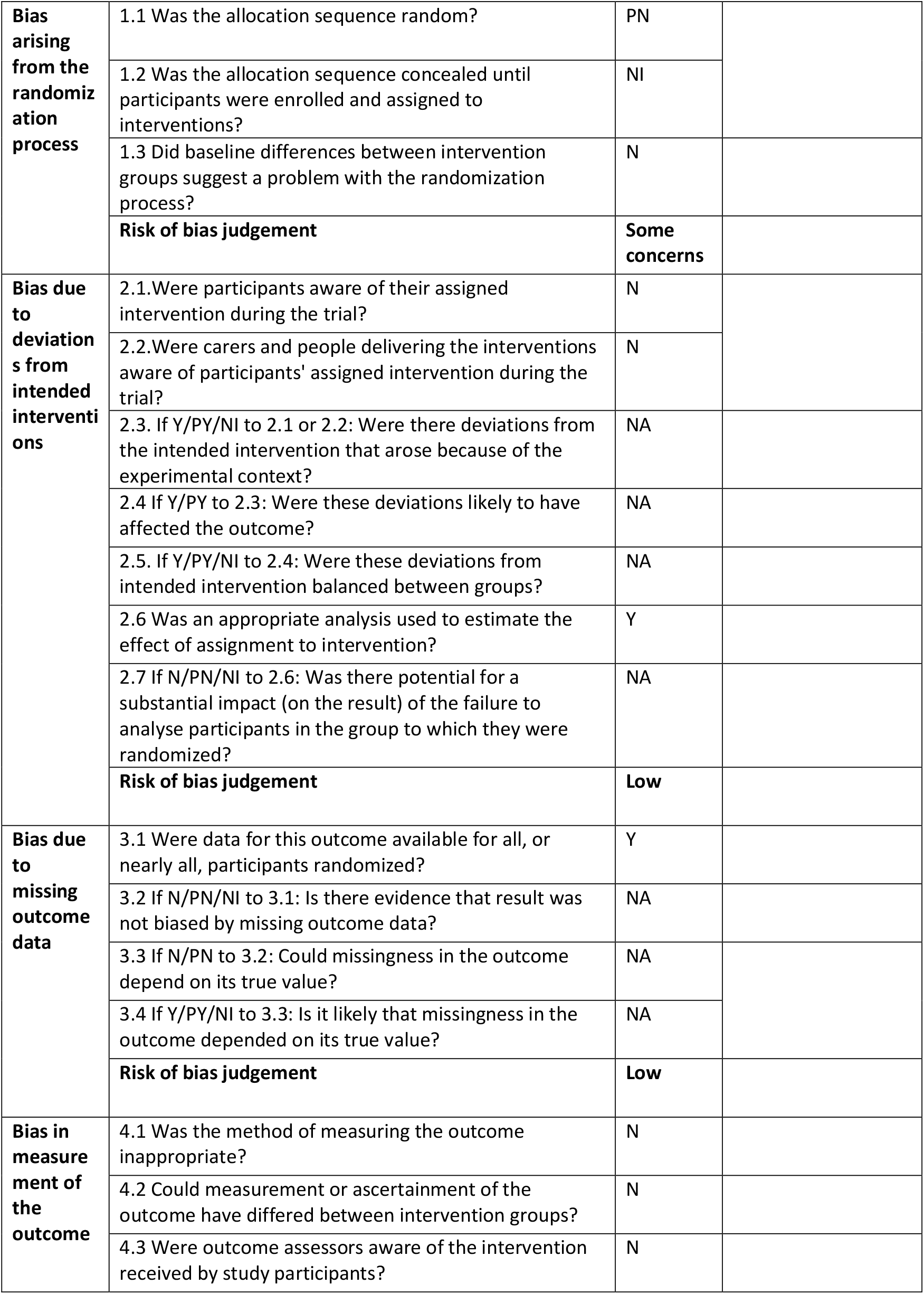
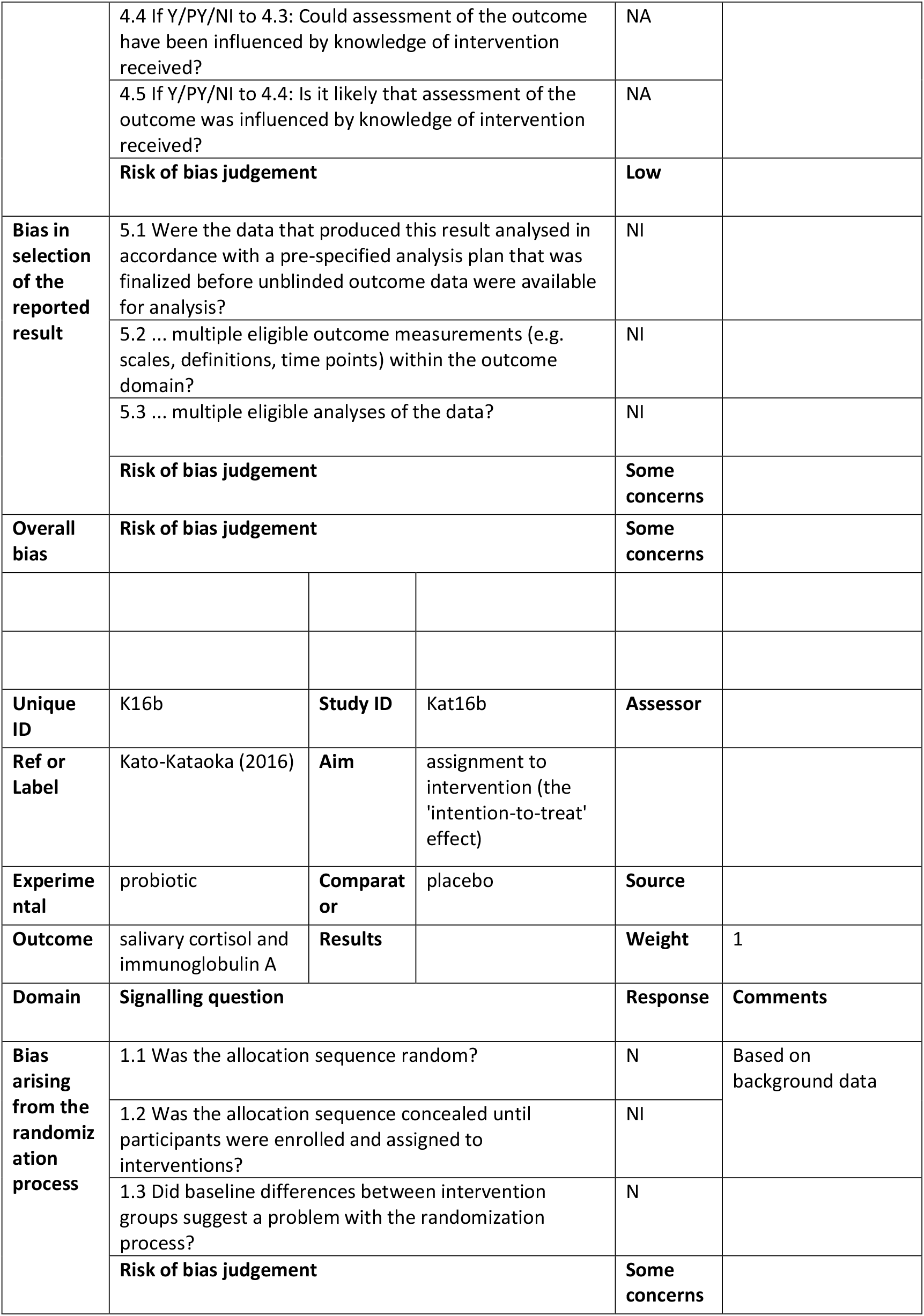
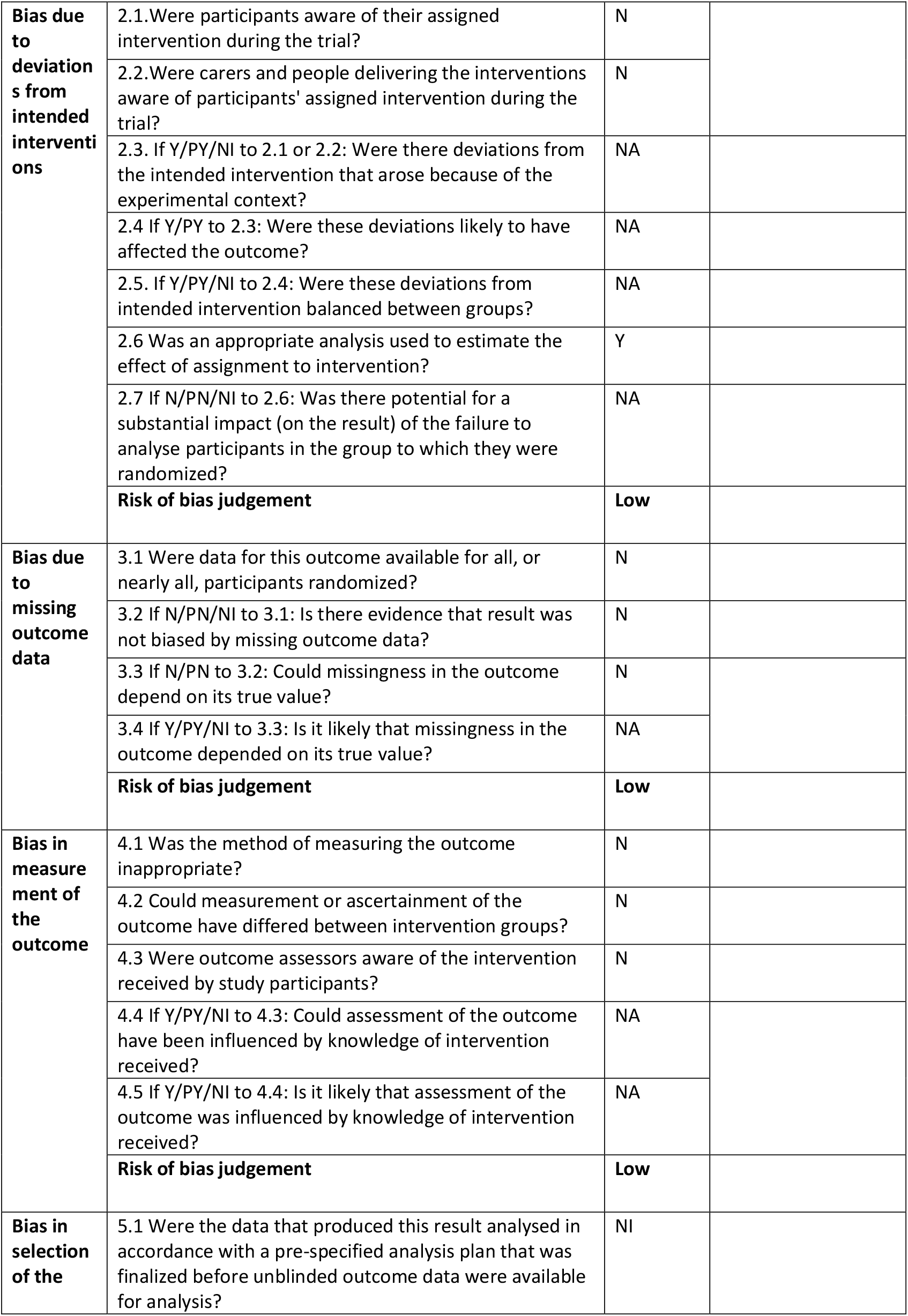
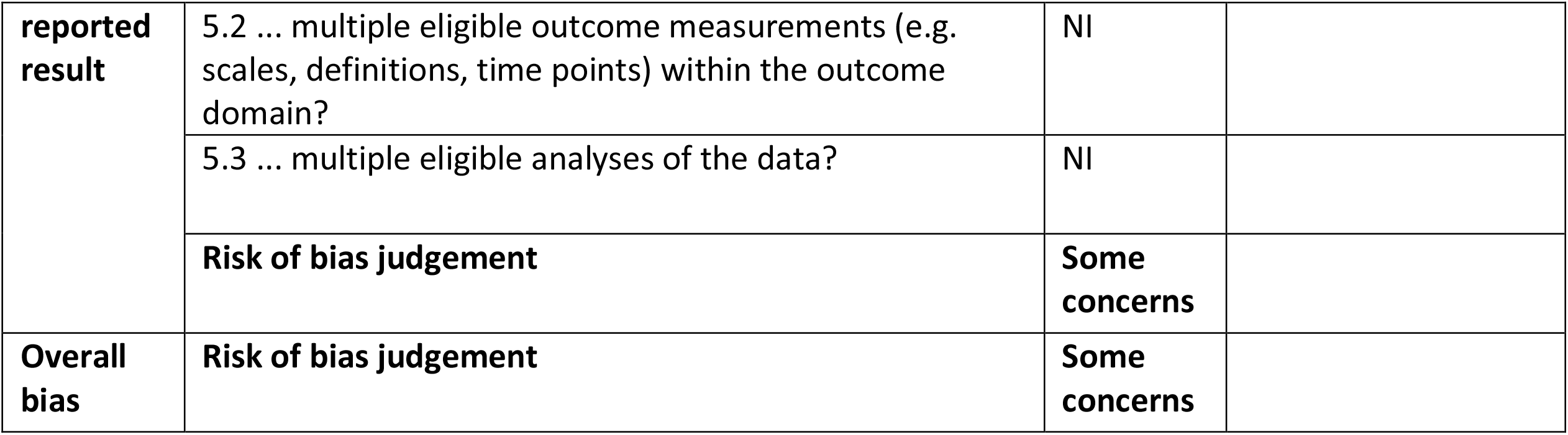

## Footnotes

1 All names have been changed

2 Note that while prebiotic status of PUFA is not yet universally accepted, new evidence supports the inclusion here: Costantini, L., R. Molinari, B. Farinon and N. Merendino (2017). “Impact of Omega-3 Fatty Acids on the Gut Microbiota.” <u>Int J Mol Sci</u> **18**(12).

